# Causal relationships between anthropometric traits, bone mineral density, osteoarthritis and spinal stenosis: a Mendelian randomization investigation

**DOI:** 10.1101/2023.08.10.23293938

**Authors:** Maria K Sobczyk, Benjamin G Faber, Lorraine Southam, Monika Frysz, April Hartley, Eleftheria Zeggini, The Genetics of Osteoarthritis Consortium, Haotian Tang, Tom R Gaunt

## Abstract

**Background:** Spinal stenosis is a common condition among older individuals, with significant morbidity attached. Little is known about its risk factors but degenerative conditions, such as osteoarthritis (OA) have been identified for their mechanistic role. This study aims to explore causal relationships between anthropometric risk factors, osteoarthritis, and spinal stenosis using Mendelian randomization (MR) techniques.

**Methods:** We applied two-sample univariable and multivariable MR to investigate the causal relationships between genetic liability for select risk factors (including adiposity and skeletal traits) and spinal stenosis. Next, we examined the genetic relationship between osteoarthritis and spinal stenosis with LD score regression and CAUSE MR method. Using multivariable MR, osteoarthritis and BMI were then tested as potential mediators of the causal pathways identified.

**Results:** Our analysis revealed strong evidence for the effect of higher BMI (OR=1.54, 95% CI: 1.41-1.69, p-value=2.7 x 10^-21^), waist (OR=1.43, 95% CI: 1.15-1.79, p-value=1.5 x 10^-3^) and hip (OR=1.50, 95% CI: 1.27-1.78, p-value=3.3 x 10^-6^) circumference on spinal stenosis. Strong associations were observed for higher bone mineral density (BMD): total body (OR=1.21, 95% CI: 1.12-1.29, p-value=1.6 x 10^-7^), femoral neck (OR=1.35, 95% CI: 1.09-1.37, p-value=7.5 x 10^-7^), and lumbar spine (OR=1.38, 95% CI: 1.25-1.52, p-value=4.4 x 10^-11^). We detected high genetic correlations between spinal stenosis and osteoarthritis (rg range: 0.47-0.66), with Bayesian CAUSE results supporting a causal effect of osteoarthritis on spinal stenosis (OR_all OA_=1.6, 95% CI:1.41-1.79). Direct effects of BMI, total body/femoral neck/lumbar spine BMD on spinal stenosis remained after adjusting for osteoarthritis and/or BMI in the multivariable MR.

**Conclusions:** Genetic susceptibility to anthropometric risk factors, particularly higher BMI and bone mineral density can increase the risk of spinal stenosis, independent of osteoarthritis status. These results improve our understanding of spinal stenosis aetiology and may inform preventative strategies and treatments.

## Introduction

Spinal stenosis is a potentially debilitating condition with symptomatic spinal stenosis affecting about10% of Western populations and prevalence only increasing with age^1–3^. It is characterised by narrowing of the spinal canal that results in compression of the spinal cord and/or nerves, leading to symptoms such as back pain, sciatica and spinal claudication^4^. Consequently, spinal stenosis often has a significant adverse impact on affected individuals’ quality of life^5^. Although spinal stenosis can occur at any level of the spine, it most commonly affects the lumbar and cervical regions^6^. Treatment can be conservative but ever increasing rates of surgery in the USA^7–9^ and Europe^10,11^ mean it now accounts for >$15 billion^12,13^ per year in healthcare spending in the USA.

The two main, mutually non-exclusive causes of spinal stenosis are degenerative (acquired), and less commonly developmental (congenital)^1^. Degenerative spinal stenosis is thought to be caused by changes associated with aging and spinal osteoarthritis^14^. For example, in UK Biobank (UKB), a large prospective cohort in the UK, 50% of individuals with spinal stenosis diagnosis had a concurrent osteoarthrosis diagnosis.

Despite its increasing prevalence and the associated increasing healthcare cost, little is known about the epidemiology of spinal stenosis, in particular its modifiable risk factors^15^. It has been hypothesised that increased risk factor burden in the population could be responsible for this rise^16^. For example, there is observational evidence that high body mass index (BMI) predisposes to degenerative spinal disease, including spinal stenosis^17–19^. Knutsson et al. (2015)^16^ showed that obese construction workers had a twofold increased risk of lumbar spinal stenosis compared with normal weight workers. However, observational studies are liable to confounding and reverse causation making causal conclusions difficult^20^. In addition, these studies are unable to assess whether raised BMI causes spinal stenosis through degenerative changes or other pathways. Previously, it has been shown that BMI is positively correlated with bone mineral density (BMD)^21–23^ and increased BMD has been associated with osteoarthritis^24,25^. It is therefore feasible that BMD and OA are mediating the relationship between BMI and spinal stenosis. An alternative explanation is that BMI is confounding the relationship between BMD and spinal stenosis.

Mendelian randomization (MR) is an increasingly popular method for causal inference in epidemiology due to biobank-driven expansion in genome-wide association studies (GWAS) on a variety of phenotypes^26^. MR utilizes genetic variants that are randomly assigned at conception to explore causal relationships between exposures and outcomes. The technique capitalizes on the Mendelian principles of inheritance, where segregation of genetic variation is independent of confounding factors and reverse causation, so MR is particularly useful when investigating risk factors that may be challenging to examine with conventional epidemiological methods^27^. While not applied to spinal stenosis so far to the best of our knowledge, MR has previously confirmed a causal effect of BMI^25^ and BMD^25,28^ on site-specific osteoarthritis.

In this study, we employ two-sample univariable MR techniques to firstly explore the total causal relationships between genetic susceptibility to anthropometric risk factors and spinal stenosis. Among individual risk factors, we focus on measures of adiposity (BMI, waist circumference, hip circumference, waist-to-hip ratio) as overall BMI may not reflect body fat distribution and its effect on spine degeneration via mechanical and inflammatory pathways^29^. We also look at height due to potentially increased mechanical stress in tall individuals^30^, bone mineral-related traits (BMD – total and lumbar, circulating calcium and phosphorus) due to importance in maintaining bone and joint health^31,32^. Next, we employ a multivariable MR (MVMR) approach to elucidate the underlying independent mechanisms contributing to aetiology of spinal stenosis adjusting for effects of osteoarthritis and BMD.

## Materials and Methods

### Genetic association studies

We used two (**Table 1**) publicly available spinal stenosis GWAS studies in European populations available from FinnGen release 8 (https://r8.risteys.finngen.fi/phenocode/M13_SPINSTENOSIS)^33^ and UK Biobank (UKBB) available via PheWeb (https://pheweb.org/UKB-TOPMed/pheno/720)^34^ with the spinal stenosis diagnosis defined as having been assigned the ICD-10 *M48*.*0* (spinal stenosis) code. The FinnGen study was used in our main results due to increased power offered by its sample size (16,698 cases in FinnGen and 3,713 in UKBB) whereas the UKBB GWAS is used as a sensitivity analysis. The reasons for reduced prevalence seen in UKBB can be potentially attributed to misclassification due to requirement for hospital inpatient admission for ICD-10 code assignment^35^ and lower MRI diagnosis rate relative to Finland (Ville Mattila, personal communication).

Since we were interested to study the effect of genetic liability for osteoarthritis (OA) on spinal stenosis, we used the Genetics of Osteoarthritis (GO)^36^ European osteoarthritis GWAS across 3 body sites and 2 composite phenotypes (hip, knee, knee/hip – i.e. knee and/or hip, spine, all – i.e. hip, knee, hand, finger, thumb and spine) in our main analyses (**Table 1**). To avoid bias induced by sample overlap between exposure and outcome^37^ in the analysis involving UKBB spinal stenosis GWAS we used custom GO GWAS with no UKBB individuals included.

We aimed to study the direct genetic effect of a number of anthropometric risk factors on spinal stenosis (**Figure 1**): adiposity (BMI^38,39^, hip circumference^40^, waist circumference^40^, waist-to-hip ratio^40^), height^41^, bone mass density (BMD: total^42^, lumbar spine^43^ and femoral neck – this study) as well as circulating albumin-adjusted calcium (this study, **Supplementary Methods and Supplementary Table 1**) and circulating phosphate (Neale Lab GWAS available via OpenGWAS^44,45^). Again, to prevent sample overlap in a subset of MR analyses we included additional BMI^38^, femoral neck BMD^43^ GWAS with low number/no UKBB participants (but adjusted for weight).

**Figure 1.**
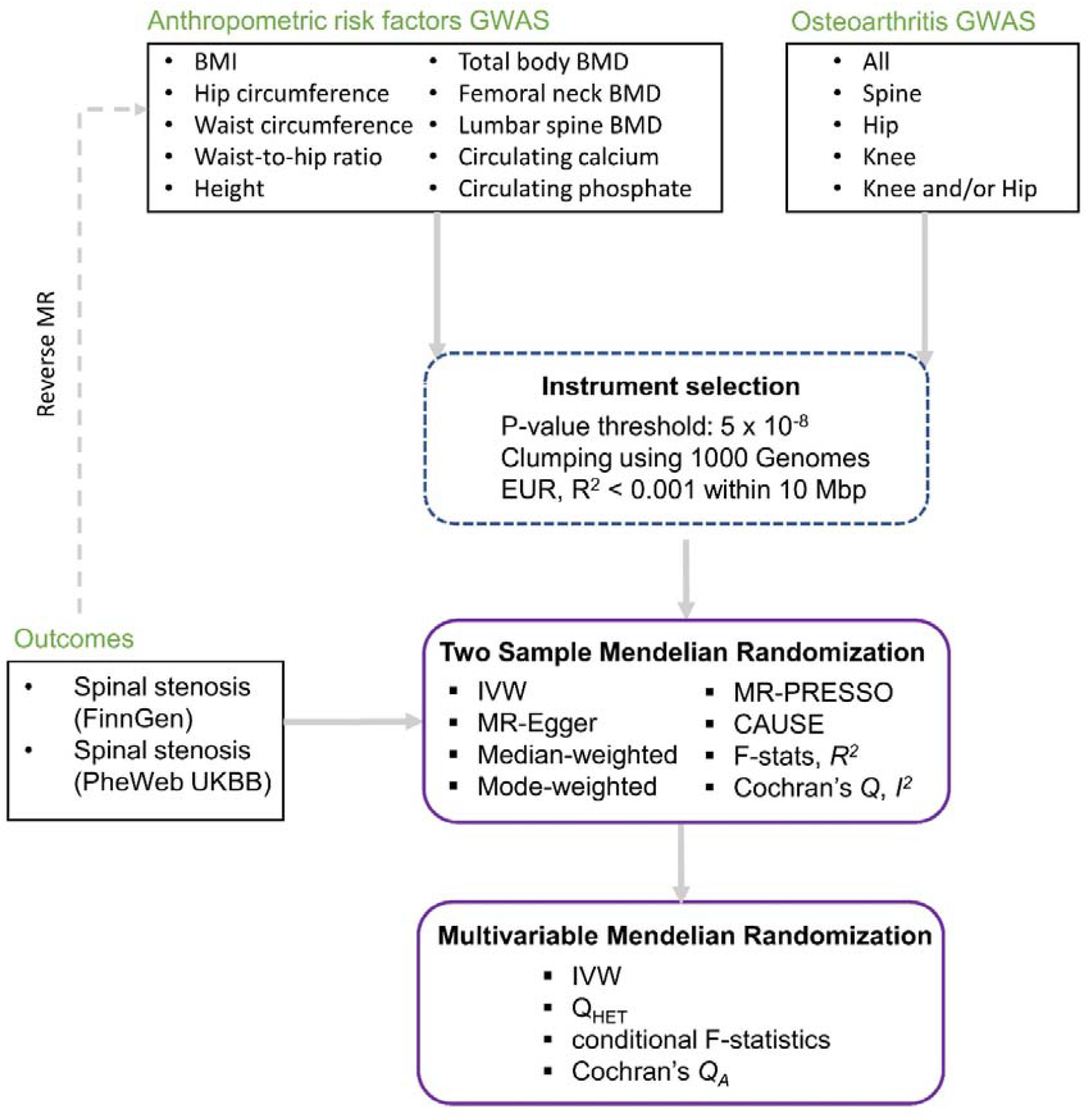
Flowchart providing overview of datasets and methods used in the current MR study.

All GWAS summary statistics files were first cast into GWAS-VCF^46^ format and then converted to TwoSampleMR package format using the *gwasvcf_to_TwoSampleMR* function from the gwasglue (https://mrcieu.github.io/gwasglue/) R package. Conversion to other formats (LDSC, MR-PRESSO, CAUSE) was carried out with custom R scripts.

### Power calculations

We used the mRnd calculator^47^ to calculate the minimum detectable odds ratio at 80% power in our main two-sample MR analyses involving spinal stenosis as the outcome.

### LD score regression

We utilized the LDSC ver 1.0.1 software^48^ to estimate the genetic correlation (rg) between osteoarthritis and spinal stenosis, using the standard procedures described in the LDSC tutorial, using HapMap 3^49^ SNPs and 1000 Genomes^50^ European ancestry reference panel to calculate LD scores.

### Selection of genetic instruments

To identify genetic instruments for each exposure, we selected SNPs that showed strong association at a genome-wide significance level (p-value < 5 × 10^-8^). We further clumped the SNPs to ensure that linkage disequilibrium (LD) as measured by r^2^ < 0.001 between any pair of significant SNPs in a 10 Mbp window in the 1000 Genomes European panel^50^ to avoid multiple instruments capturing the same causal effect. This was done using plink ver 1.9^51^ as called by ld_clump function in the ieugwasr R package (https://mrcieu.github.io/ieugwasr). In each case, genetic variant associations for the outcome trait were extracted and harmonised using default settings in the TwoSampleMR^44^ package. We next calculated the F-statistics and R^2^ to check for weak instrument bias^52^.

### Two-sample Mendelian Randomization analyses

We applied the two-sample MR approach, which utilizes summary-level data from two non-overlapping GWAS, to estimate the causal effect of anthropometric risk factors on spinal stenosis using the TwoSampleMR^44^ R package. We used the inverse-variance weighted (IVW) method as the primary analysis, where the causal estimate is obtained by combining the SNP-specific Wald ratios using a random-effects inverse-variance weighted meta-analysis. To combine the causal estimates obtained using FinnGen and UKBB spinal stenosis outcomes, we meta-analysed them with fixed-effects inverse variance method used for pooling in the R meta package^53^. Effect estimates are interpretable as change in outcome per 1 standard deviation increase in continuous exposure or per doubling in the risk of binary exposure.

### Sensitivity analyses

To assess the robustness of our findings and potential violation of MR assumptions, we performed several sensitivity analyses, including:

#### Weighted median and mode estimator^54,55^

These approaches estimate the causal effect by calculating the median and mode of the individual Wald ratios, respectively, providing a consistent estimate if at least50% of the weight comes from valid instruments (for median estimator) and the largest subset of variants identifies the same causal effect (for mode estimator).

#### MR-Egger regression^56^

This method is robust to balanced pleiotropic effects, i.e. positive and negative effects of the instrument acting through alternative pathway cancelling each other out^57^. It provides an estimate of the causal effect by regressing the SNP-outcome associations on the SNP-exposure associations, while allowing for an intercept term that captures the average pleiotropy across instruments.

#### MR-PRESSO^58^

The MR-Pleiotropy RESidual Sum and Outlier (MR-PRESSO) test was used to detect and correct for horizontal pleiotropy by identifying and removing outlier SNPs that could bias the causal effect, using the default settings.

#### CAUSE^59^

The Causal Analysis Using Summary Effect estimates (CAUSE) is a Bayesian MR method which harnesses the full genome-wide set of variant summary statistics (as opposed to only genome-wide significant SNPs in a traditional MR) to distinguish the causal effect from correlated pleiotropy (when a variant affects the exposure and outcome through a shared heritable factor) and uncorrelated horizontal pleiotropy (when a variant affects the exposure and outcome through separate mechanisms). We used it to help discern if the effect of osteoarthritis on spinal stenosis seen in standard MR analysis was driven more by shared genetic heritability of the two traits or causal effect.

#### Reverse MR

We also carried out reverse Mendelian Randomization, i.e. we used the FinnGen spinal stenosis GWAS (**Table 1**) as the exposure to detect any potential causal effect of genetic liability for spinal stenosis on any of the tested risk factors.

#### Heterogeneity

We used the standard statistics of Cochran’s *Q* and *I*^**2**^ to assess heterogeneity in our MR IVW analyses^60^.

### Multivariable Mendelian Randomization analyses

In order to test if the effect of risk factors with significant effect on spinal stenosis, as identified in the univariable analysis, is mediated by osteoarthritis, a multivariable MR model was used combining both exposure variables in a single regression test and meta-analysed using IVW method. We also carried out MVMR analyses adjusting simultaneously for BMI and body fat distribution traits^40^, as well as BMI and BMD^23^ as these are strongly positively genetically correlated.

The instrument strength (conditional F-statistics, *F*_*TS*_) and effect heterogeneity (Cochran’s *Q*_*A*_) in MVMR context were calculated using the MVMR^61^ package with the covariance between genetic associations with each exposure fixed at zero in the primary analysis, but a range of values was also tested. Since we detected presence of weak instrument bias towards the (likely) confounded observational association^52^, the Q-minimisation approach (*Q*_*HET*_) from the MVMR package^61^ was run as a sensitivity analysis to complement the MVMR-IVW results^62^.

#### MR-Strobe reporting

The study conforms to the STROBE-MR guidelines for reporting Mendelian Randomization research^63^ (**Supplementary File**).

## Results

### Investigation of total effect of risk factors on spinal stenosis using two sample MR

Our power analysis showed that we had at least 80% power for detecting small-to-moderate effects (odd ratio: 1.07-1.27) for a range of anthropometric risk factors using the FinnGen spinal stenosis GWAS (**Supplementary Table 2**). Unless otherwise stated, all the main results presented are derived using the IVW estimator and FinnGen spinal stenosis outcome. Among the adiposity traits, we found strong evidence for the effect of higher BMI (OR=1.54, 95% CI: 1.41-1.69, p-value= 2.7 x 10^-21^, **Figure 2A**), hip circumference (OR=1.50, 95% CI: 1.27-1.78, p-value=3.3 x 10^-6^, **Figure 2B**) and waist-circumference (OR=1.43, 95% CI: 1.15-1.79, p-value=1.5 x 10^-3^, **Figure 2C**) but not waist-to-hip ratio (OR=1.10, 95% CI: 0.83-1.47, p-value=0.49, **Figure 2D**) on spinal stenosis. Among the skeletal traits, we found weak evidence for a causal effect of increased height (FinnGen: OR=1.06, 95% CI: 0.99-1.14, p-value=0.10; UKBB: OR=1.15, 95% CI: 1.04-1.28, p-value=6.6 x 10^-3^; **Figure 3A**) but strong evidence for a causal effect of higher total BMD (OR=1.21, 95% CI: 1.12-1.29, p-value=1.6 x 10^-7^, **Figure 3B**), femoral neck BMD (OR=1.22, 95% CI: 1.09-1.37, p-value=5.9 x 10^-4^, **Figure 3C**) and lumbar spine BMD (OR=1.38, 95% CI: 1.25-1.52, p-value= 4.4 x 10^-11^, **Figure 3D**) on spinal stenosis. On the other hand, little evidence of an effect was found for circulating calcium (OR=1.02, 95% CI: 0.93-1.11, p-value=0.69, **Supplementary Figure 1A**) and phosphate (OR=0.94, 95% CI: 0.85-1.03, p-value=0.19, **Supplementary Figure 1B**).

**Figure 2.**
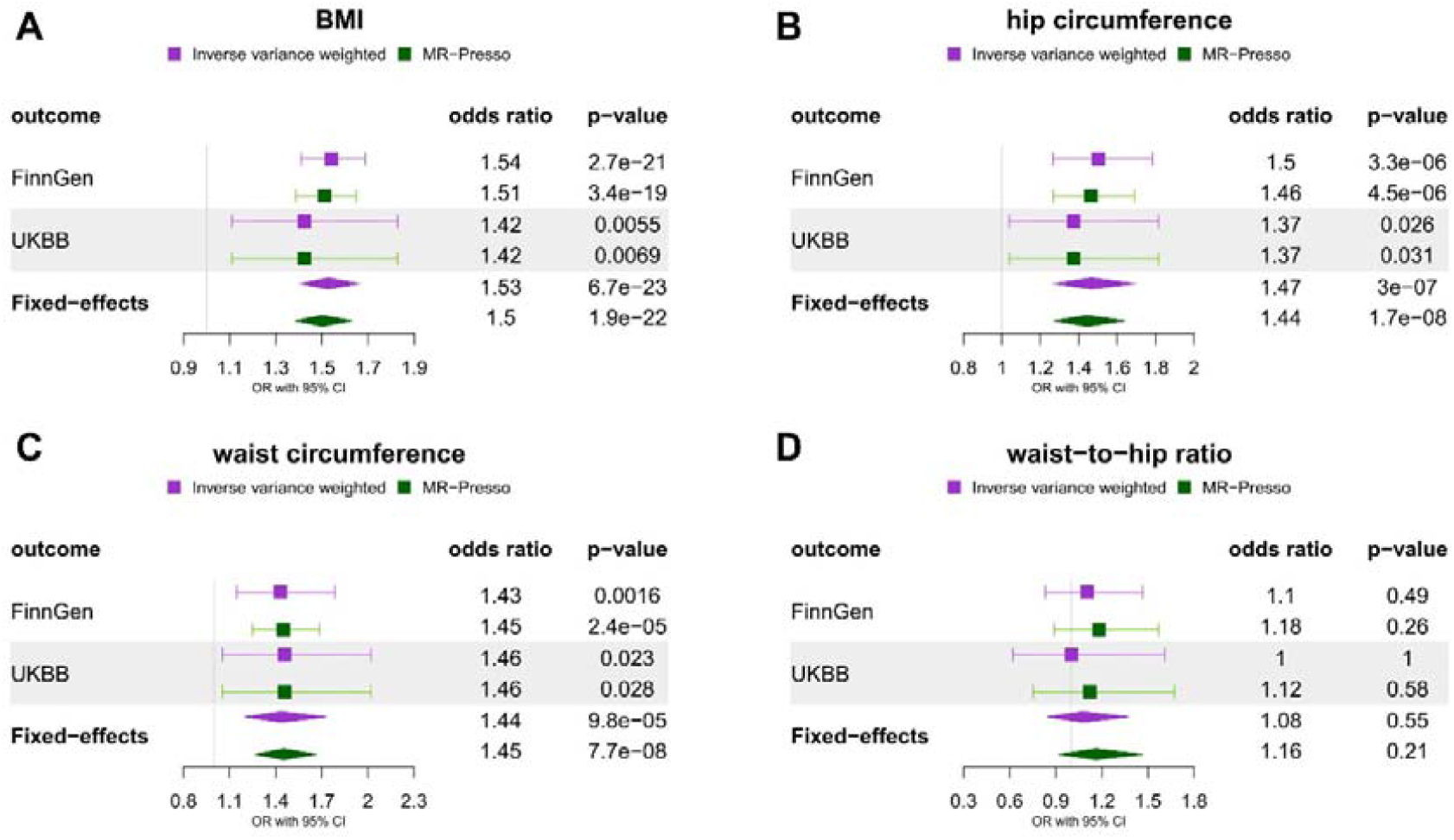
Two sample Mendelian randomization results for the effect of genetic susceptibility for adiposity traits (A – BMI, B – hip circumference, C - waist circumference, D – waist-to-hip ratio) on spinal stenosis (FinnGen and UK BioBank). Plots compare results obtained using IVW and outlier-robust MR-PRESSO method and display fixed-effects meta-analysis results of the odds ratio per SD increase in exposure obtained using FinnGen and UK Biobank outcomes.

**Figure 3.**
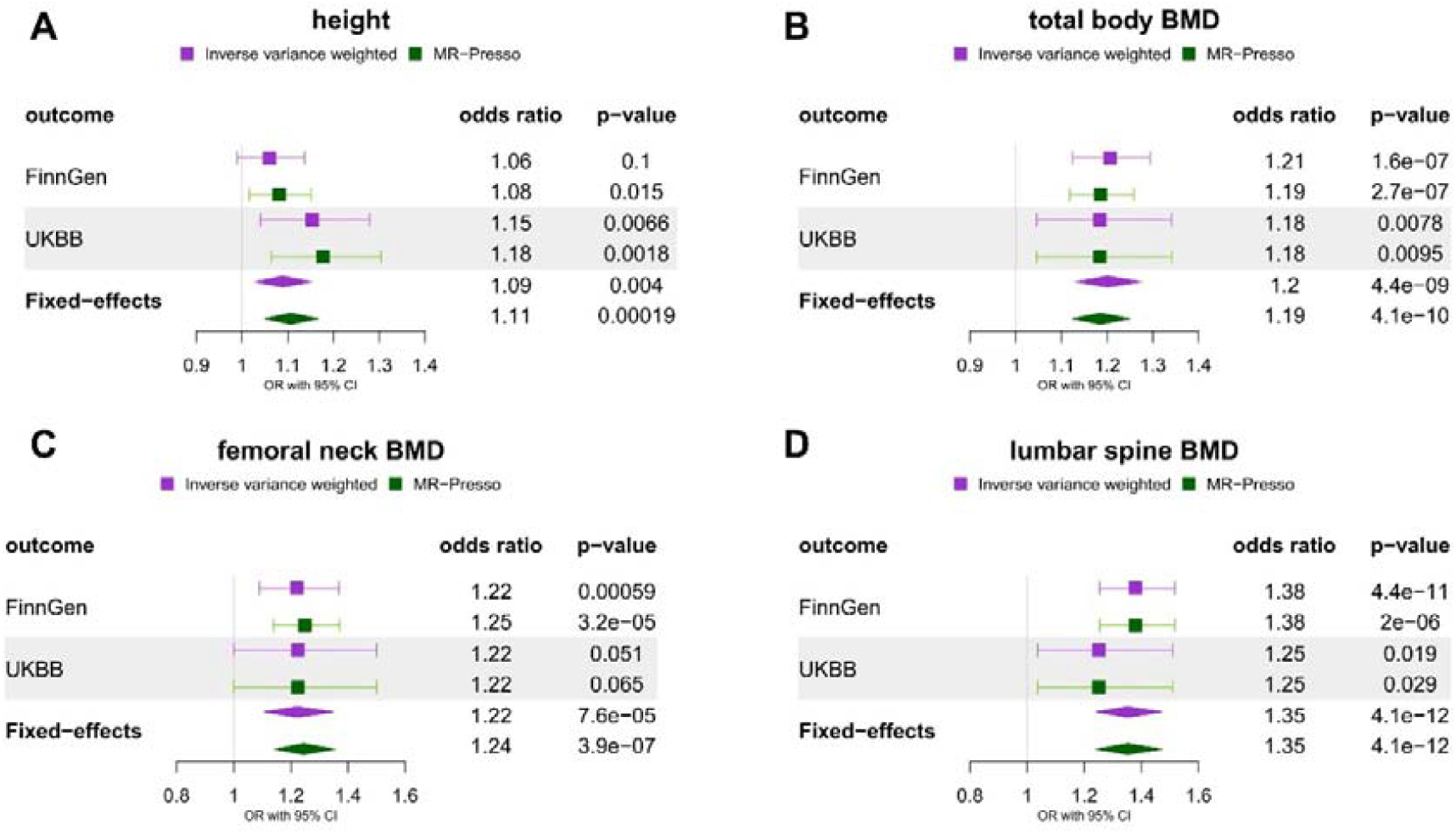
Two sample Mendelian randomization results for the effect of genetic susceptibility for skeletal traits (A – height, B – total body BMD, C – femoral neck BMD, D – lumbar spine BMD) on spinal stenosis (FinnGen and UK Biobank). Plots compare results obtained using IVW and outlier-robust MR-PRESSO method and display fixed-effects meta-analysis results of the odds ratio per SD increase in exposure obtained using FinnGen and UK Biobank outcomes. *BMD* – bone mineral density

### Sensitivity analyses – two sample MR

Fixed-effects meta-analysis of MR results based on both FinnGen and UK Biobank spinal stenosis outcome GWAS resulted in similar estimates to those obtained using solely FinnGen (**Figure 2-3**). SNP outliers apparent in the scatter plots (**Supplementary Figure 9-18**) along with significant Cochran’s **Q** values for all exposures (except for lumbar spine BMD, **Supplementary Table 5**) suggested presence of effect heterogeneity. However, outlier-robust sensitivity method MR-PRESSO (**Figure 2-3**) reproduced the same magnitude of associations, while other methods (MR Egger, weighted median and mode) were consistent with IVW/MR-PRESSO estimates overall (**Supplementary Table 3**). Non-significant Egger’s intercept (**Supplementary Table 7**) suggested limited presence of horizontal pleiotropy. Reverse MR analysis (**Supplementary Table 3**) with the FinnGen spinal stenosis found little evidence of effect on all risk factor traits, apart from lumbar spine BMD (OR=1.15, 95% CI: 1.06-1.26, p-value=1.5 x 10^-3^).

### Shared genetic liability for spinal stenosis and osteoarthritis

We then investigated the magnitude of LD score-derived genetic correlation between spinal stenosis and osteoarthritis across various sites (**Figure 4**). As expected, the highest correlation was found between the two spinal stenosis GWAS (rg = 0.77, p-value=1.1 x 10^-23^), however high genetic correlation was also revealed between spine osteoarthritis (rg_FinnGen_=0.66, p-value=4 x 10^-22^; rg_UKBB_=0.73, p-value=1.3 x 10^-11^) and spinal stenosis. Genetic correlation across other osteoarthritis sites was high in the FinnGen spinal stenosis GWAS (from rg=0.47, p-value=2.5 x 10^-23^ and p-value= 2.8 x 10^-17^ for knee and hip OA, respectively, to rg=0.52, p-value=2.4 x 10^-25^ for all OA) and moderate in the UKBB spinal stenosis GWAS (rg ranging 0.3-0.38).

**Figure 4.**
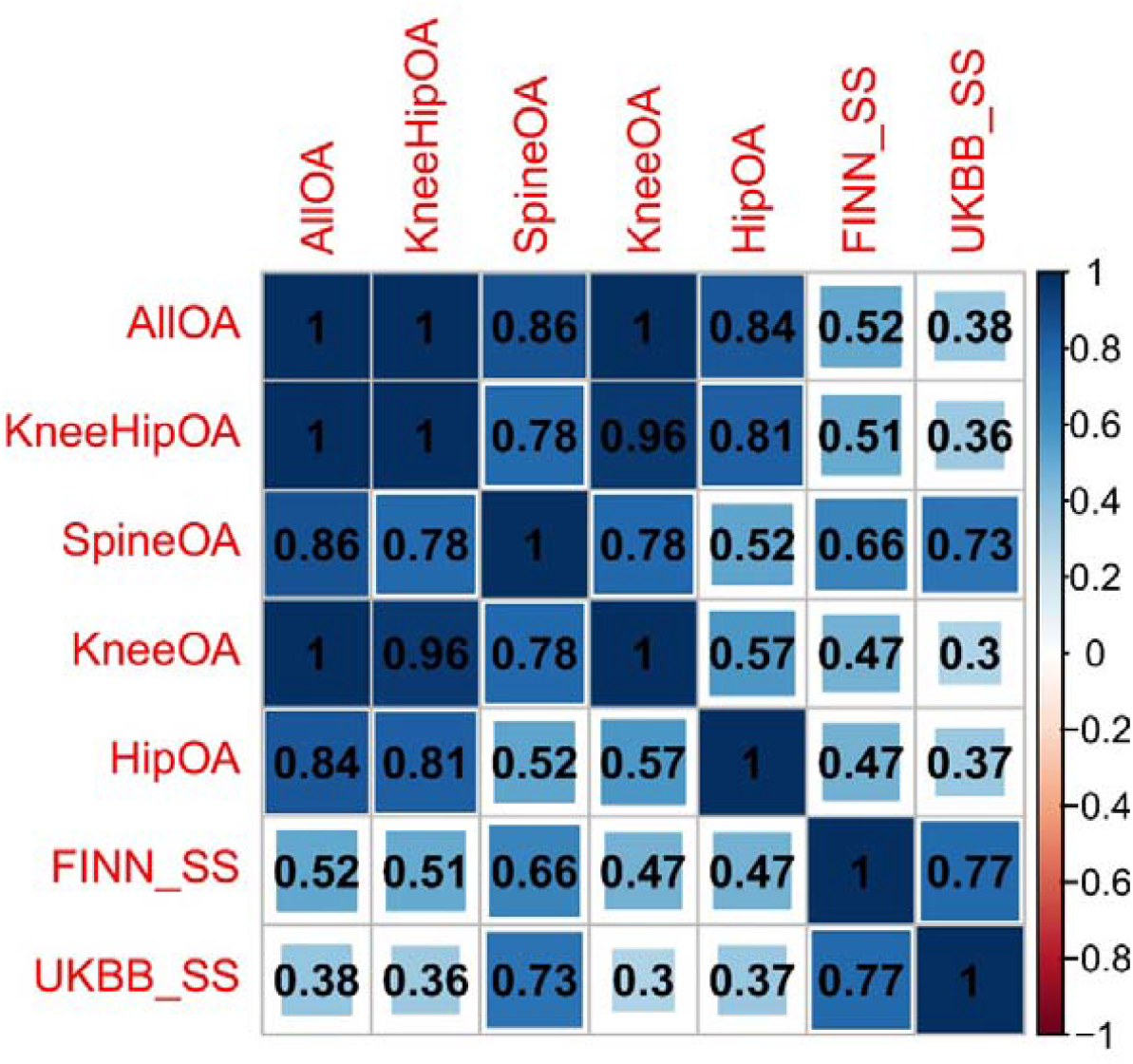
Genetic correlation of osteoarthritis (OA) and spinal stenosis phenotypes estimated by LD score regression. Correlation coefficients are displayed within cells and the colour/area of the cells are proportionally scaled. All p-values are significant after FDR correction. *FINN_SS* – FinnGen spinal stenosis, *UKBB_SS* – UK Biobank spinal stenosis.

### Shared risk factors for osteoarthritis and spinal stenosis

Given the substantial genetic correlation of spinal stenosis with osteoarthritis and evidence of causal effects of adiposity traits as well as BMD on osteoarthritis in previous MR studies^25,28,64,65^, we hypothesised that osteoarthritis may be a major mediator of the effects of these risk factors on spinal stenosis (**Figure 5**). First, to investigate this hypothesis in the two-step MR framework^66^, we replicated the evidence for causal effects of anthropometric risk factors on osteoarthritis (**Supplementary Table 4, 6, 8**): BMI (**Supplementary Figure 2**), hip circumference (**Supplementary Figure 3**), waist circumference (**Supplementary Figure 4**), height (**Supplementary Figure 5**), total BMD (**Supplementary Figure 6**), femoral neck BMD (**Supplementary Figure 7**) and lumbar spine BMD (**Supplementary Figure 8**). As the next step, we assessed the bidirectional relationship between osteoarthritis and spinal stenosis (**Supplementary Figure 19-22**). The main IVW result confirmed the causal effect of all site osteoarthritis on spinal stenosis (**Figure 6, Supplementary Figure 23, Supplementary Table 3**) with odds ratio of 1.44 (95% CI: 1.13-1.84, p-value=3.1 x 10^-3^). IVW result for knee osteoarthritis (OR=1.16, 95% CI: 0.98-1.38, p-value=0.09) was markedly increased after outlier correction using MR-PRESSO (OR=1.34, 95% CI: 1.18-1.52, p-value=1.3 x 10^-4^) and a very uncertain estimate was available for spine osteoarthritis (OR=1.13, 95% CI: 0.75-1.71, p-value=0.56) as calculated using a single instrument (F-statistic=30, **Table 1**).

**Figure 5.**
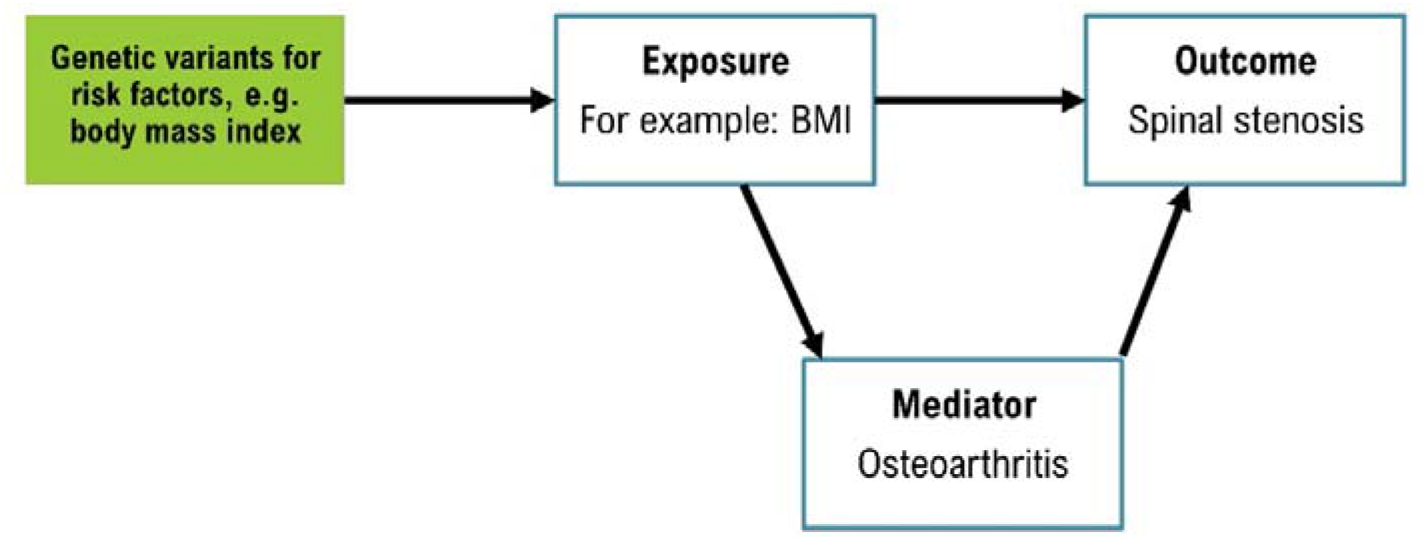
A directed acyclic graph (DAG) showing the tested associations between anthropometric risk factors (e.g. body mass index), osteoarthritis and spinal stenosis.

**Figure 6.**
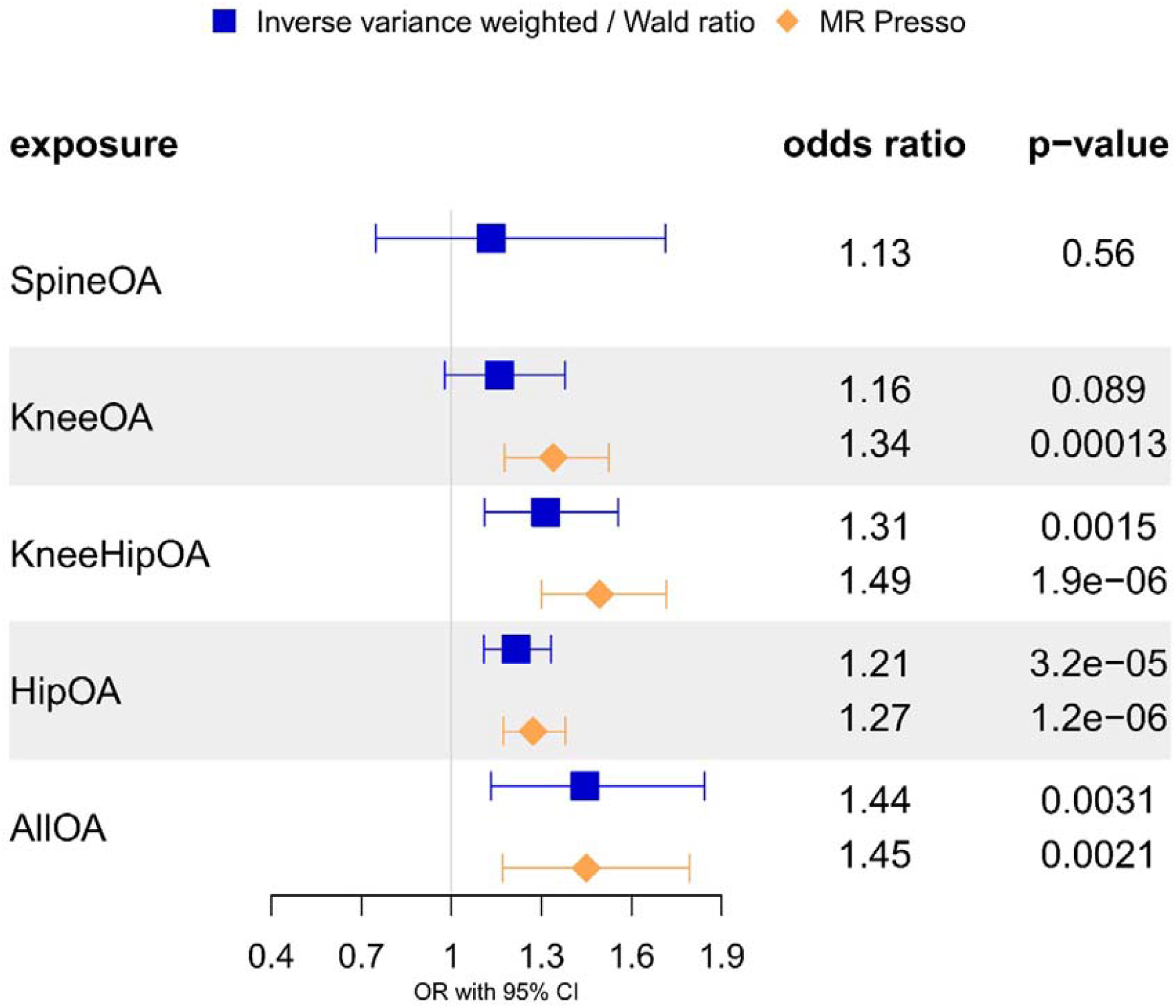
Two sample Mendelian randomization results for the effect of genetic susceptibility for osteoarthritis on spinal stenosis (FinnGen). Plot compares results obtained using IVW/Wald ratio and outlier-robust MR-PRESSO method. The odds ratios displayed are scaled per doubling in odds of exposure.

### Spinal stenosis is causally downstream of osteoarthritis

To help overcome this power limitation and establish the true causal path between osteoarthritis and spinal stenosis given their shared genetic heritability, we applied the Bayesian CAUSE method (**Supplementary Table 9**). When evaluating the bidirectional relationship between osteoarthritis and FinnGen spinal stenosis GWAS, the causal model was always picked over the sharing model (p-value from 1.4 x 10^-6^ to 4.9 x 10^-3^). In each case, effect size for the osteoarthritis to spinal stenosis direction dominated (OR_all OA_=1.6, 95% CI: 1.41-1.79; OR_spinal OA_=1.4, 95% CI: 1.21-1.62) over the reverse direction (OR_all OA_=1.07, 95% CI: 1.05-1.09; OR_spinal OA_=1.13, 95% CI: 1.09-1.17).

### Investigation of direct effect of risk factors on spinal stenosis independent of osteoarthritis using multivariable MR

In light of the predicted strong causal effect of osteoarthritis on spinal stenosis and both osteoarthritis and spinal stenosis sharing the same set of anthropometric risk factors in our two-sample MR analyses, we employed multivariable MR to estimate the direct effect of a given risk factor on spinal stenosis accounting for OA (**Supplementary Table 10**). The direct effect of higher BMI on spinal stenosis (**Figure 7A, Supplementary Figure 24-25**) ranged from OR=1.29 for all osteoarthritis mediator (95% CI: 1.16-1.45, p-value=7.2 x 10^-6^) to OR=1.37 for spine osteoarthritis mediator (95% CI: 1.24-1.51, p-value=4.7 x 10^-10^) which corresponded to all OA mediating 16.2% (95% CI: 14.2%-17.8%) of the total effect of BMI on spinal stenosis. For height, adjusting for OA resulted in the direct effect being consistent with the null hypothesis (**Figure 7B, Supplementary Figure 26**) for all OA (OR=1.01, 95% CI: 0.94-1.08, p-value=0.79) and spine OA (OR=1.01, 95% CI:0.94-1.08, p-value=0.85), albeit a weak direct effect remained in the UKBB analysis (**Supplementary Figure 27**). Next, total body BMD direct effect adjusted for OA (**Figure 7C, Supplementary Figure 28-29)** resulted in OR=1.19 (95% CI: 1.11-1.29, p-value=6.6 x 10^-6^) for all OA and in OR=1.2 (95% CI: 1.11-1.29, p-value=1 x 10^-6^) for spine OA. Interestingly, unadjusted odds-ratio for total body BMD did not meaningfully differ (OR=1.21, 95% CI: 1.12-1.29, p-value=1.6 x 10^-7^) suggesting total body BMD affects osteoarthritis through an independent pathway. This was not unlike femoral neck BMD (**Figure 7D, Supplementary Figure 30-31**), where the direct effect accounting for all OA (OR=1.19, 95% CI: 1.06-1.33, p-value=3.2 x 10^-3^) and spine OA (OR=1.15, 95% CI: 1.01-1.30, p-value=0.03) equated to all OA mediating 2.5% of the total effect of femoral neck BMD on spinal stenosis. Similarly, relatively low (5.8%) degree of mediation was found for the lumbar spine BMD outcome (**Supplementary Figure 32-33**).

**Figure 7.**
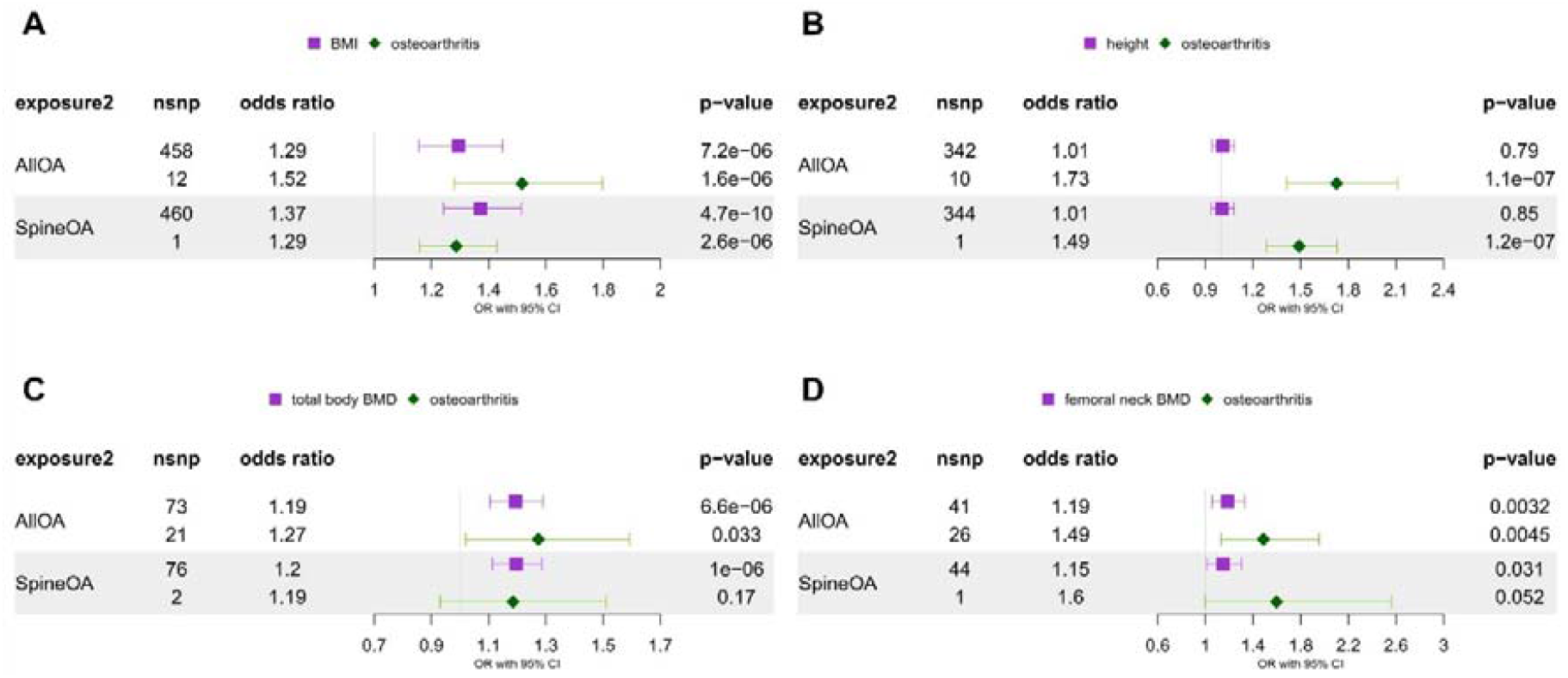
Multivariable Mendelian randomization results for the jointly modelled effect of genetic susceptibility for risk factors (A – BMI, B – height, C – total body BMD, D – femoral neck BMD) and liability for osteoarthritis (all or spine) on spinal stenosis (FinnGen). The odds ratios are scaled per SD increase of risk factors and doubling in the odds of osteoarthritis.

### Direct effect of waist/hip circumference on spinal stenosis independent of BMI

Since the two non-BMI adiposity risk factors which we identified (waist and hip circumference) are phenotypically and genetically correlated with BMI, we used the MVMR approach to arrive at direct estimates adjusted for BMI (**Figure 8, Supplementary Figure 34**). We found that the corrected estimates shifted towards the null for both waist (OR=1.13, 95% CI=0.82-1.55, p-value=0.45) and hip circumference (OR=1.12, 95% CI=0.85-1.46, p-value=0.42).

**Figure 8.**
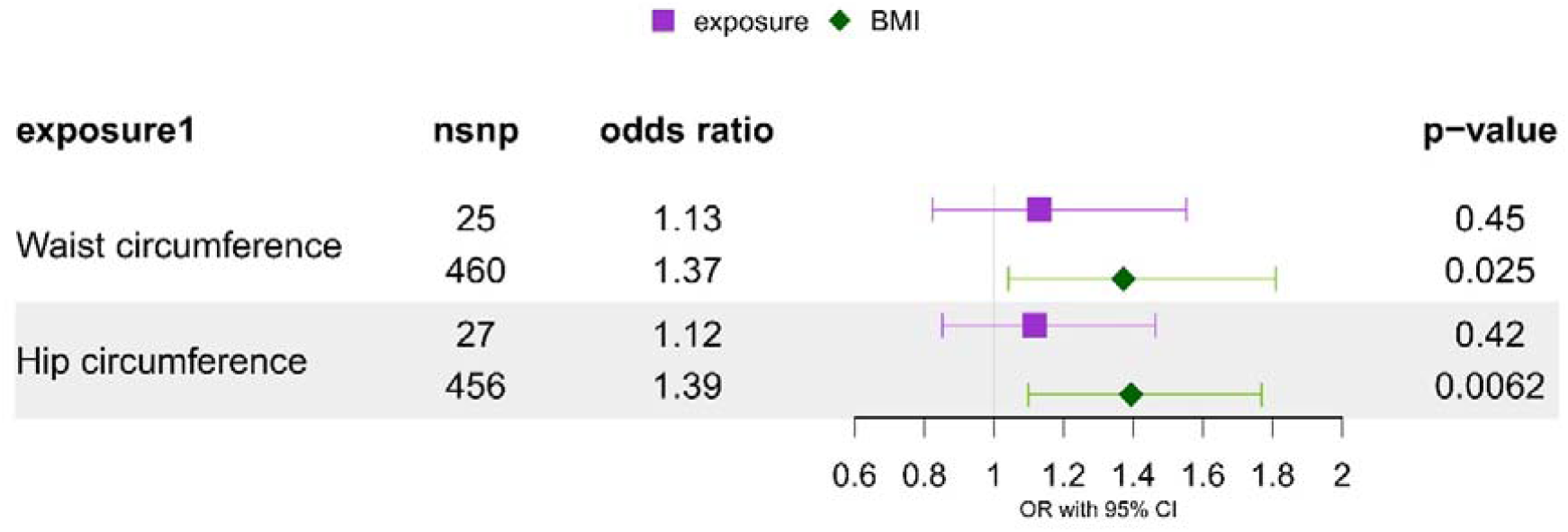
Multivariable Mendelian randomization results for the jointly modelled effect of genetic susceptibility for risk factors (waist and hip circumference) and body mass index on spinal stenosis (FinnGen). The odds ratios are scaled per SD increase of risk factors.

### Direct effect of BMD on spinal stenosis independent of both osteoarthritis and BMI

Lastly, since previous research hypothesised that BMI can be a confounder of a relationship between BMD and osteoarthritis^28^, we were interested in studying the mutually adjusted effect of the three variables on spinal stenosis (**Figure 9, Supplementary Table 11**). In the model including total body BMD and all OA exposures, the estimated effect of BMI on spinal stenosis remained consistent (OR=1.32, 95% CI: 1.16-1.47, p-value=3.2 x 10^-6^) with the model including only OA covariate, while the total BMD estimate was slightly attenuated (OR=1.13, 95% CI: 1.03-1.24, p-value=8.3 x 10^-3^) but there is a large amount of uncertainty in the estimate. The results for the model involving spine OA rather than all OA, and femoral neck BMD rather than total body BMD (**Supplementary Figure 35-38**) were analogous. There is some evidence that BMI is common cause of both lumbar spine BMD and OA (**Supplementary Figure 39-40**) shown by the significant reduction in the effect of lumbar spine BMD on spinal stenosis (adjusted for both BMI and all OA: OR=1.01, 95% CI: 0.90-1.14, p-value=0.83; adjusted for all OA only: OR=1.30, 95% CI: 1.15-1.47, p-value=4.4 x 10^-5^).

**Figure 9.**
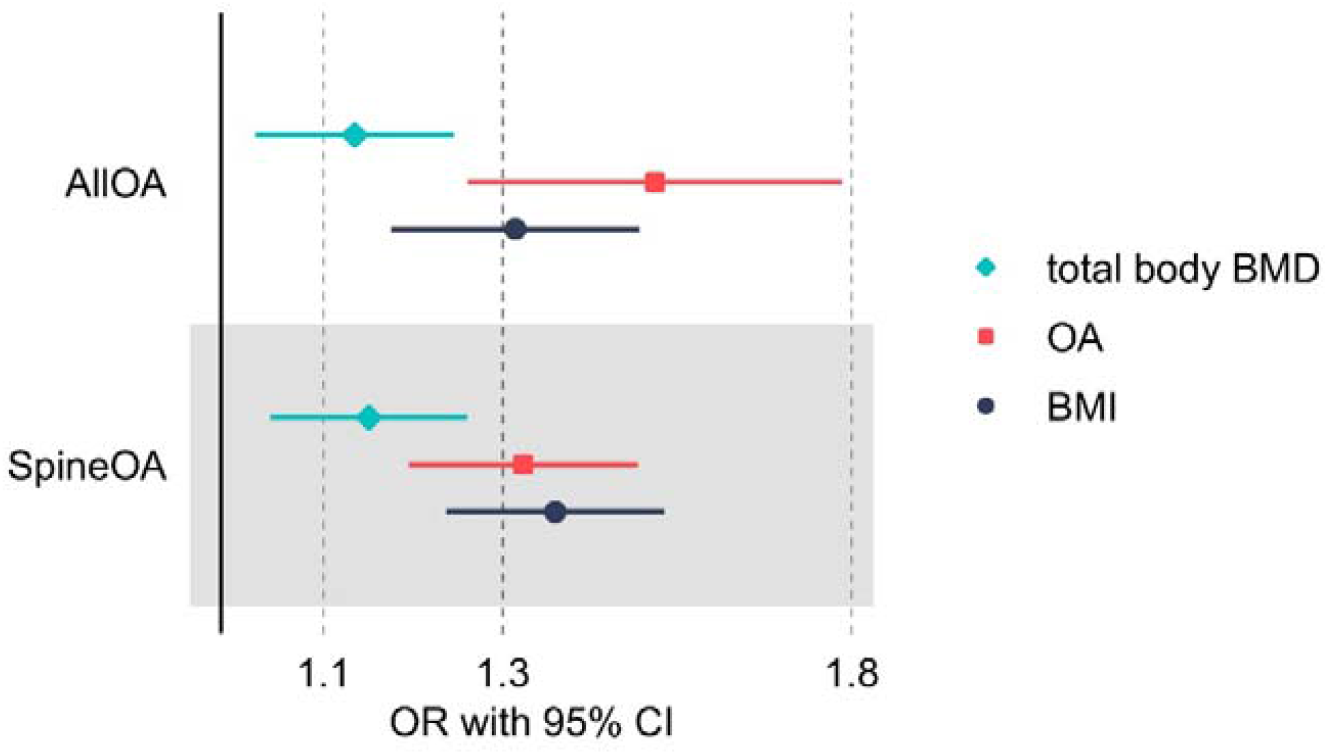
Multivariable Mendelian randomization results for the jointly modelled effect of genetic susceptibility for total body BMD, BMI and liability for osteoarthritis (all or spine) on spinal stenosis (FinnGen). The odds ratios are scaled per SD increase of risk factors and doubling in the odds of osteoarthritis.

### Sensitivity analyses – multivariable MR

As we detected presence of potential pleiotropy due to high heterogeneity as measured by Cochran’s **Q**_**A**,_ and weak instrument bias evidenced by conditional F-statistics < 10 in our MVMR analyses (**Supplementary Table 12, 13**), we applied the robust estimator Q_HET_ in a sensitivity analyses. The method produced results generally consistent with the IVW MVMR results, albeit with a much higher degree of uncertainty around the true causal value (**Supplementary Tables 14-15**).

## Discussion

Our understanding of spinal stenosis epidemiology remains quite limited despite the condition’s relatively high prevalence among older adults and its association with substantial pain and mobility impairment. In this study, we applied a genetic epidemiology method (MR) to investigate the causal relationships between anthropometric risk factors, osteoarthritis and spinal stenosis.

When analysed independently, BMI was found to act as a strong risk factor for spinal stenosis (meta-analysed OR=1.53 per 1 SD increase in exposure), similar to hip circumference (OR=1.47) and waist circumference (OR=1.44) but these attenuated to the null after adjusting for BMI in multivariable analysis. BMD across different sites also showed a substantial effect on spinal stenosis: total (OR=1.2), hip (femoral neck, OR=1.22) and lumbar spine (OR=1.35). As lumbar spine BMD measurement is liable to falsely increase with degenerative change^67–69^ and spinal stenosis liability affects lumbar spine BMD in our reverse MR analysis, we subsequently focussed on total and hip BMD. Interestingly, in a previous case-control study higher BMD was found in lumbar spinal stenosis cases across not only the lumbar spine, but also femoral neck and total hip^70^. In addition, we found that circulating calcium and phosphate exhibited little to no evidence for an effect on spinal stenosis.

Osteoarthritis, in particular facet joint osteoarthritis of the spine, can contribute to the narrowing of the spinal canal thanks to joint hypertrophy and formation of synovial cysts^15^. In agreement with this biological mechanism, our MR analysis found a positive effect of a genetic predisposition to osteoarthritis (when measured at all sites) on the development of spinal stenosis. These results were further supported by the Bayesian CAUSE model which found our results were more likely to be driven by a causal effect of a genetic predisposition to OA than by correlated and horizontal pleiotropy. We also identified a reverse causal effect, hypothesized to be indicative mostly of a shared genetic aetiology, as supported by LD score regression. It is worth noting that while our spinal OA signals showed consistent results, in terms of direction of effect, the estimates were less preciselikely due to the reduced number of genetic instruments as compared with OA at all sites.

MVMR identified a largely OA-independent causal pathway between BMI, BMD and spinal stenosis, with OA mediating < 20% of the effect of BMI and <6% of BMD. However, weak evidence for the causal effect of height on spinal stenosis (OR=1.09) was diminished to the null in the MVMR analyses suggesting that the univariable effect was driven by the causal association with osteoarthritis. Moreover, we did not find compelling evidence for BMI to be acting as a confounder for the association of BMD, OA and spinal stenosis.

MR can only provide reliable causal estimates subject to meeting three key assumptions which were tested in multiple ways in our analysis. The first criterion (“relevance”), that the genetic variants are robustly associated with the risk factor of interest was met by using variants with genome-wide significant associations with exposure and using variants with F-statistics >30 that should minimize weak instrument bias, which can arise when the genetic variant explains only a small proportion of the variance in the risk factor^71^. The second criterion, that the genetic variant shares no unmeasured confounder with the outcome (“independence” / “exchangeability”) is usually concerned with confounding by population stratification^63^ which is addressed during the initial GWAS analysis. In addition, bidirectional MR analysis confirmed that associations between risk factors and spinal stenosis were not confounded by reverse causation in all but one case.

Perhaps the most pervasive problem plaguing MR analysis is the violation of the third assumption, that the genetic variant affects the outcome only through its association with the risk factor, and not through any other independent pathways (“exclusion restriction”, i.e. no horizontal pleiotropy). We evaluated this assumption with the MR Egger intercept test and MR-PRESSO analysis. Also included were a range of MR sensitivity methods (MR-Egger, weighted median, weighted mode) whose results are consistent in magnitude with the main IVW results and so indicate that the independence and exclusion restriction assumptions were not violated.

Our IVW MVMR analysis typically suffered from low strength of the genetic instrument for 1-2 exposures. We tried to rectify that by applying the Q-minimisation approach which is more robust to these violations of MR assumptions but there remains a possibility that our MVMR direct estimates are incompletely adjusted.

Since there was no gold standard diagnostic tool for spinal stenosis at the time of data collection with diagnosis based on clinical history, physical examination, and imaging^72,73^, varying case definition will introduce an additional layer of heterogeneity into GWAS and reduce its power. Using a severe end of the phenotype spectrum can lead to reduced power in GWAS, and so fewer genomewide-significant hits. This is demonstrated by 0 versus 21 genome-wide significant loci in the UKBB (3,713 cases) and FinnGen (16,698 cases) spinal stenosis GWAS, respectively. Likewise, the OA outcomes from the GO consortium included a range of definitions, including hospital diagnosis, radiographic evidence and self-reporting, which can inflate estimate heterogeneity, and so increase the risk of a weak instrument bias. Furthermore, this MR study could benefit from inclusion of more ancestrally diverse populations to compare the estimated effects of identified risk factors but currently no suitable spinal stenosis outcome GWAS in non-Europeans is available.

Our study has public health implications, as efforts to minimise prevalence of high adiposity in the population should lead to reduction in spinal stenosis incidence and associated benefits regarding quality of life and healthcare costs. Previously identified obese individuals with elevated BMD measurement could be especially targeted for weight loss intervention due to higher compounded risk of spinal stenosis. Moreover, while the current MR study uses condition prevalence as the outcome, it is quite likely that the risk factors identified could contribute to progression of symptoms.

## Conclusions

In this study, we examined a variety of potential anthropometric risk factors for spinal stenosis, both independently and in conjunction with potential mediators. Our findings, confirmed by two-sample IVW MR, MR-PRESSO, and CAUSE analyses, demonstrate that a genetic predisposition to osteoarthritis causally contributes to the development of spinal stenosis. Overall, we have found evidence for osteoarthritis-independent causal effect of BMI on spinal stenosis, in addition to BMI- and osteoarthritis-independent causal effect of BMD. Further investigation is necessary to elucidate the mechanisms through which elevated BMD and BMI contribute to spinal stenosis, as well as to explore the functional genomics of spinal stenosis, including potential drug targets.

## Supporting information

Main Tables

## Data Availability

All data produced in the present work are contained in the manuscript.

## Abbreviations

BMD: bone mineral density
BMI: body mass index
CI: confidence interval
OA: osteoarthritis
OR: odds ratio

## Data availability

GWAS summary statistics availability is provided by relevant publications. Custom summary statistics were obtained from the GO Consortium on application.

## Funding

This work was funded by the UK Medical Research Council (MRC) as part of the MRC Integrative Epidemiology Unit (MC_UU_00032/03). This study was supported by the National Institute for Health and Care Bristol Biomedical Research Centre. The views expressed are those of the authors and not necessarily those of the NIHR or the Department of Health and Social Care. BGF is funded by an NIHR Academic Clinical Lectureship. MF is funded by a Wellcome Trust collaborative award (reference number 209233).

## Competing interests

T.R.G. receives funding from Biogen for unrelated research.

## Author contributions

MKS: Conceptualization, Formal analysis, Visualization, Writing – original draft, Writing – review and editing

BGF: Writing – review and editing

GO consortium, MF, AH, LS, EZ, HT: Resources, Contributions to the acquisition of data, Critical revision of the manuscript and final approval of the version to be published

TG: Supervision, Writing – review and editing, Funding acquisition

## Acknowledgements

This research has been conducted using the UK Biobank Resource (application number 17295).

## Ethics approval

UK Biobank received ethical approval from the Research Ethics Committee (REC reference: 11/NW/0382). As this study did not involve human subjects or individual-level data, no ethics approval was required.

