## Supplementary Figures for "Causal relationships between anthropometric traits, bone mineral density, osteoarthritis and spinal stenosis: a Mendelian randomization investigation"

**Supplementary Figure 1.** Two sample Mendelian randomization results for the effect of genetic susceptibility for circulating calcium (A) and phosphate (B) on spinal stenosis (FinnGen). The odds ratios are scaled per SD increase in exposure.

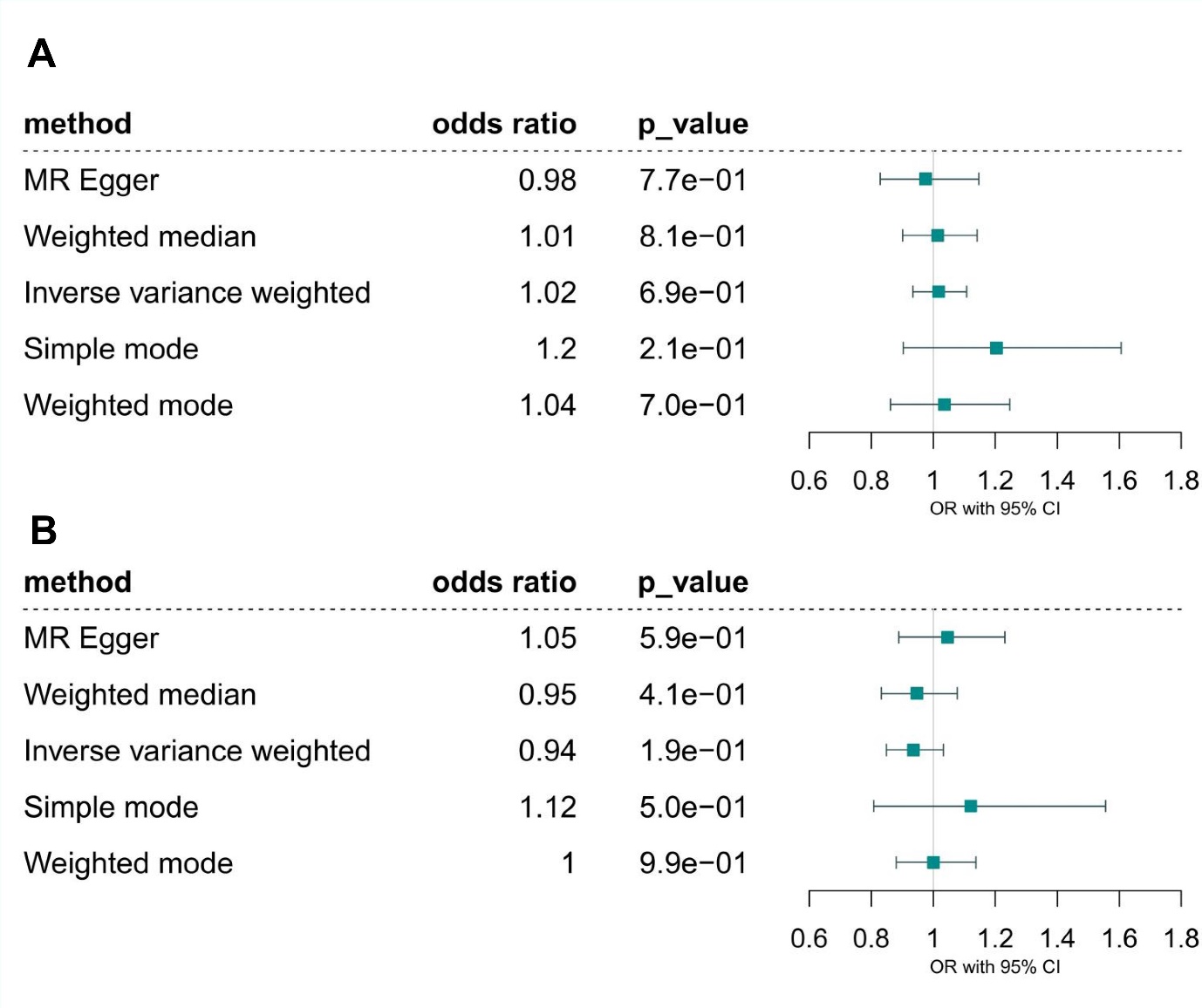

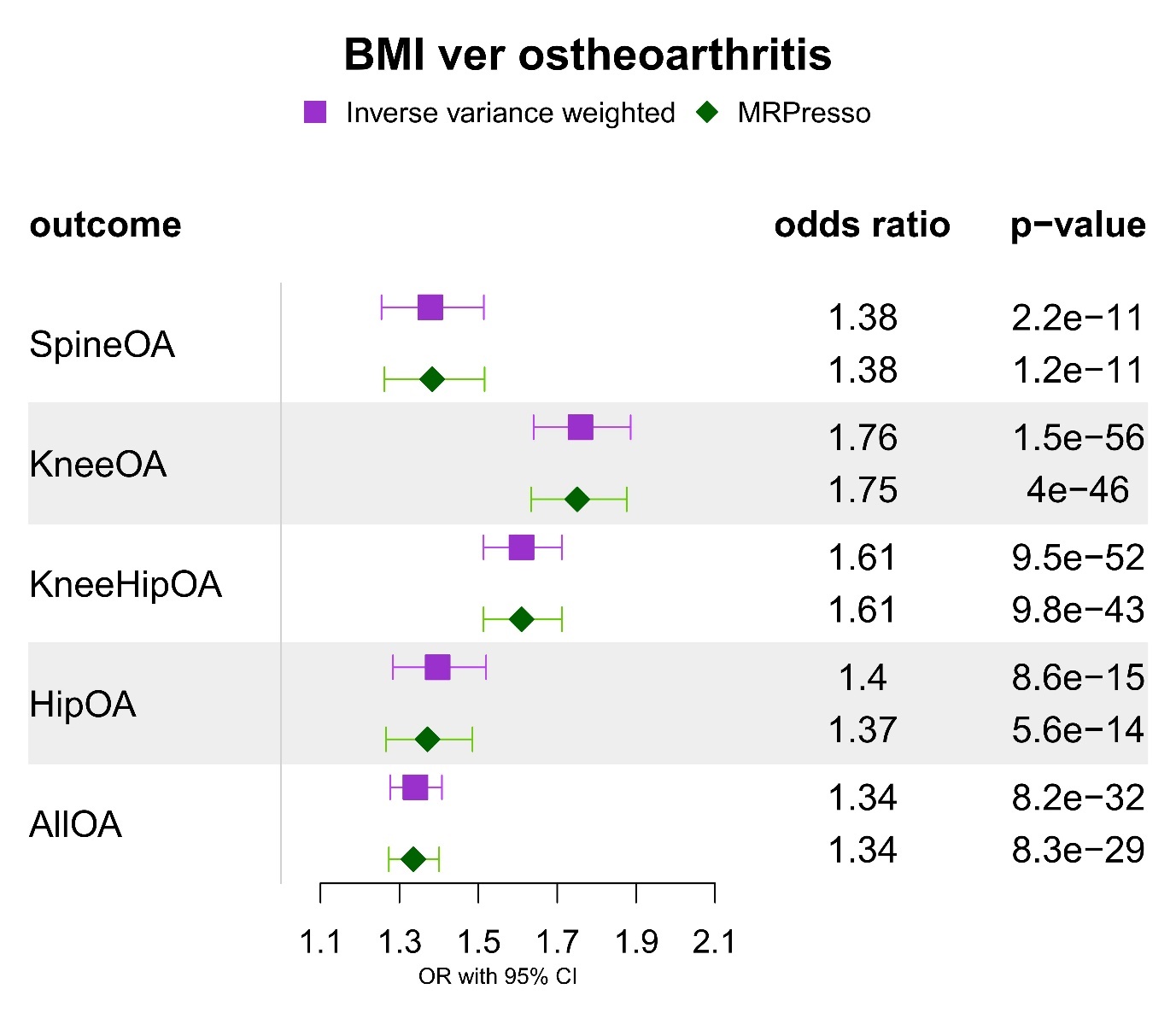
**Supplementary Figure 2.** Two sample Mendelian randomization results for the effect of genetic susceptibility for body mass index on site-specific and all osteoarthritis. The odds ratios are scaled per SD increase in exposure.

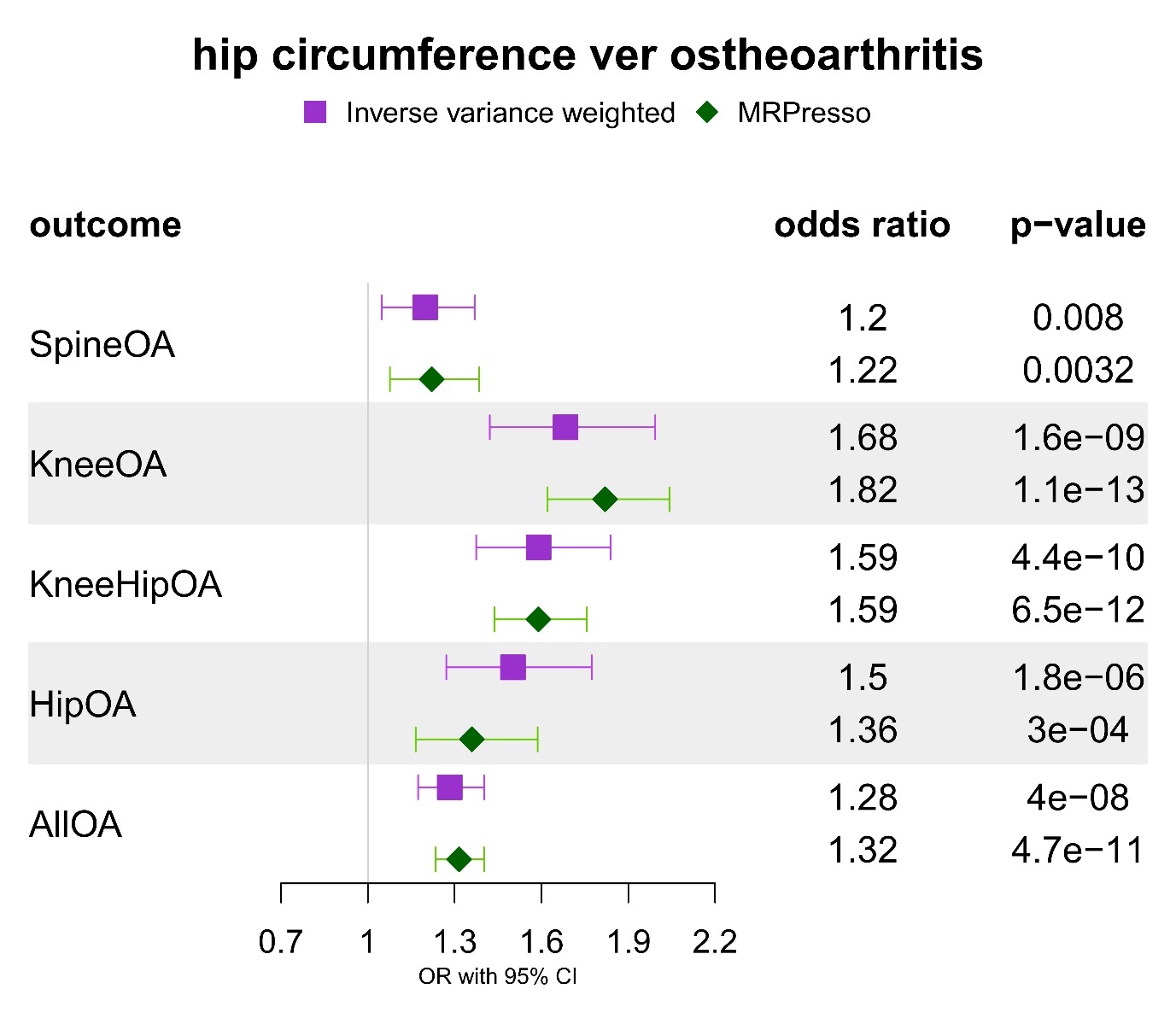
**Supplementary Figure 3.** Two sample Mendelian randomization results for the effect of genetic susceptibility for hip circumference on site-specific and all osteoarthritis. The odds ratios are scaled per SD increase in exposure.

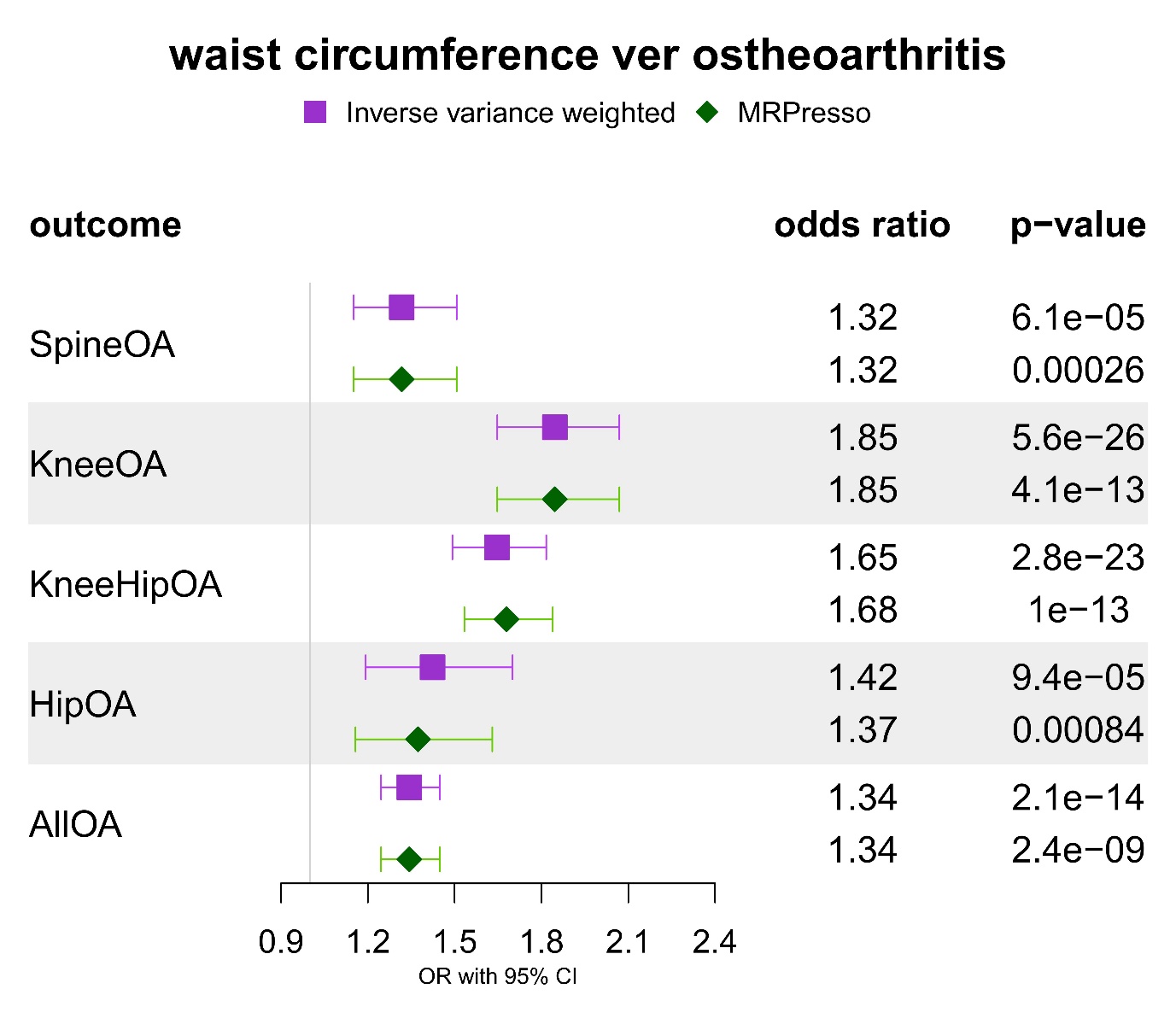
**Supplementary Figure 4.** Two sample Mendelian randomization results for the effect of genetic susceptibility for waist circumference on site-specific and all osteoarthritis. The odds ratios are scaled per SD increase in exposure.

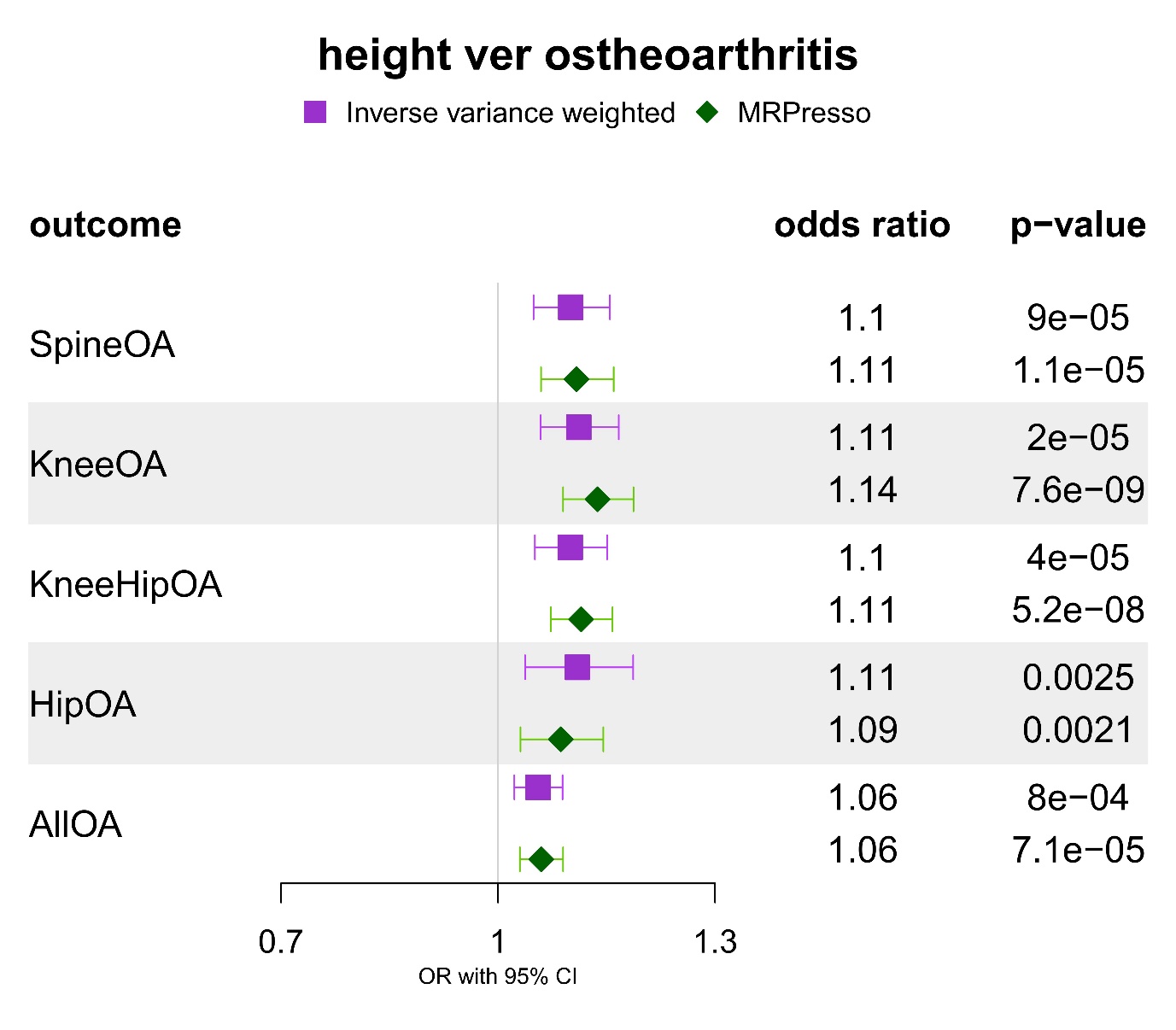
**Supplementary Figure 5.** Two sample Mendelian randomization results for the effect of genetic susceptibility for height on site-specific and all osteoarthritis. The odds ratios are scaled per SD increase in exposure.

**Supplementary Figure 6.** Two sample Mendelian randomization results for the effect of genetic susceptibility for total body bone mineral density on site-specific and all osteoarthritis. The odds ratios are scaled per SD increase in exposure.

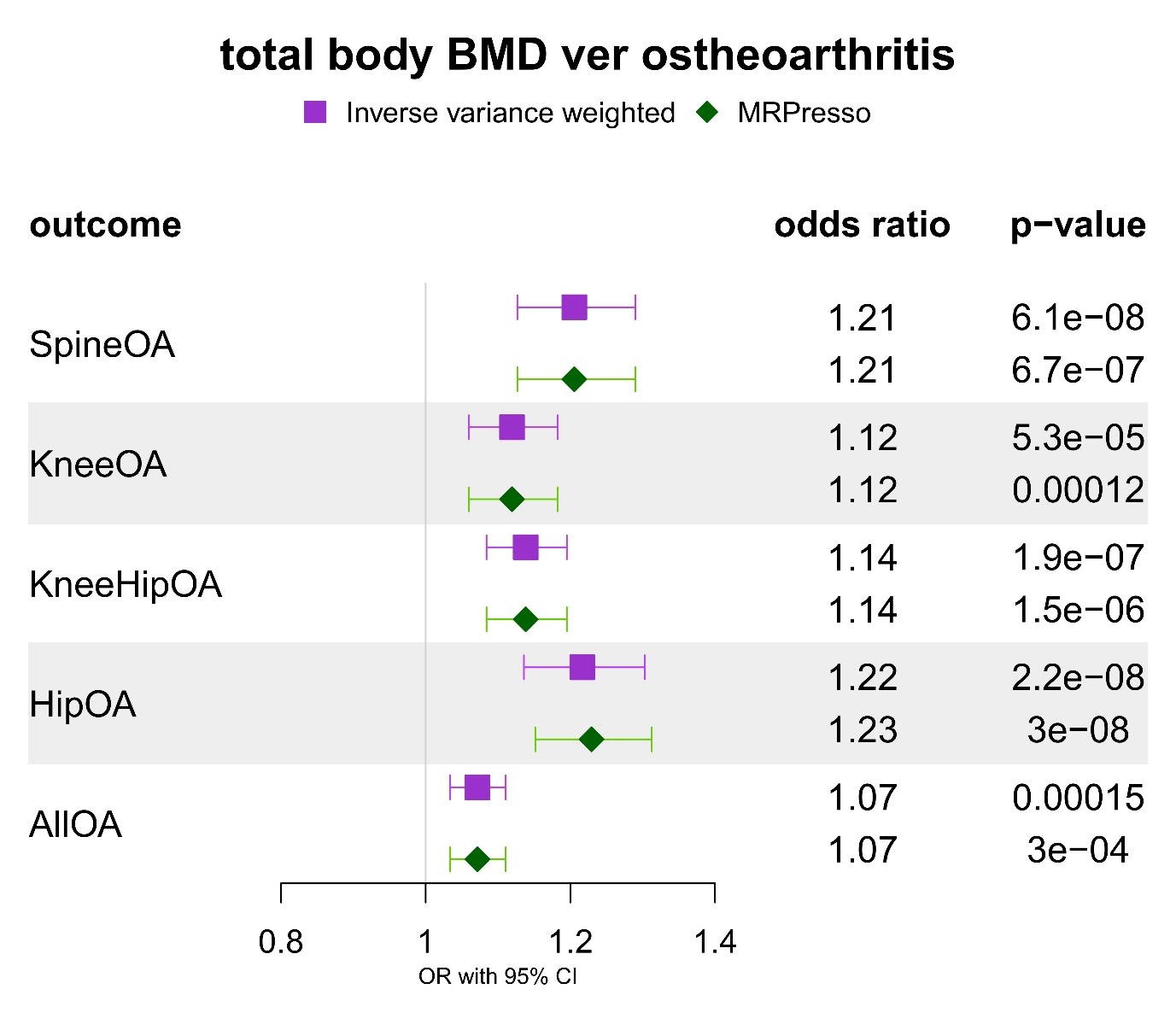

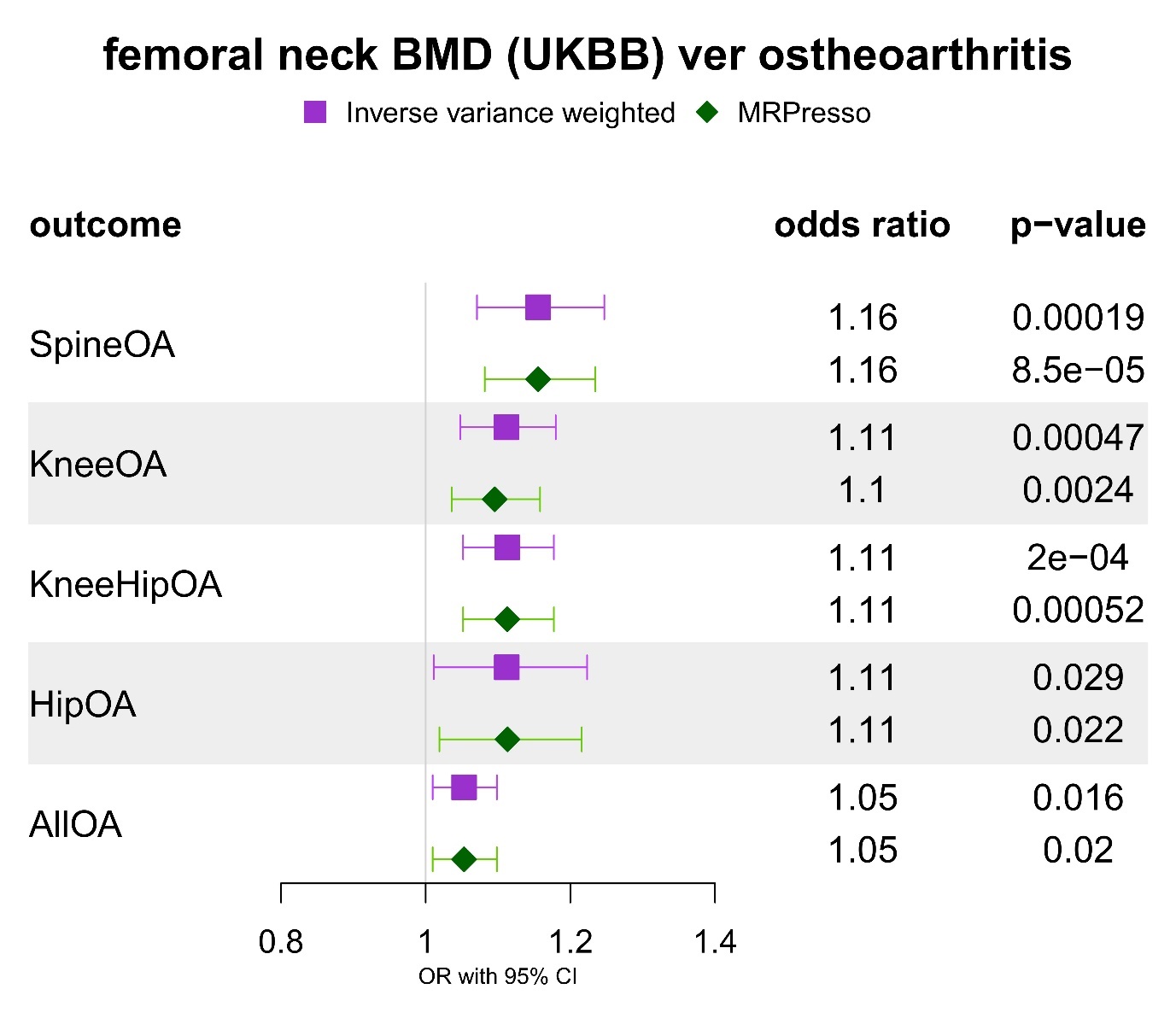
**Supplementary Figure 7.** Two sample Mendelian randomization results for the effect of genetic susceptibility for femoral neck bone mineral density on site-specific and all osteoarthritis. The odds ratios are scaled per SD increase in exposure.

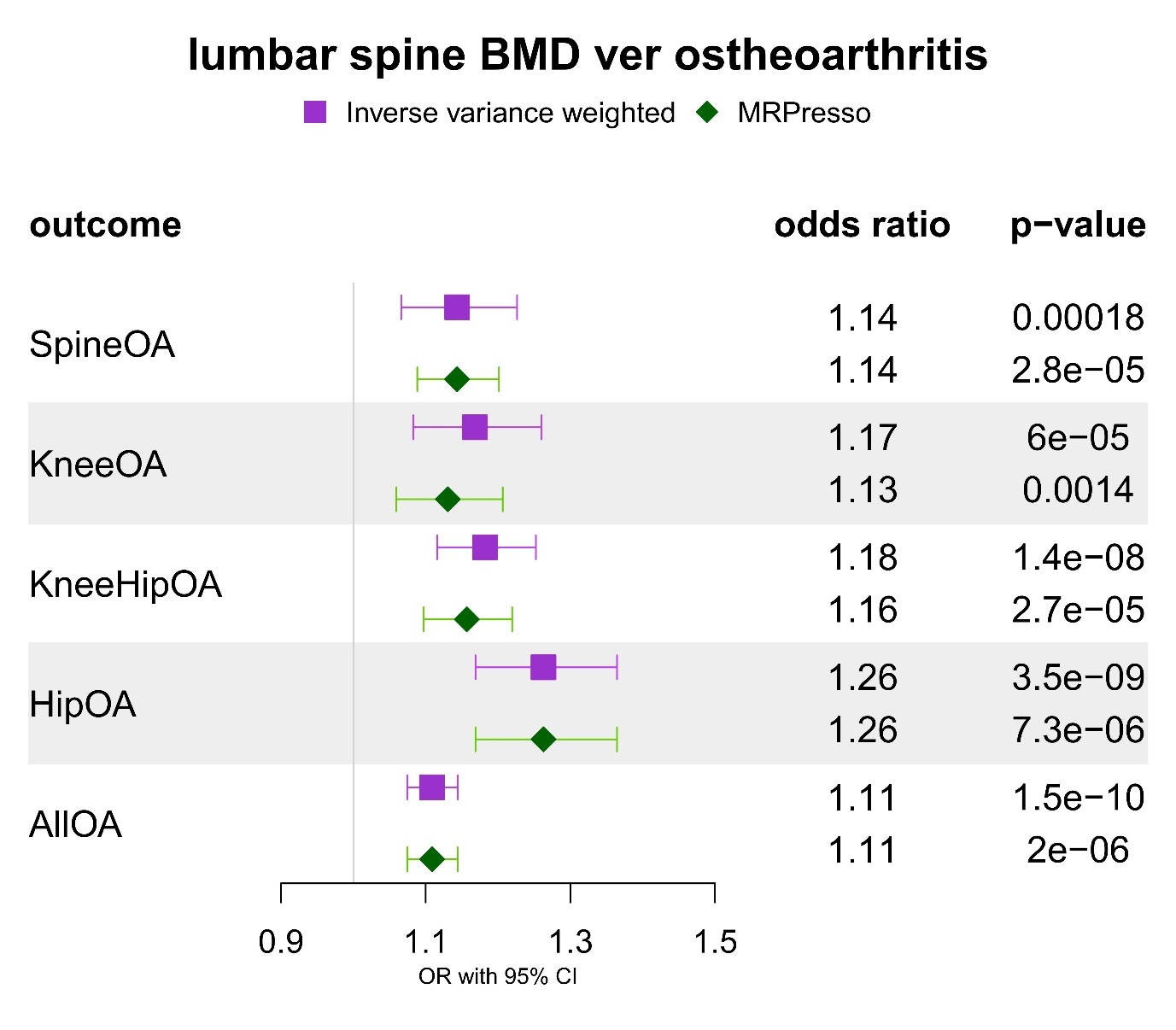
**Supplementary Figure 8.** Two sample Mendelian randomization results for the effect of genetic susceptibility for lumbar spine bone mineral density on site-specific and all osteoarthritis. The odds ratios are scaled per SD increase in exposure.

**Supplementary Figure 9.** Scatter plot for the effect of body mass index on spinal stenosis.

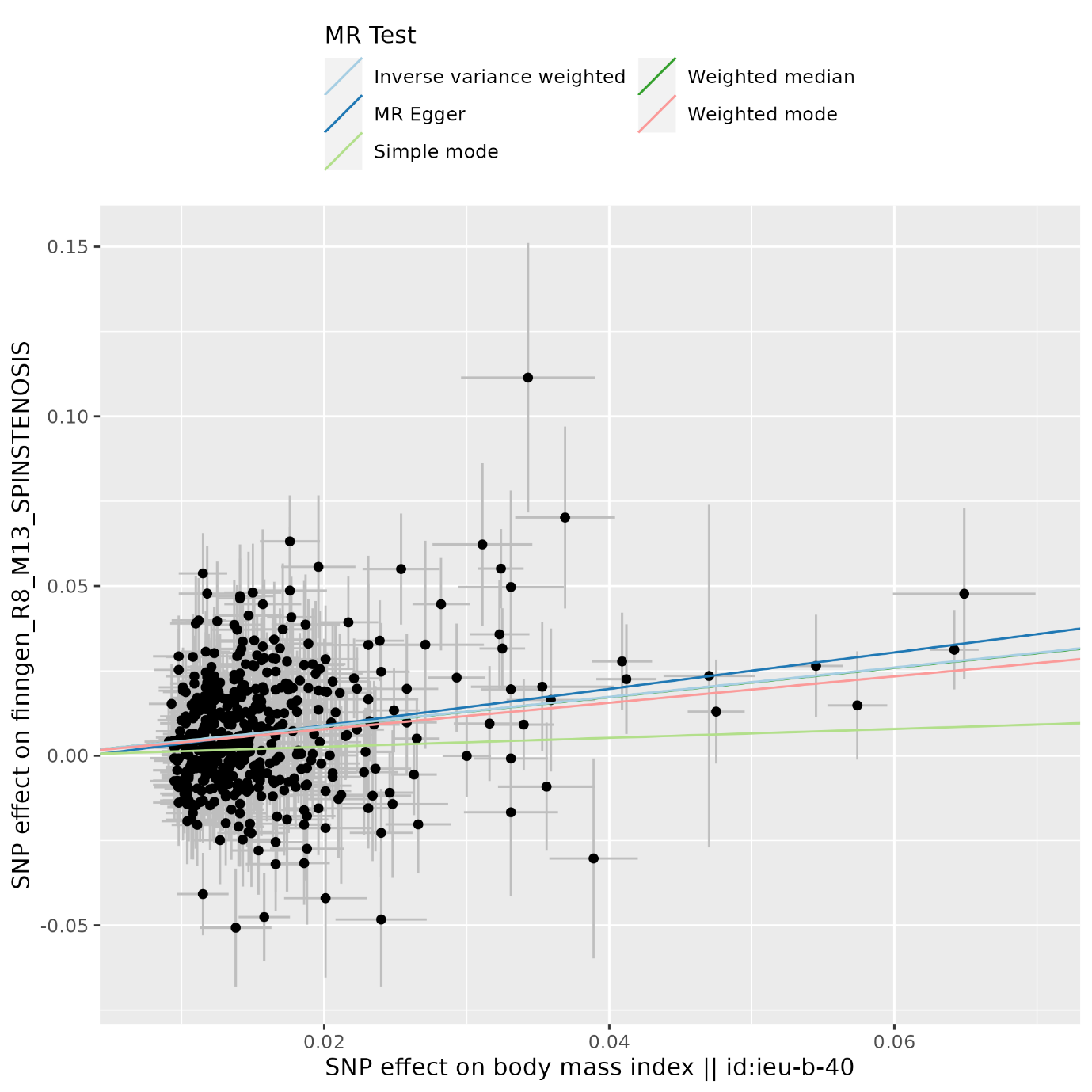

**Supplementary Figure 10.** Scatter plot for the effect of hip circumference on spinal stenosis.

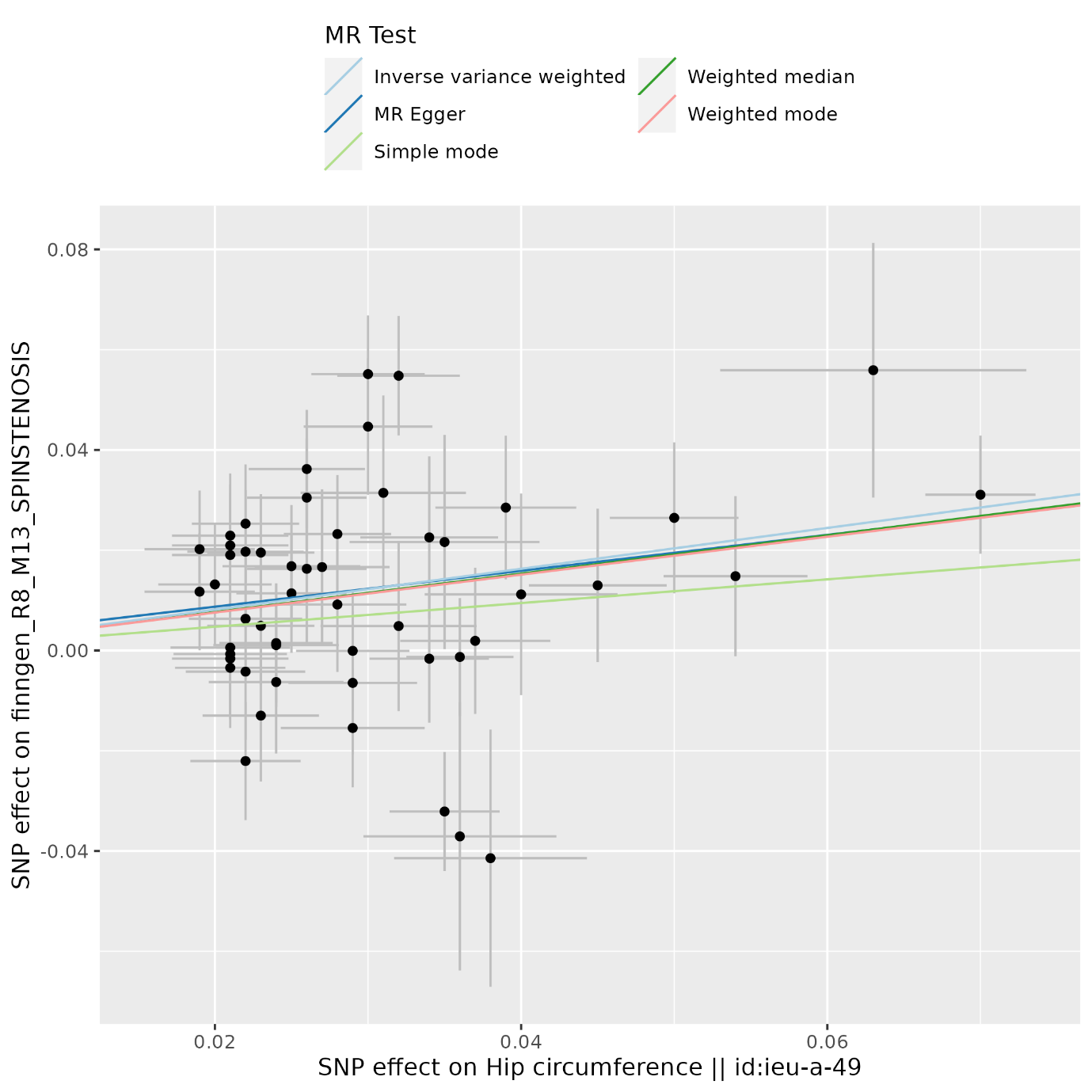

**Supplementary Figure 11.** Scatter plot for the effect of waist circumference on spinal stenosis.
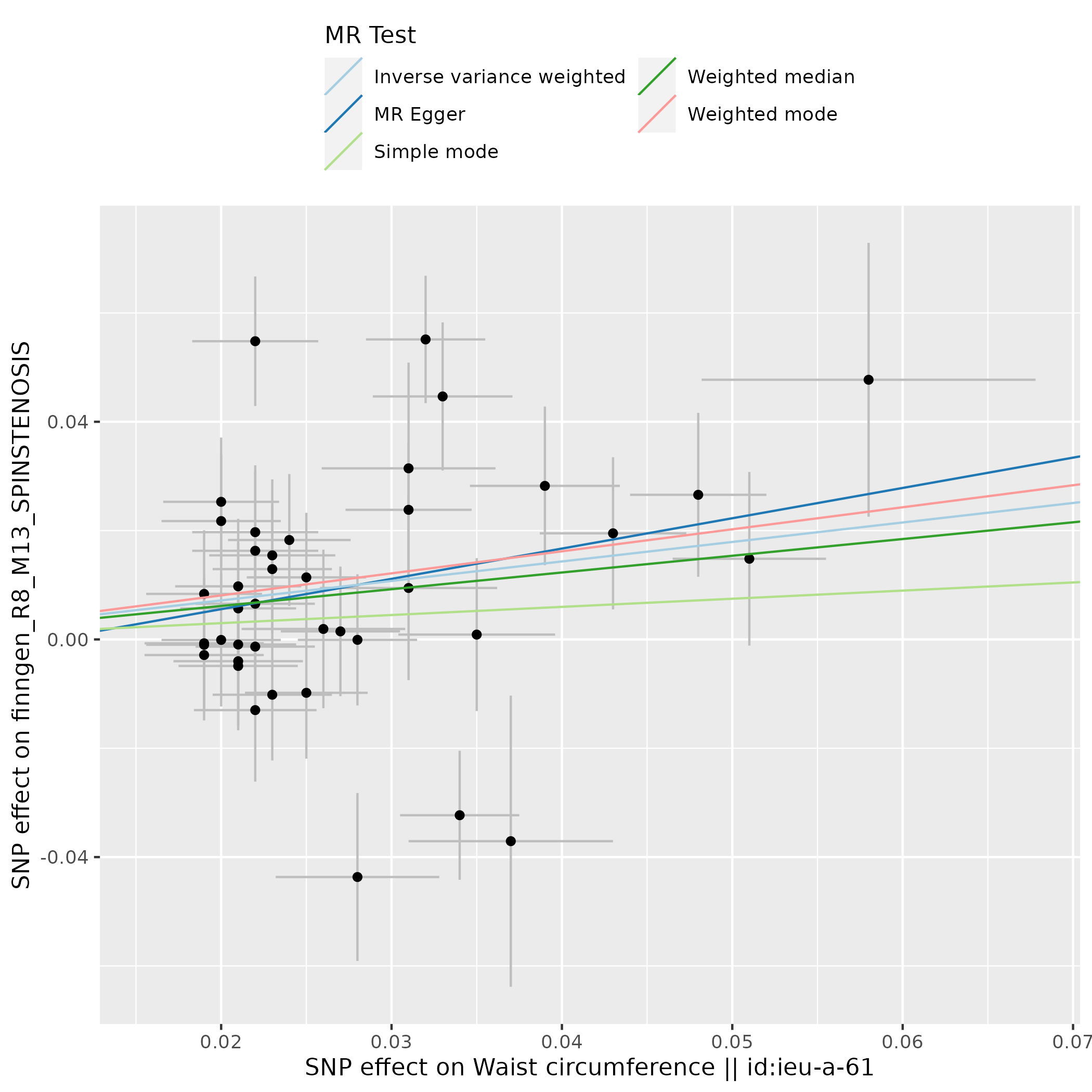

**Supplementary Figure 12.** Scatter plot for the effect of waist-to-hip ratio on spinal stenosis.
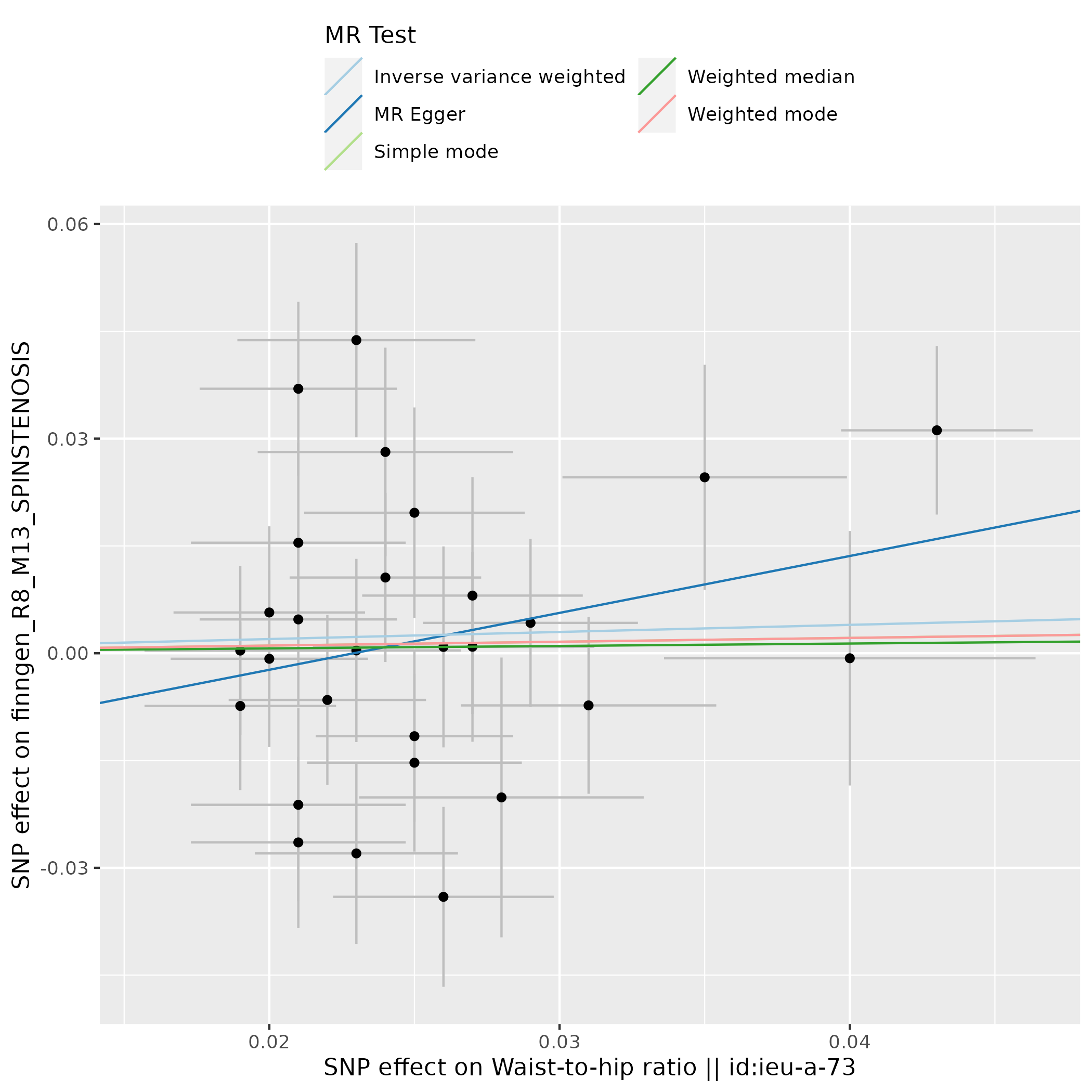

**Supplementary Figure 13.** Scatter plot for the effect of height on spinal stenosis.

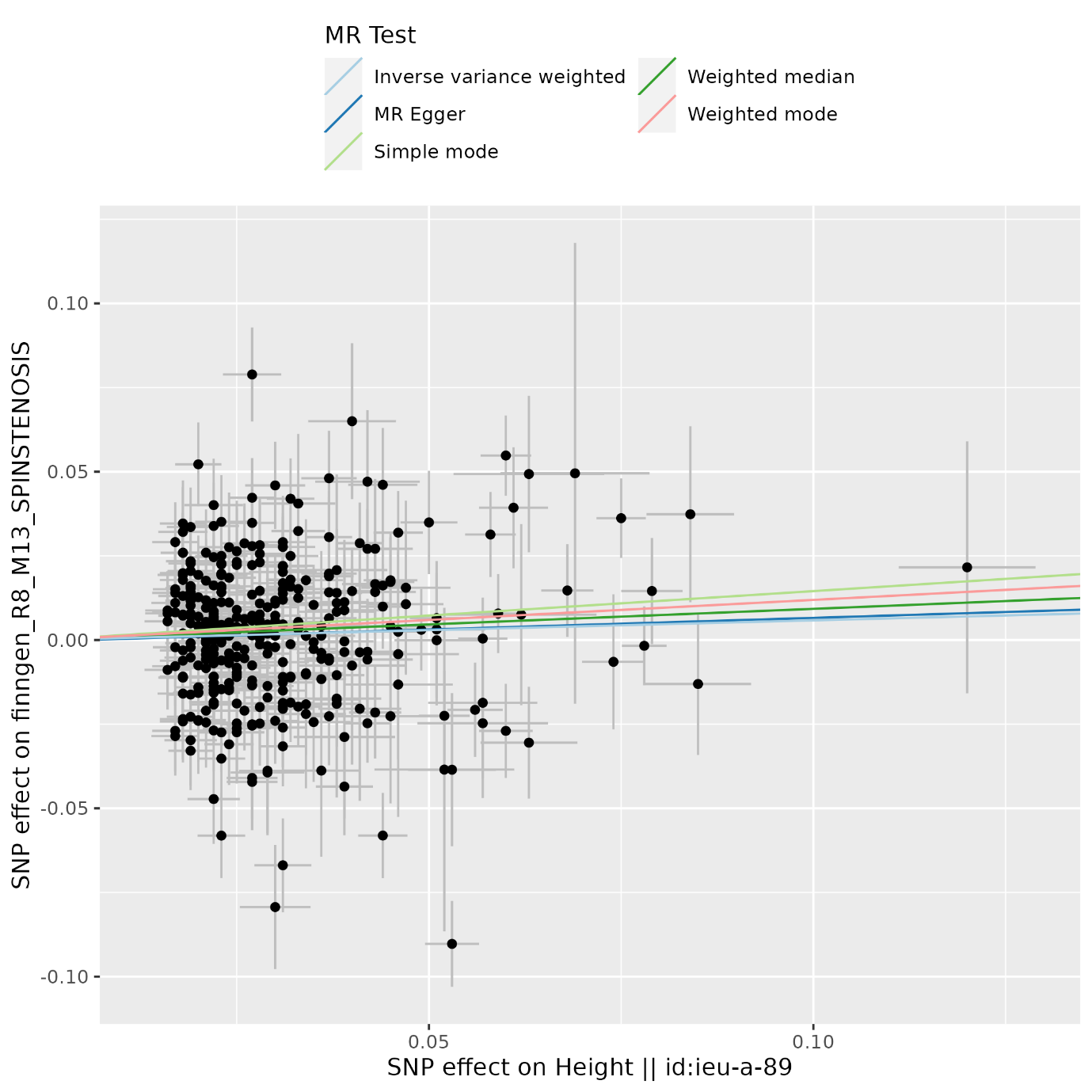

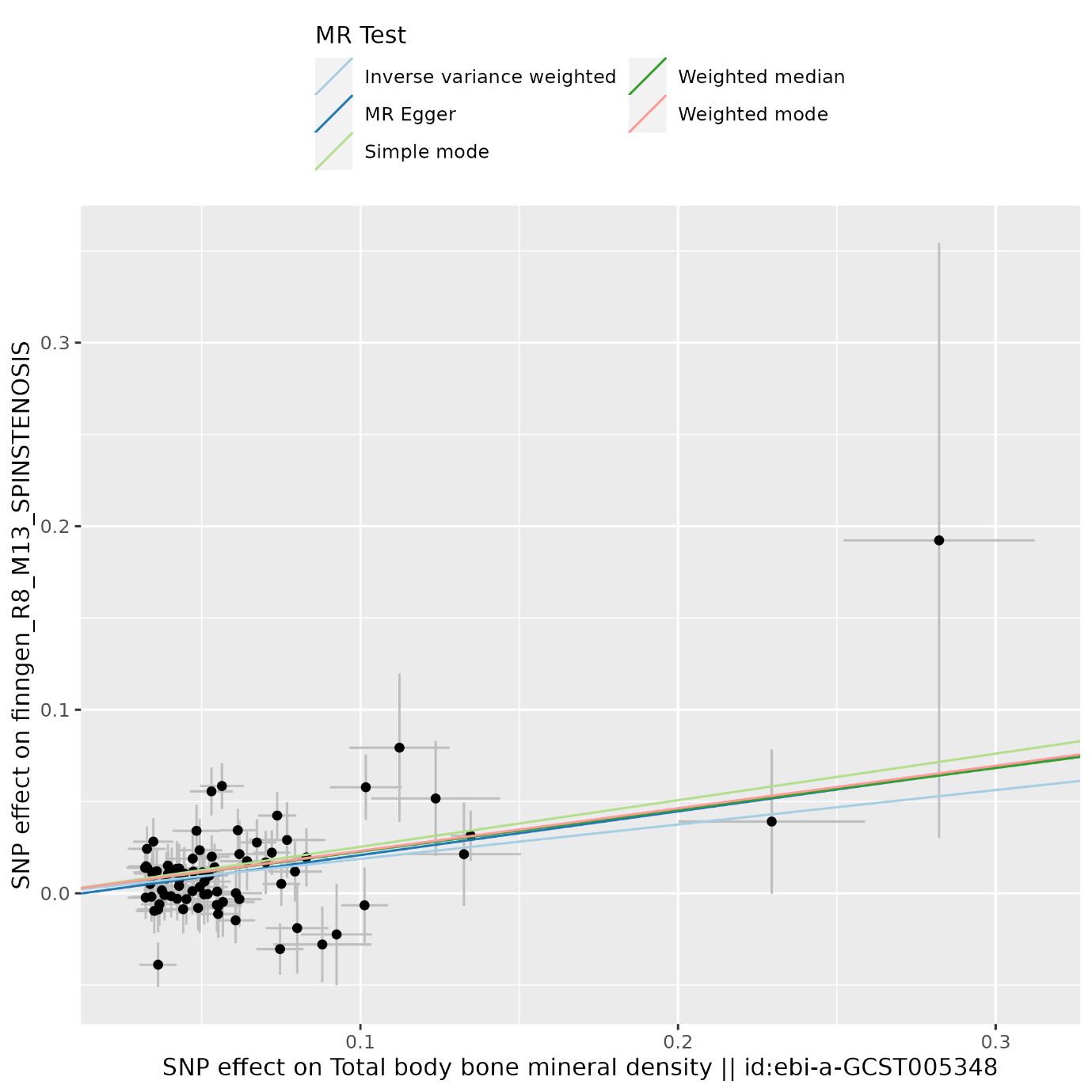
**Supplementary Figure 14.** Scatter plot for the effect of total body bone mineral density on spinal stenosis.

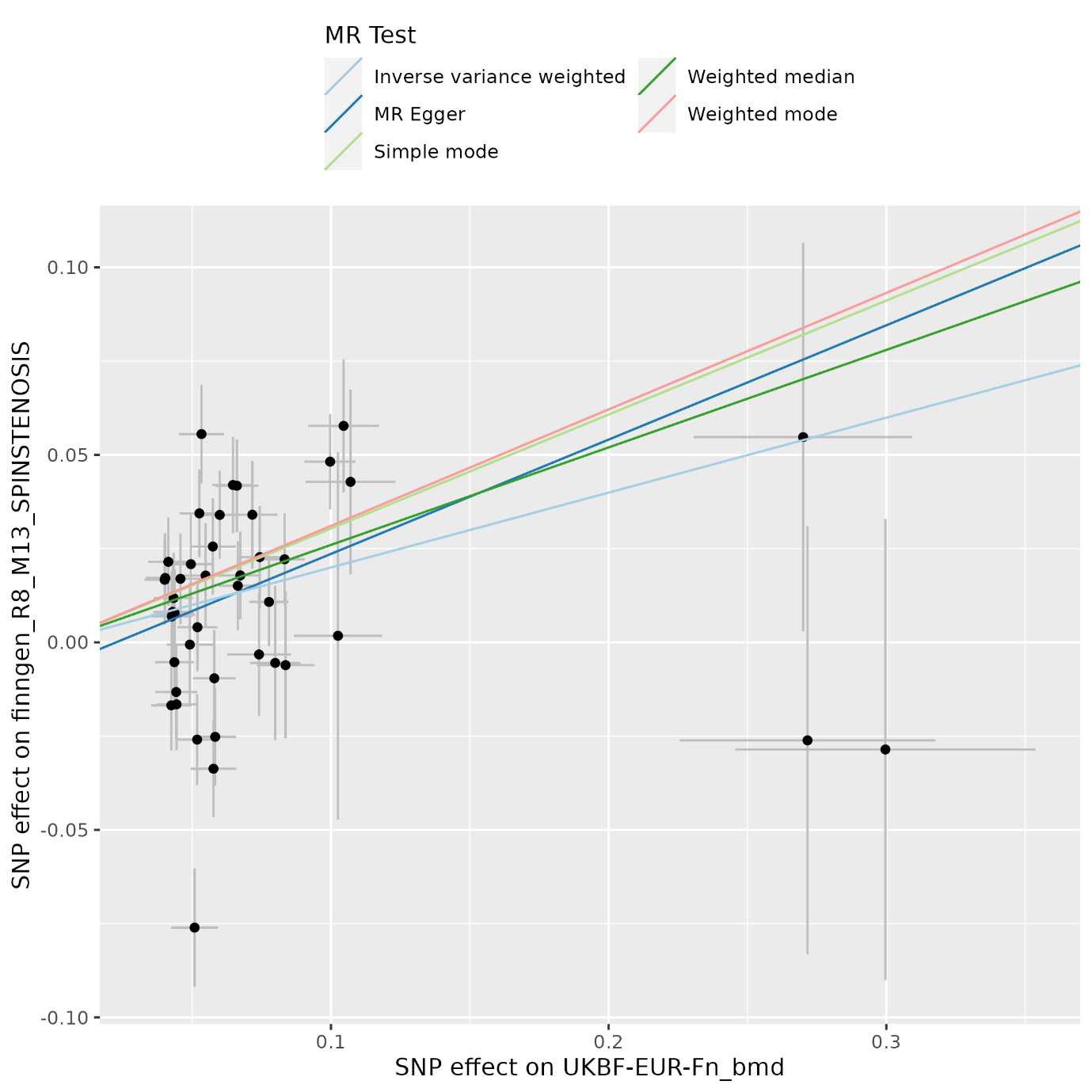
**Supplementary Figure 15.** Scatter plot for the effect of femoral neck bone mineral density on spinal stenosis.

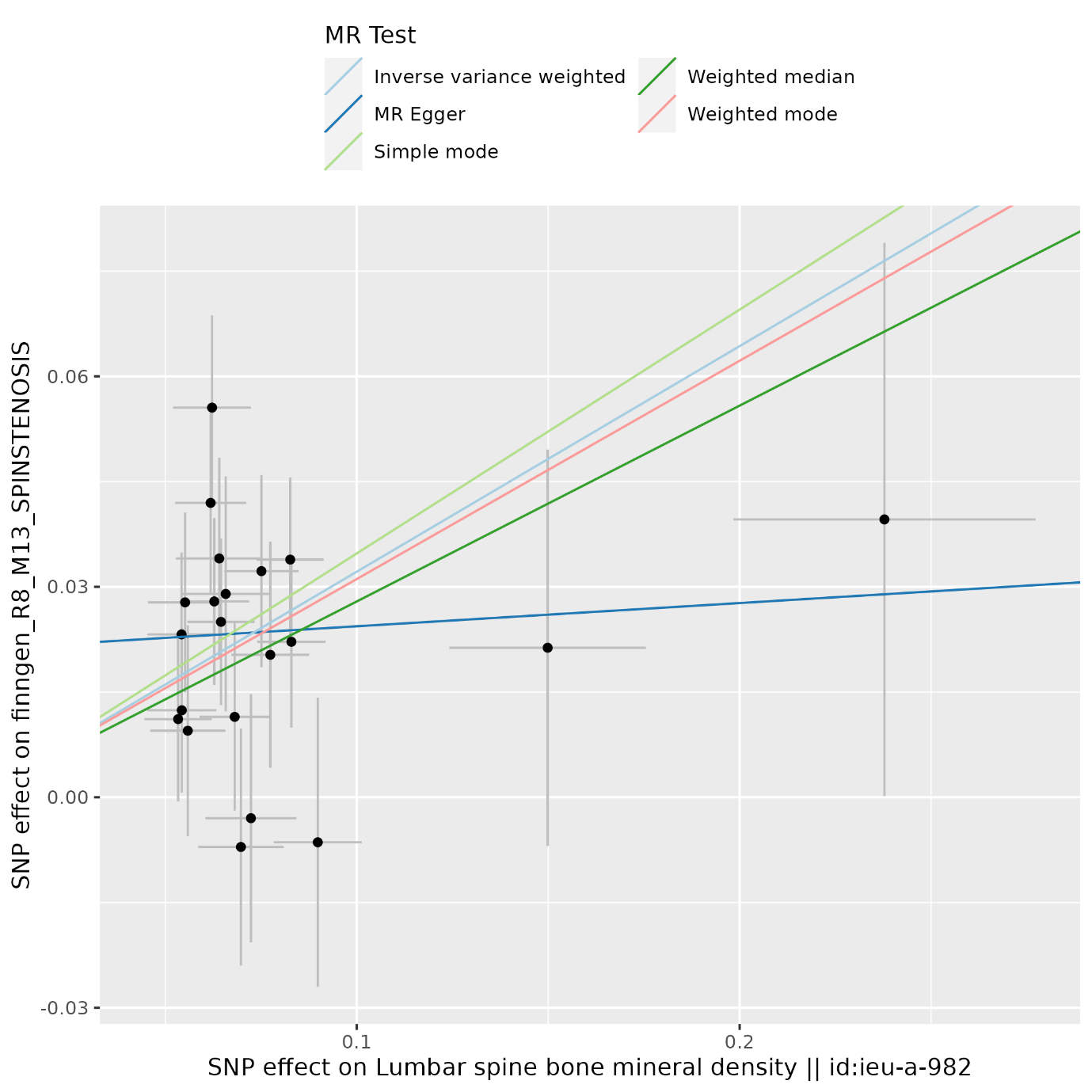
**Supplementary Figure 16.** Scatter plot for the effect of lumbar spine bone mineral density on spinal stenosis.

**Supplementary Figure 17.** Scatter plot for the effect of albumin-adjusted circulating calcium on spinal stenosis.

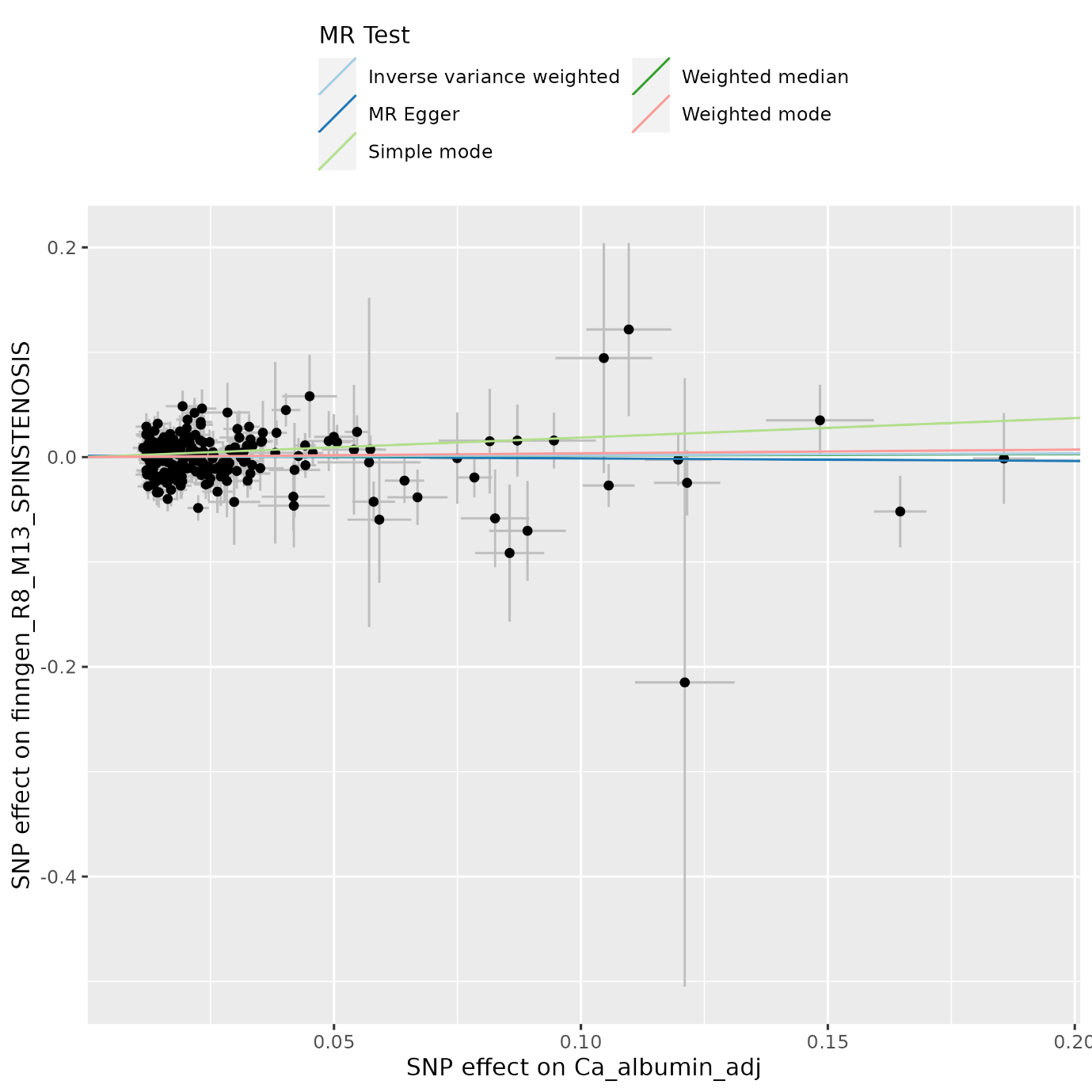

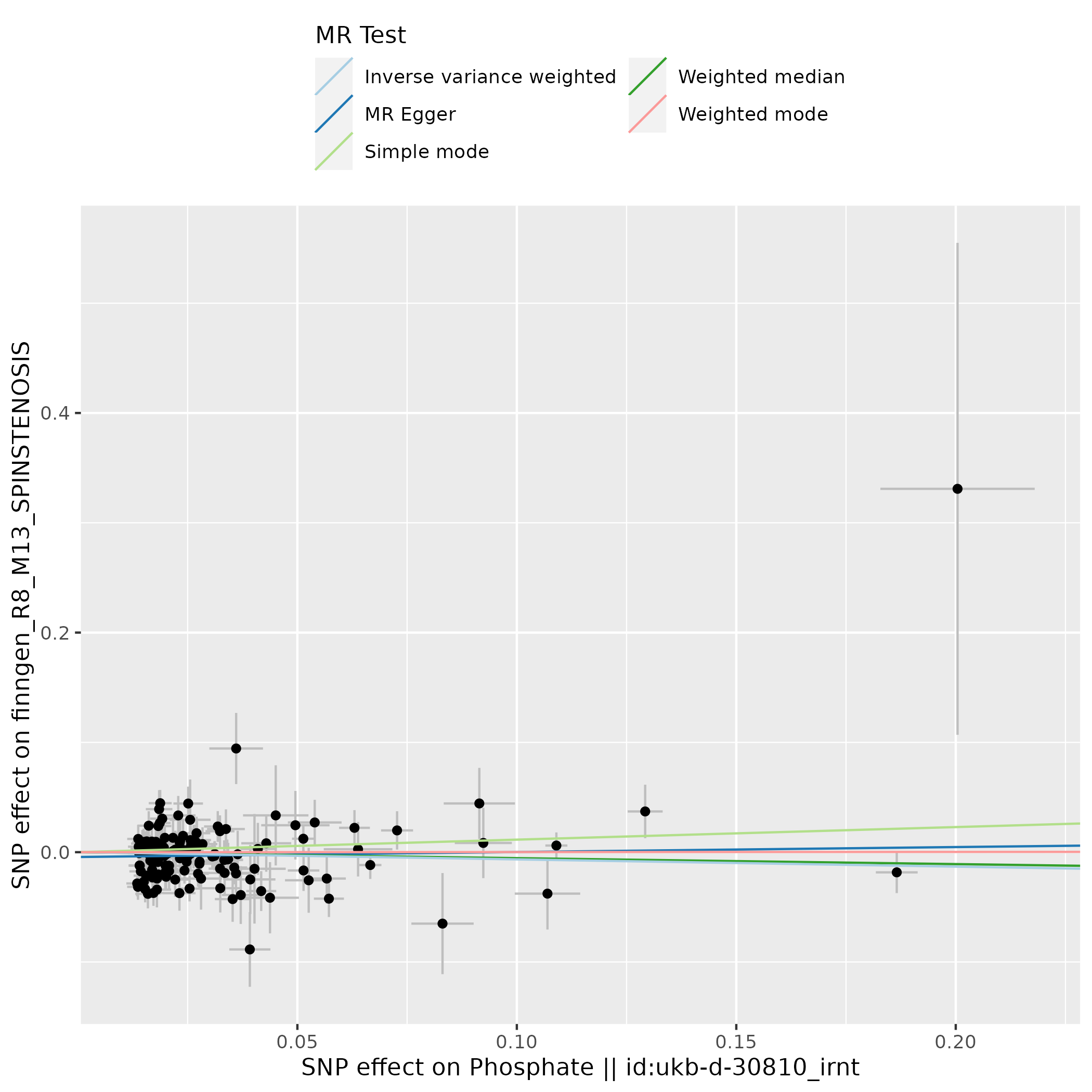
**Supplementary Figure 18.** Scatter plot for the effect of circulating phosphate on spinal stenosis.

**Supplementary Figure 19.** Scatter plot for the effect of all osteoarthritis on spinal stenosis.

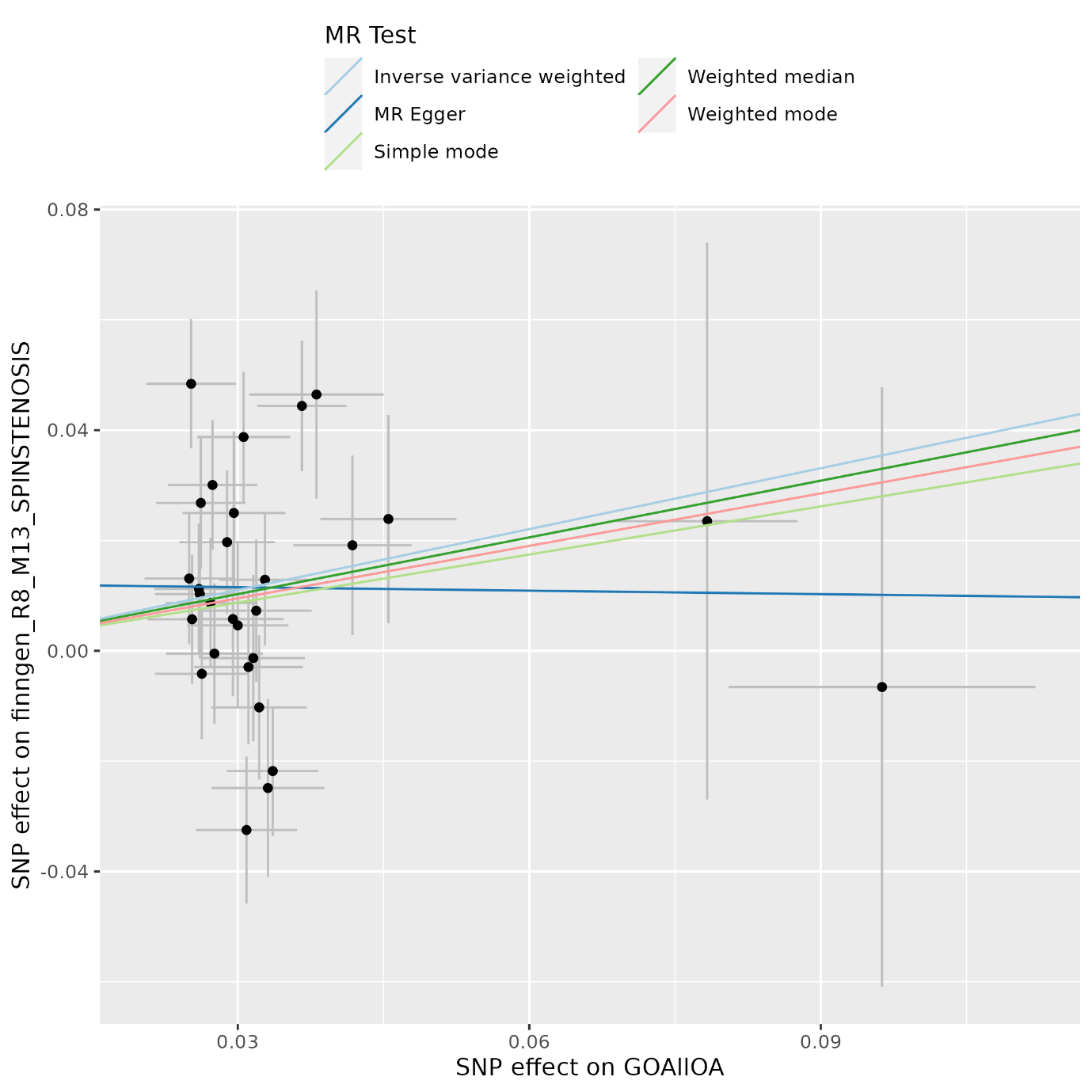
**Supplementary Figure 20.** Scatter plot for the effect of hip osteoarthritis on
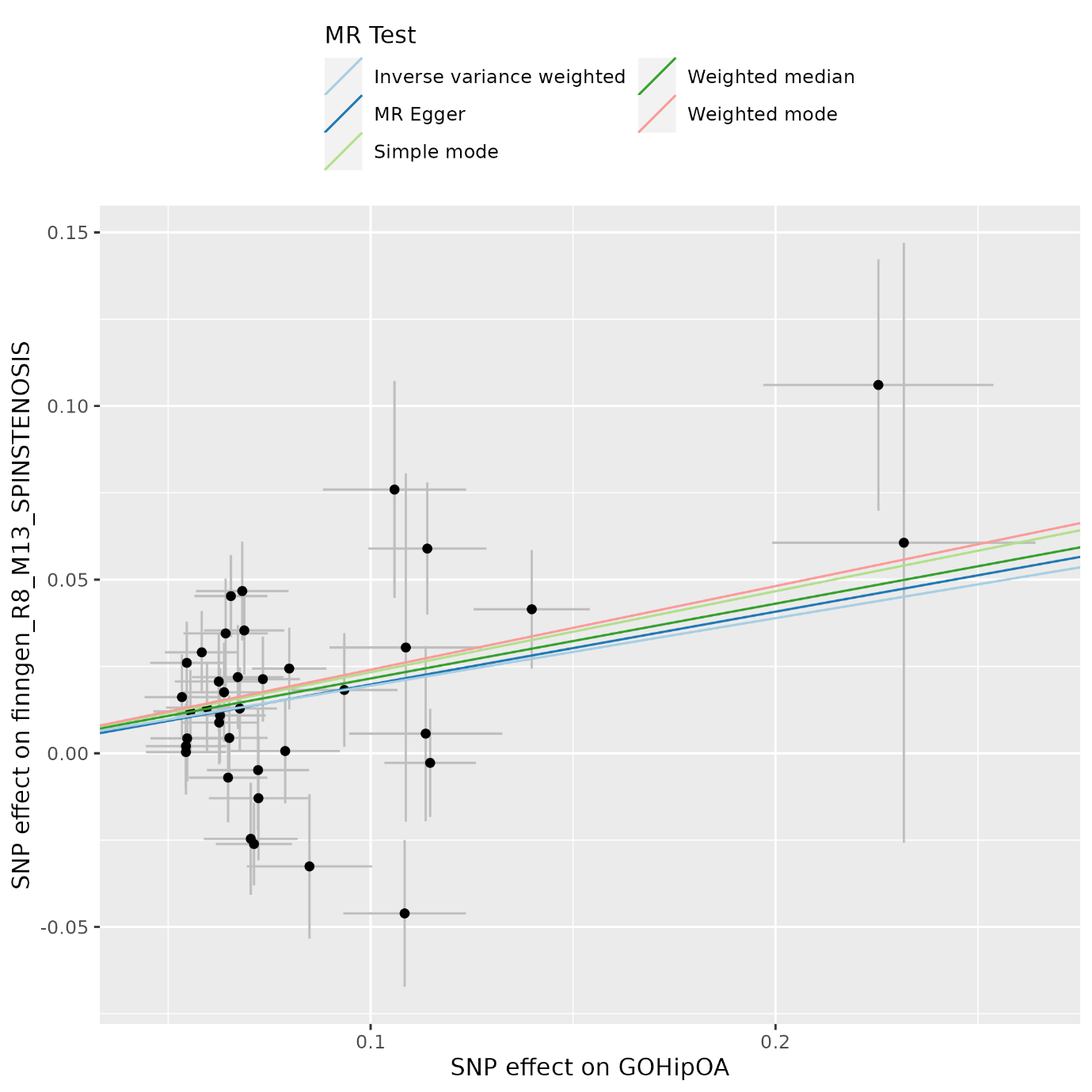
spinal stenosis.

**Supplementary Figure 21.** Scatter plot for the effect of knee and/or hip osteoarthritis on spinal stenosis.

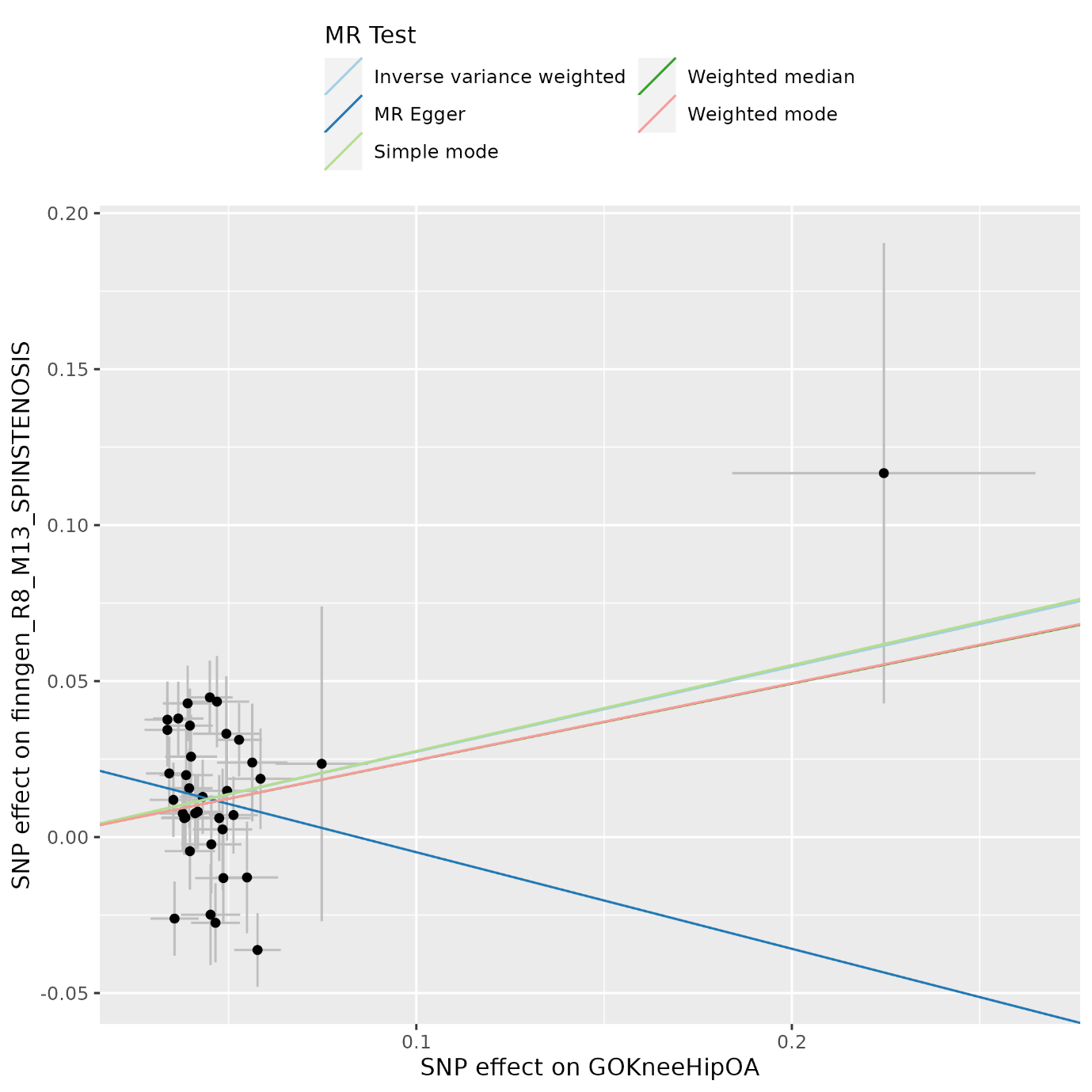

**Supplementary Figure 22.** Scatter plot for the effect of knee osteoarthritis on spinal stenosis.

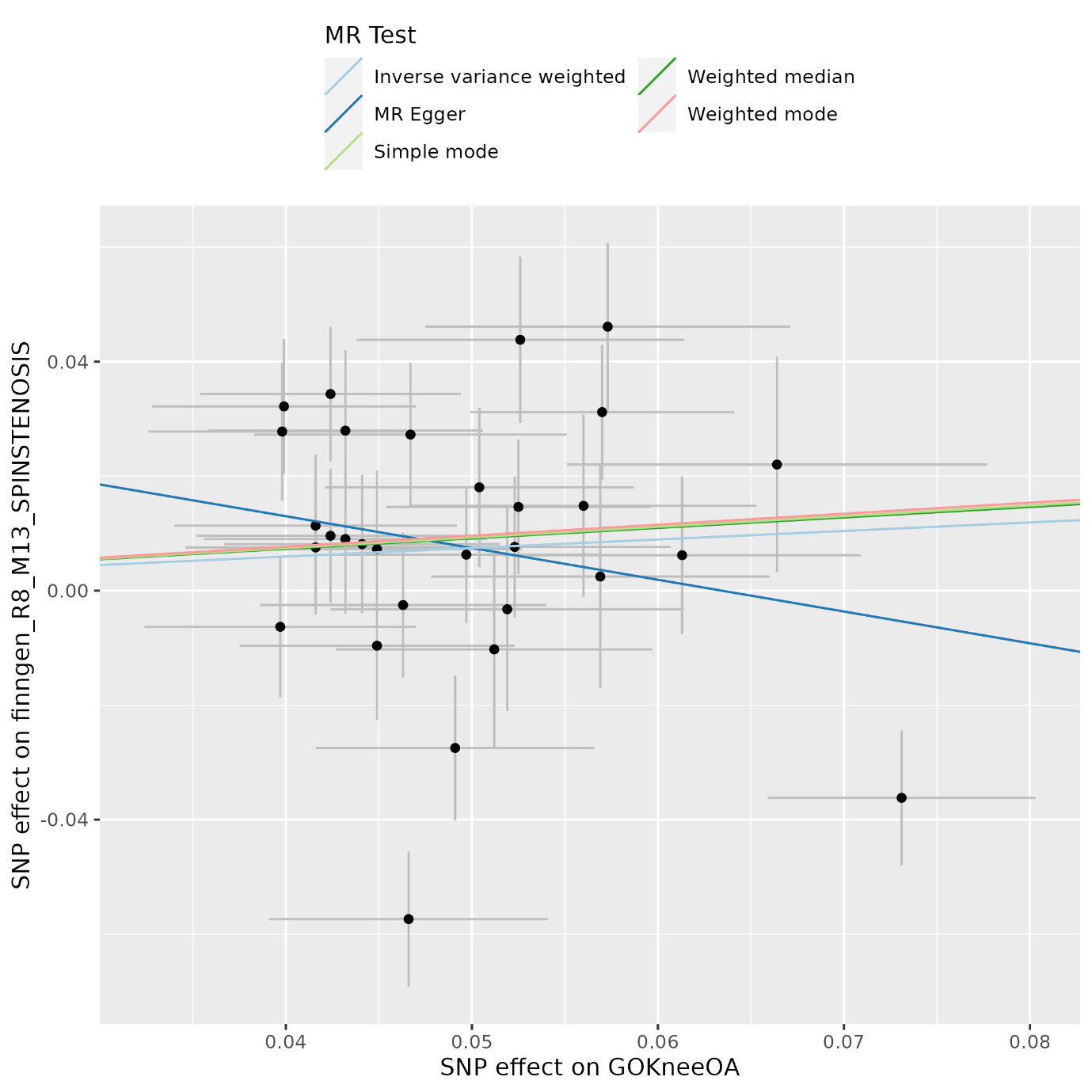

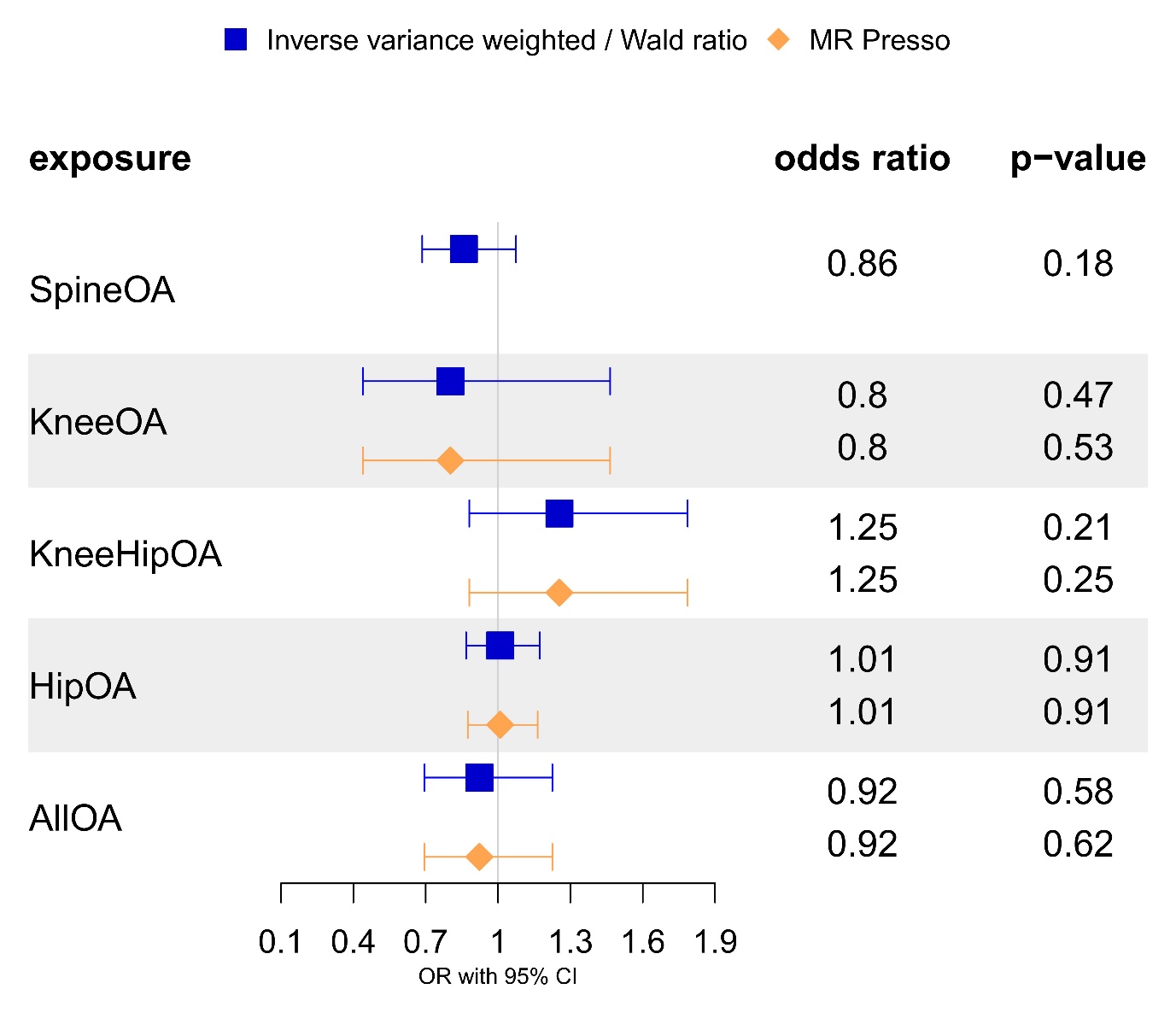
**Supplementary Figure 23.** Two sample Mendelian randomization results for the effect of genetic liability for osteoarthritis on spinal stenosis (UKBB). The odds ratios are scaled per doubling in risk of exposure.

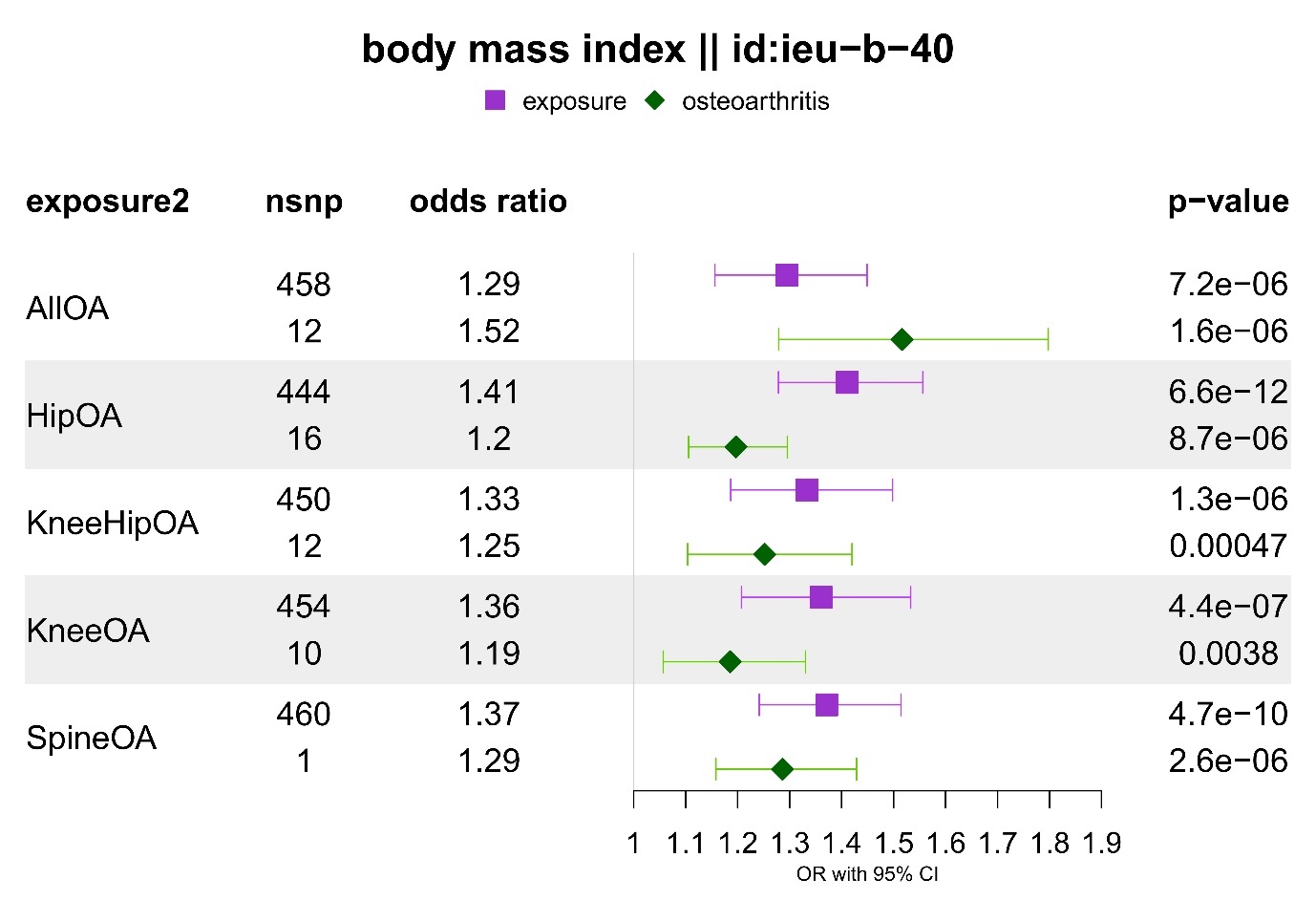
**Supplementary Figure 24.** Multivariable Mendelian randomization results for the jointly modelled effect of genetic susceptibility for body mass index and liability for osteoarthritis (all or site-specific) on spinal stenosis (FinnGen). The odds ratios are scaled per SD increase of BMI and doubling in the risk of osteoarthritis.

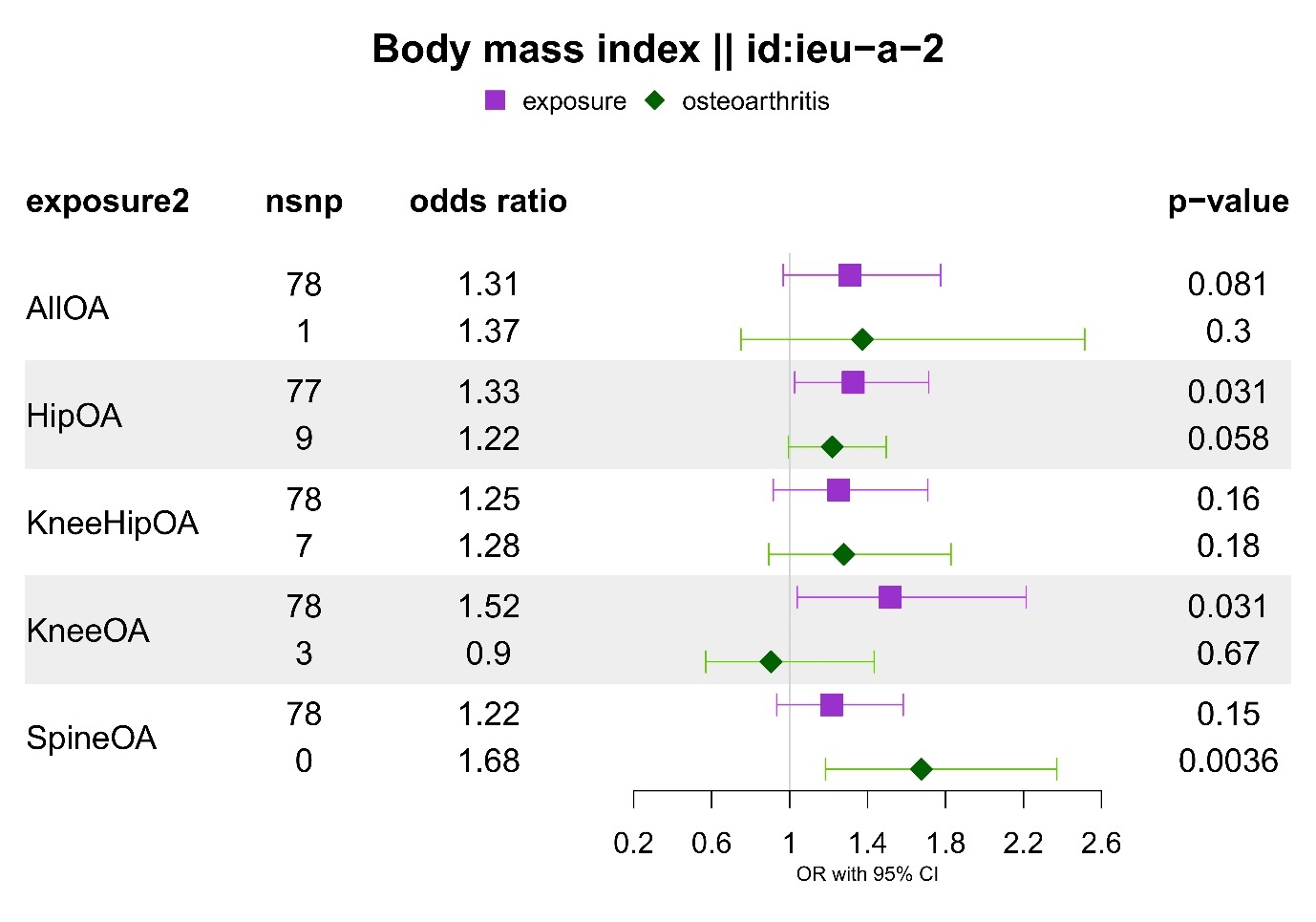
**Supplementary Figure 25.** Multivariable Mendelian randomization results for the jointly modelled effect of genetic susceptibility for body mass index and liability for osteoarthritis (all or site-specific) on spinal stenosis (UKBB). The odds ratios are scaled per SD increase of BMI and doubling in the risk of osteoarthritis.

**Supplementary Figure 26.** Multivariable Mendelian randomization results for the jointly modelled effect of genetic susceptibility for height and liability for osteoarthritis (all or site-specific) on spinal stenosis (FinnGen). The odds ratios are scaled per SD increase of BMI and doubling in the risk of osteoarthritis
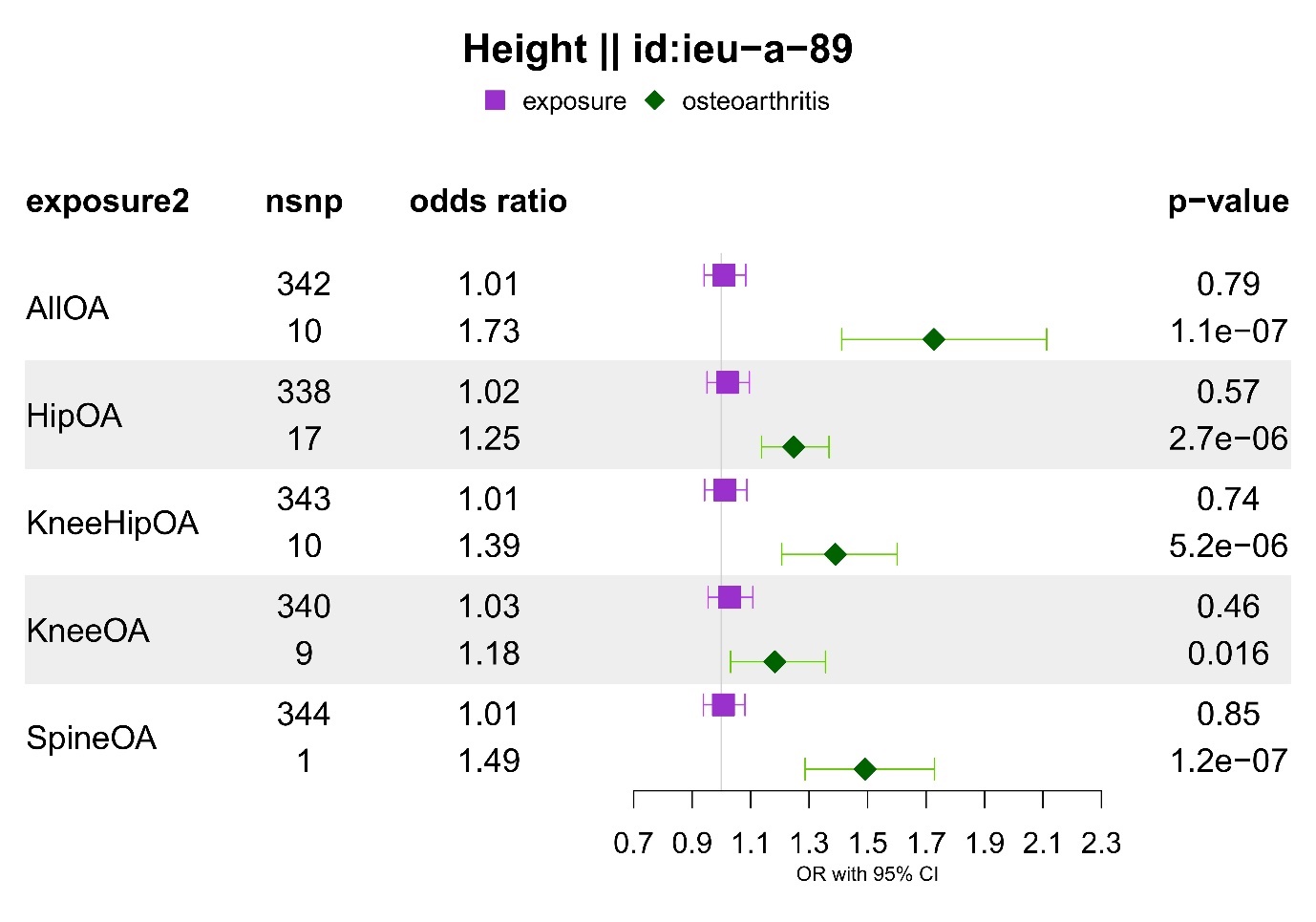
.

**Supplementary Figure 27.** Multivariable Mendelian randomization results for the jointly modelled effect of genetic susceptibility for height and liability for osteoarthritis (all or site-specific) on spinal stenosis (UKBB). The odds ratios are scaled per SD increase of BMI and doubling in the risk of osteoarthritis
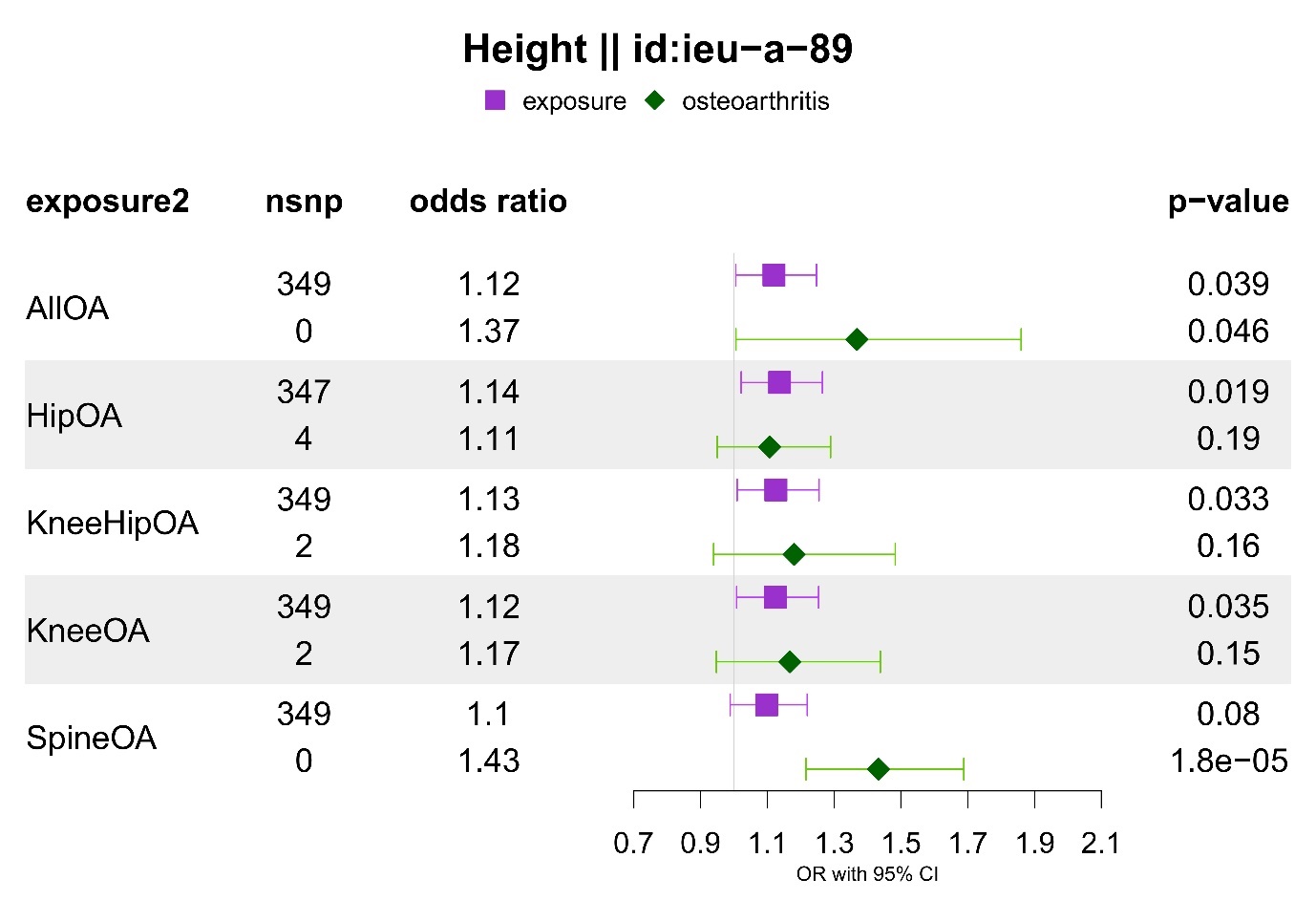
.

**Supplementary Figure 28.** Multivariable Mendelian randomization results for the jointly modelled effect of genetic susceptibility for total body bone mineral density and liability for osteoarthritis (all or site-specific) on spinal stenosis (FinnGen). The odds ratios are scaled per SD increase of BMI and doubling in the risk of osteoarthritis
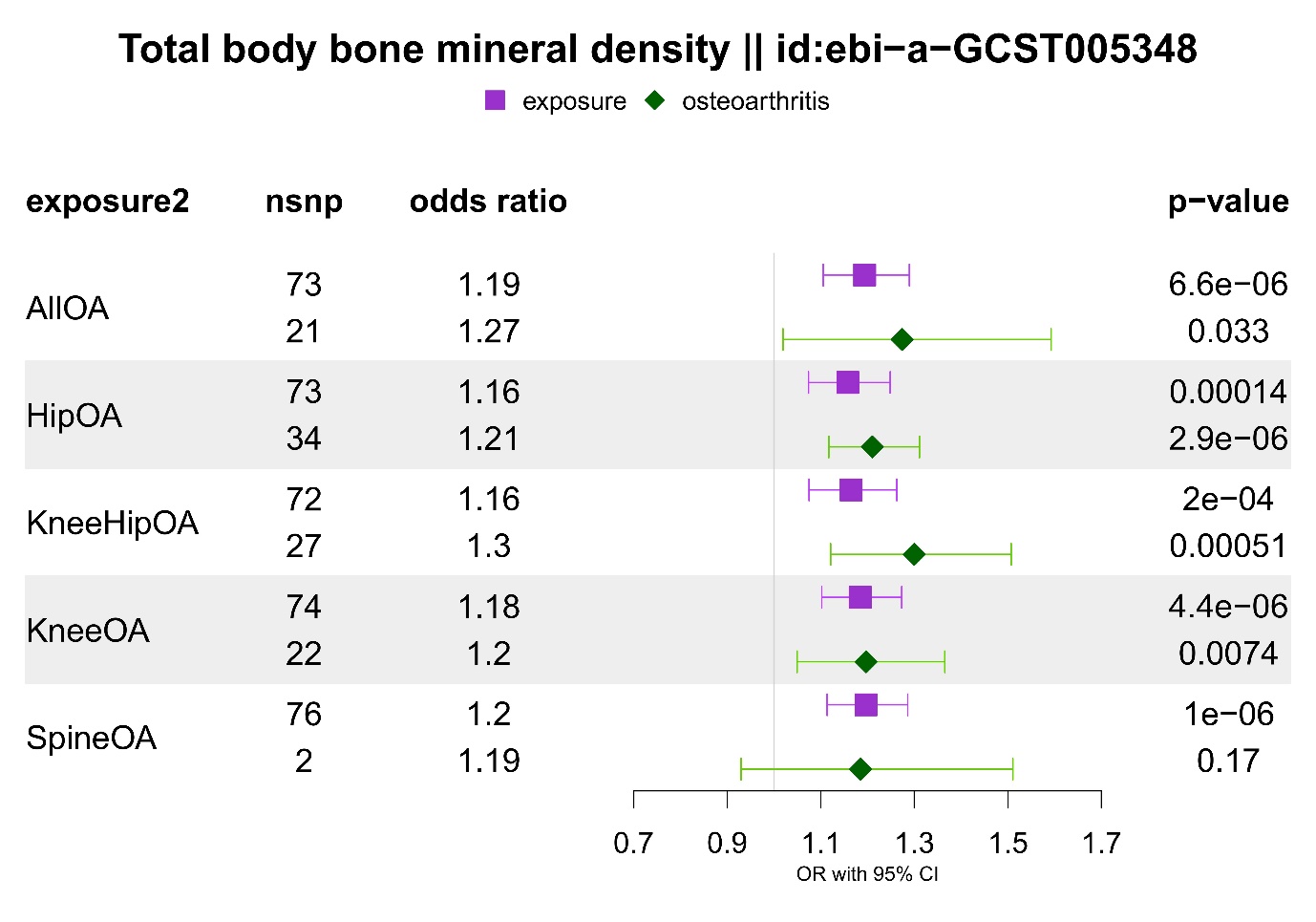
.

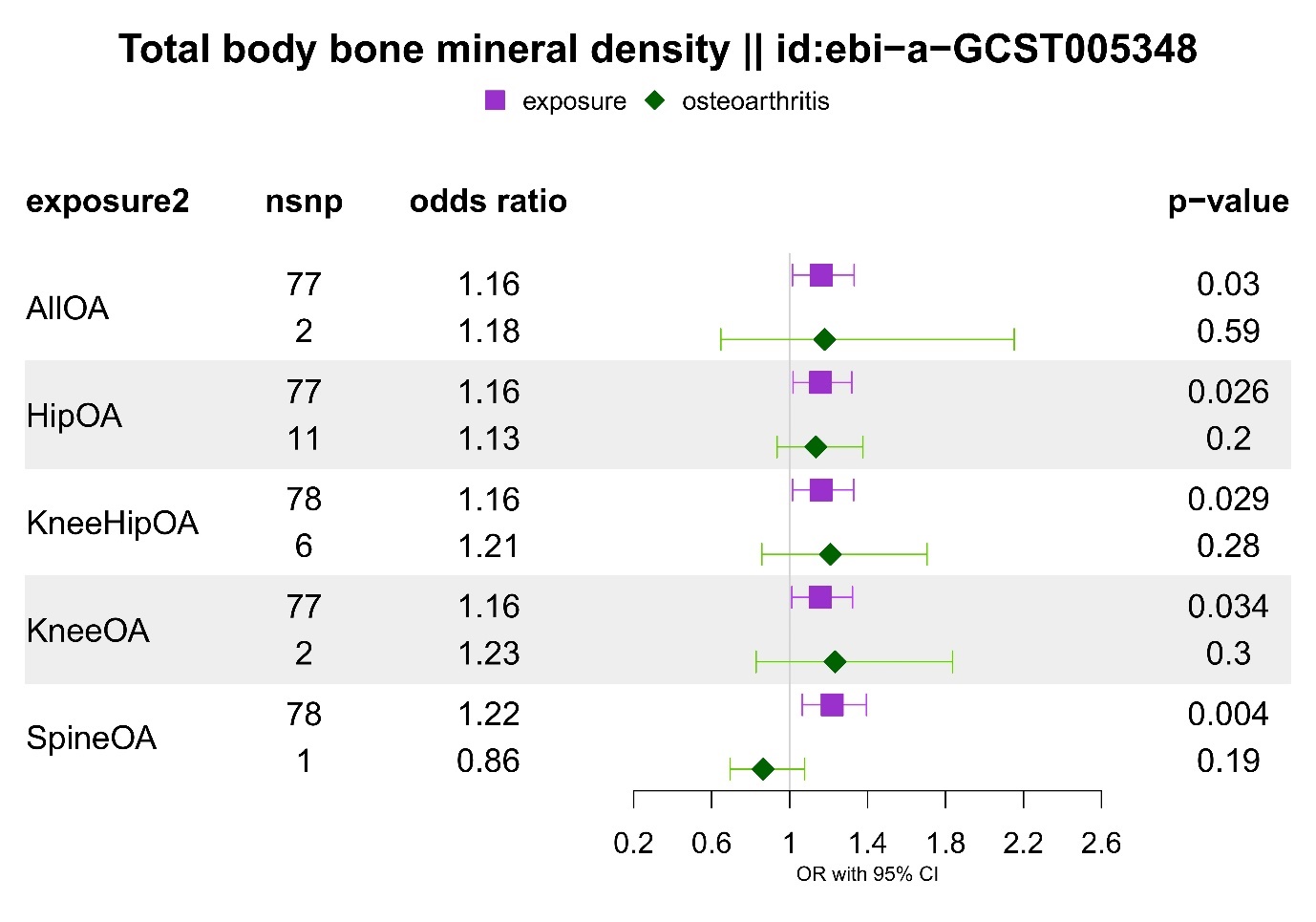
**Supplementary Figure 29.** Multivariable Mendelian randomization results for the jointly modelled effect of genetic susceptibility for total body bone mineral density and liability for osteoarthritis (all or site-specific) on spinal stenosis (UKBB). The odds ratios are scaled per SD increase of BMI and doubling in the risk of osteoarthritis.

**Supplementary Figure 30.** Multivariable Mendelian randomization results for the jointly modelled effect of genetic susceptibility for femoral neck bone mineral density and liability for osteoarthritis (all or site-specific) on spinal stenosis (FinnGen). The odds ratios are scaled per SD increase of BMI and doubling in the risk of osteoarthritis
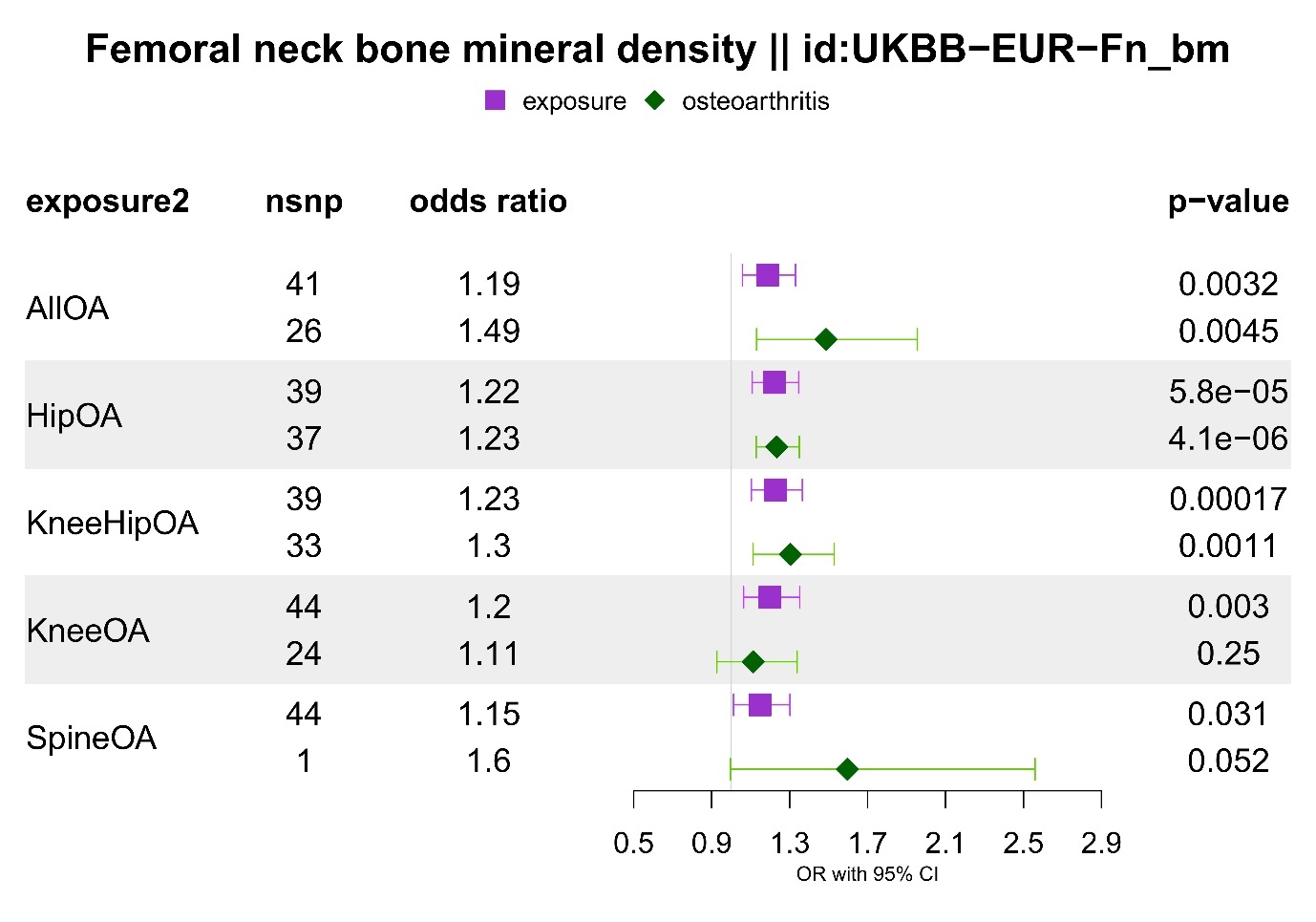
.

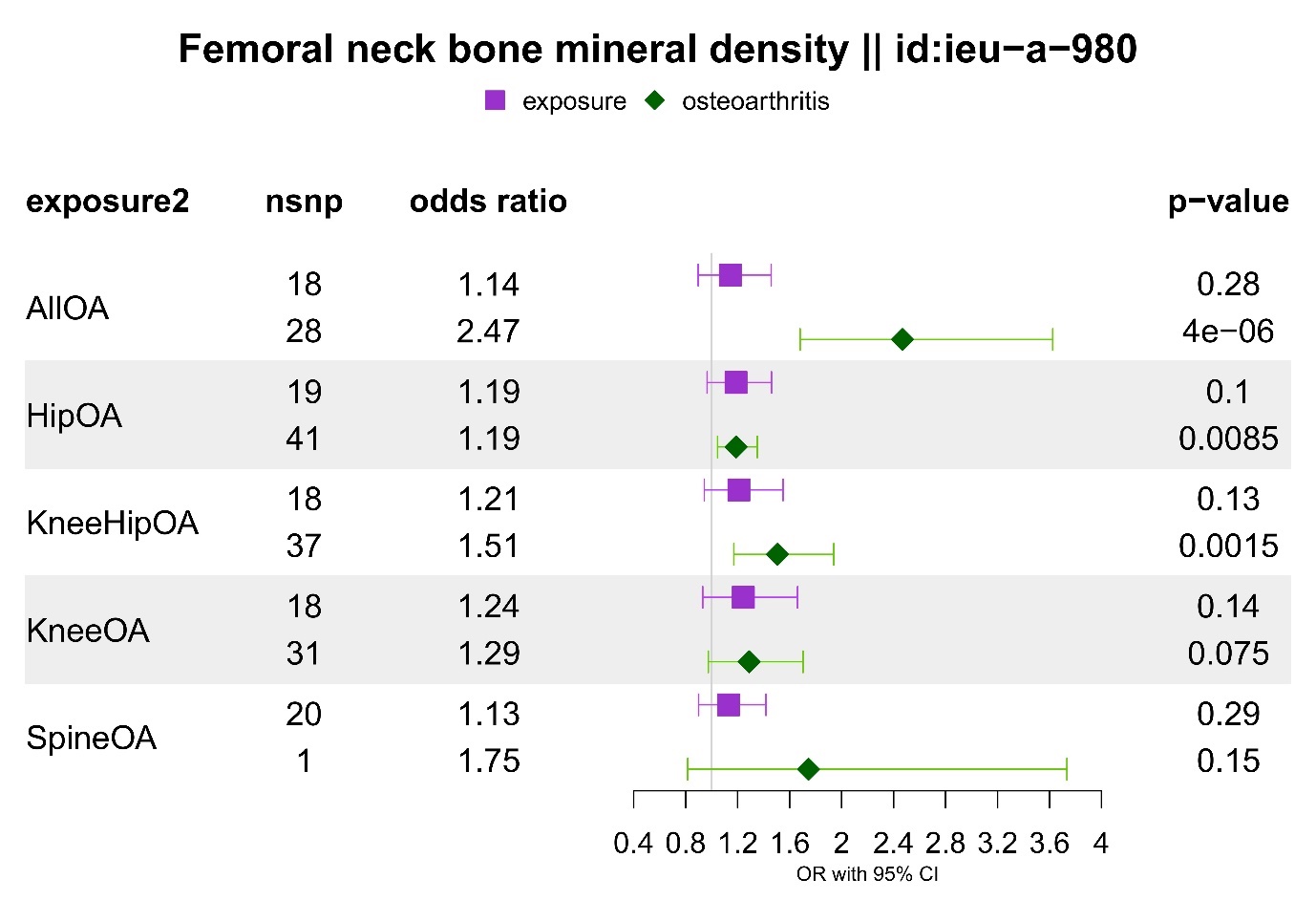
**Supplementary Figure 31.** Multivariable Mendelian randomization results for the jointly modelled effect of genetic susceptibility for femoral neck bone mineral density and liability for osteoarthritis (all or site-specific) on spinal stenosis (UKBB). The odds ratios are scaled per SD increase of BMI and doubling in the risk of osteoarthritis.

**Supplementary Figure 32.** Multivariable Mendelian randomization results for the jointly modelled effect of genetic susceptibility for lumbar spine bone mineral density and liability for osteoarthritis (all or site-specific) on spinal stenosis (FinnGen). The odds ratios are scaled per SD increase of BMI and doubling in the risk of osteoarthritis.
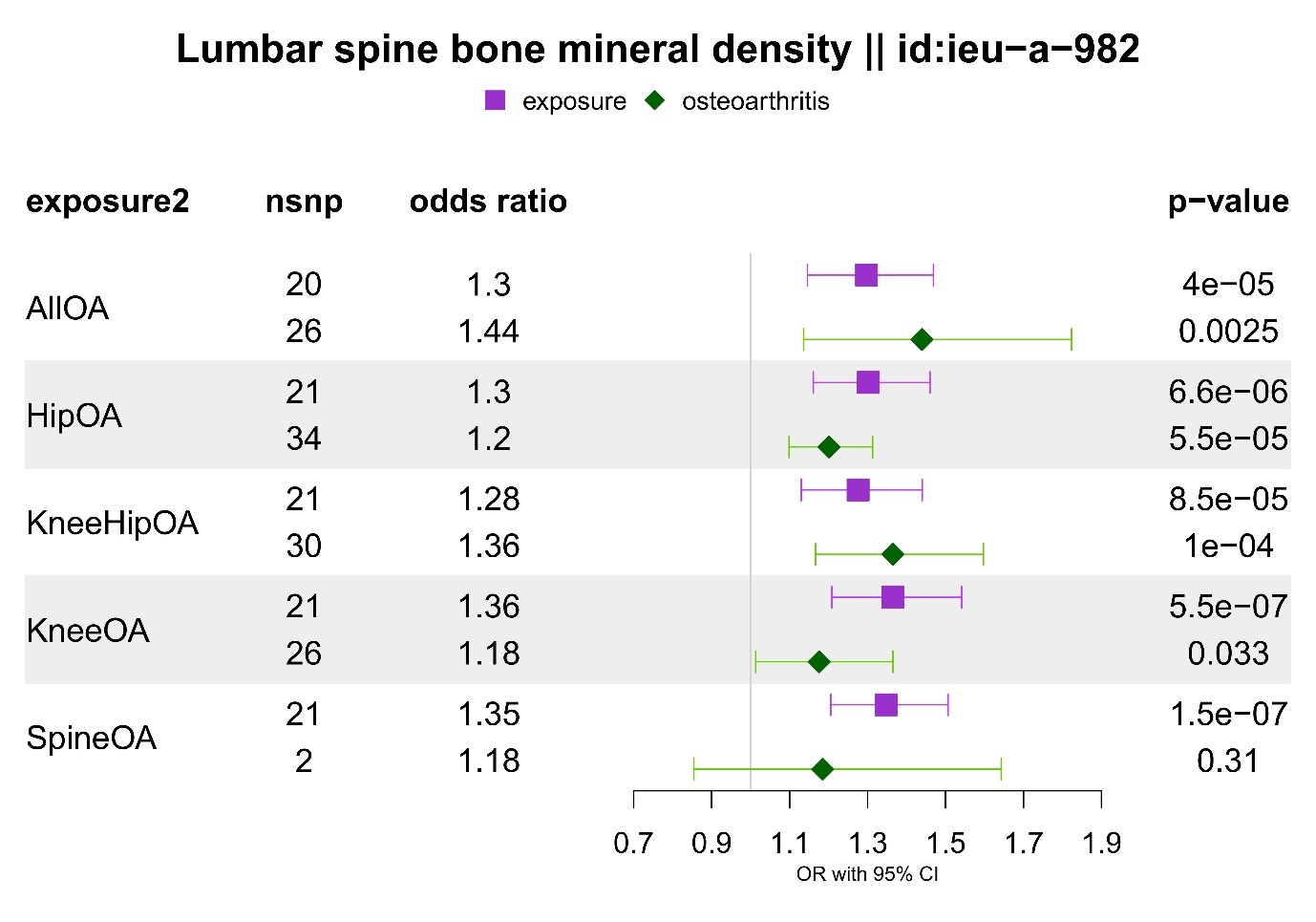

**Supplementary Figure 33.** Multivariable Mendelian randomization results for the jointly modelled effect of genetic susceptibility for lumbar spine bone mineral density and liability for osteoarthritis (all or site-specific) on spinal stenosis (UKBB). The odds ratios are scaled per SD increase of BMI and doubling in the risk of osteoarthritis.

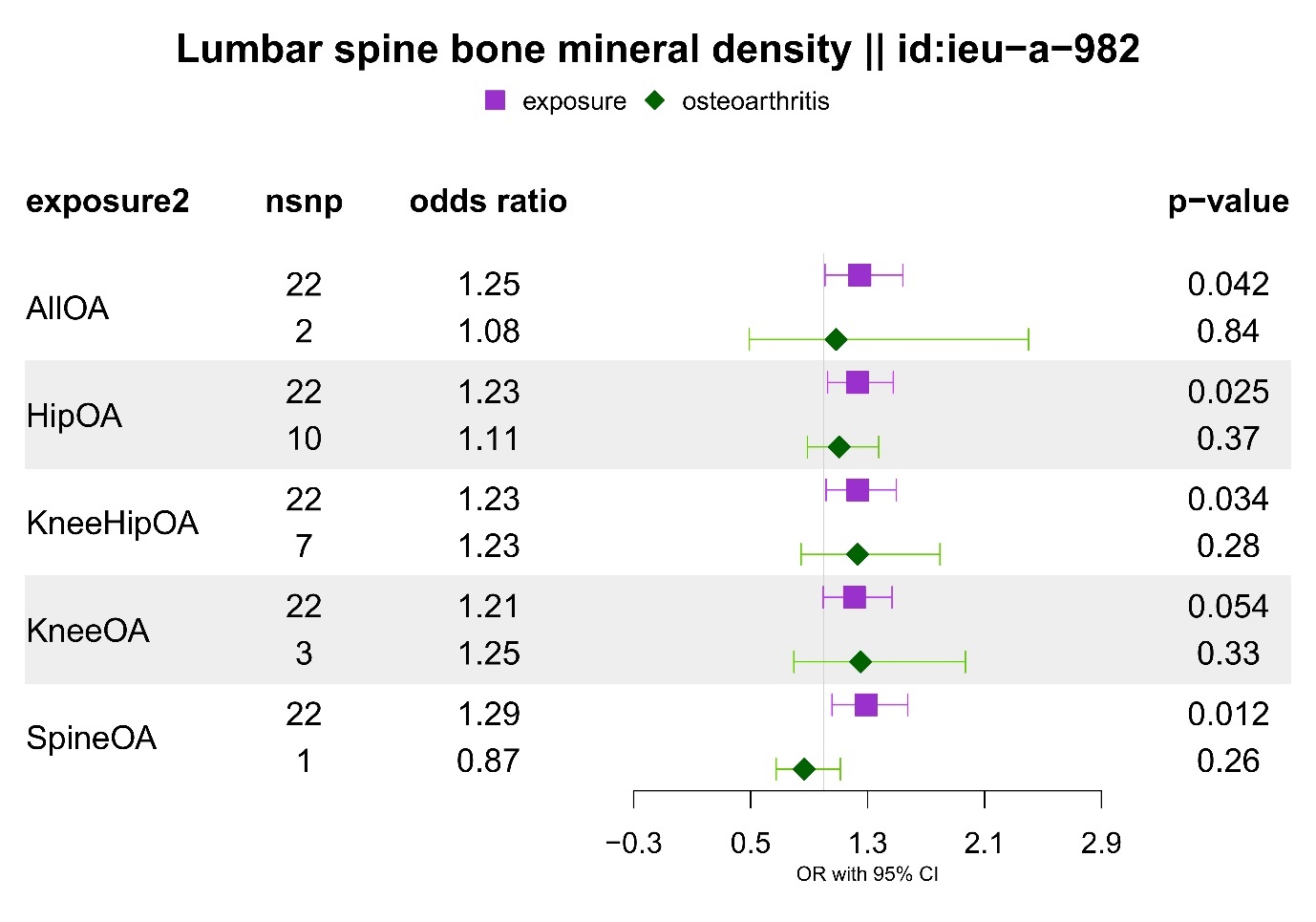

**Supplementary Figure 34.** Multivariable Mendelian randomization results for the jointly modelled effect of genetic susceptibility for risk factors (waist and hip circumference) and body mass index on spinal stenosis (UKBB). The odds ratios are scaled per SD increase of risk factors.

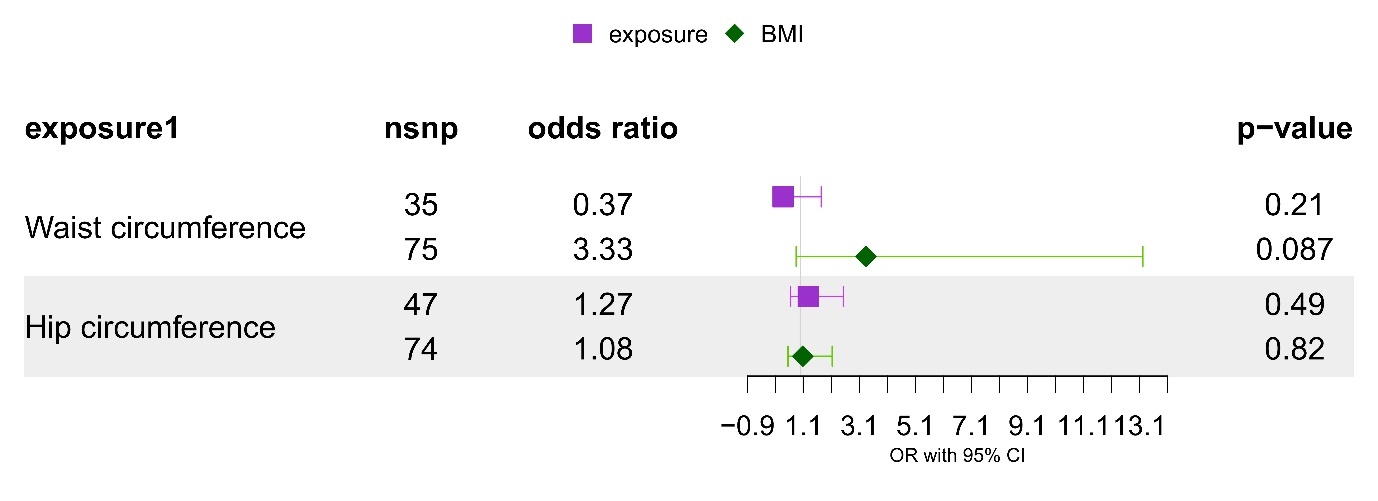

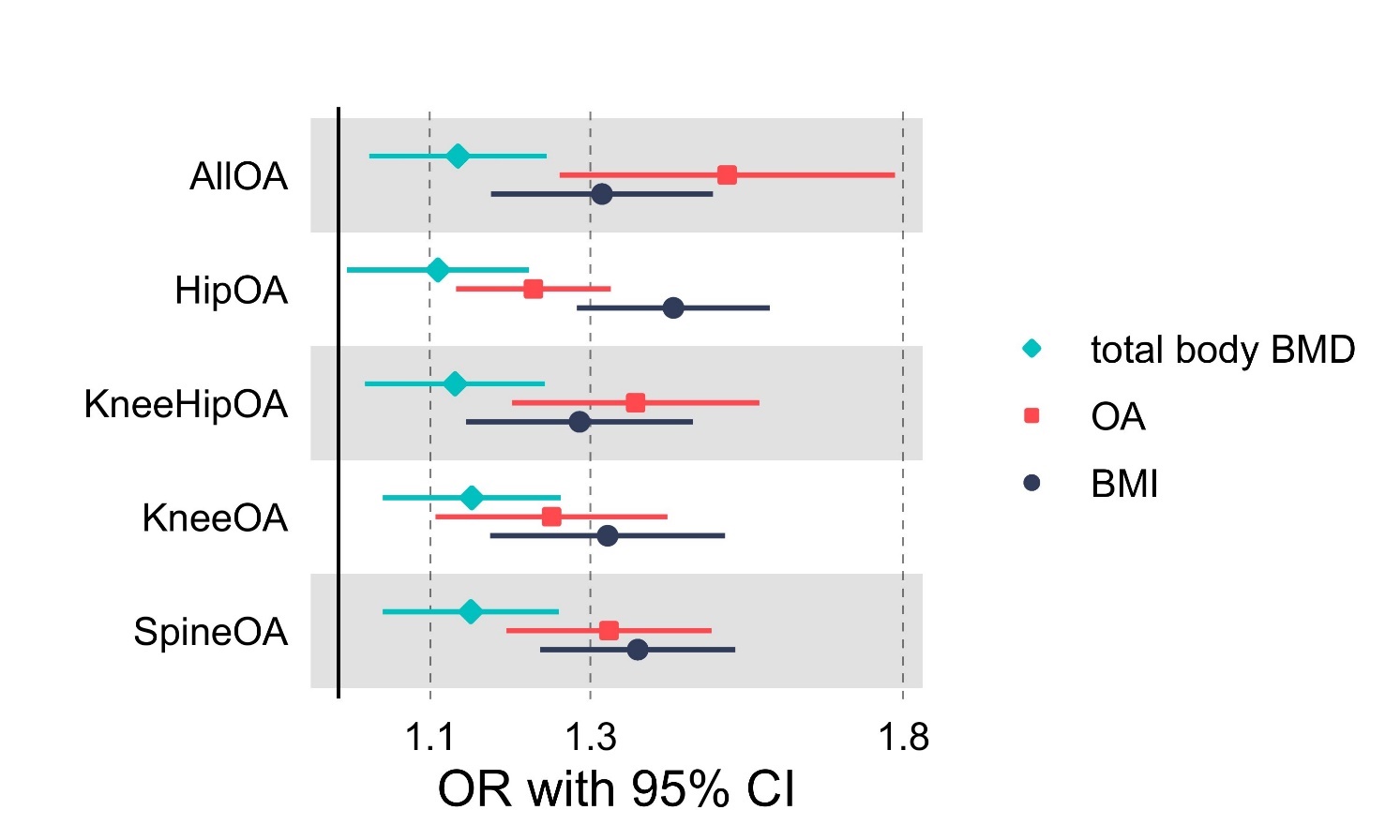
**Supplementary Figure 35.** Multivariable Mendelian randomization results for the jointly modelled effect of genetic susceptibility for total body BMD, BMI and liability for osteoarthritis (all or site-specific) on spinal stenosis (FinnGen). The odds ratios are scaled per SD increase of risk factors and doubling in the risk of osteoarthritis. Filled point shapes represent p-value < 0.05.

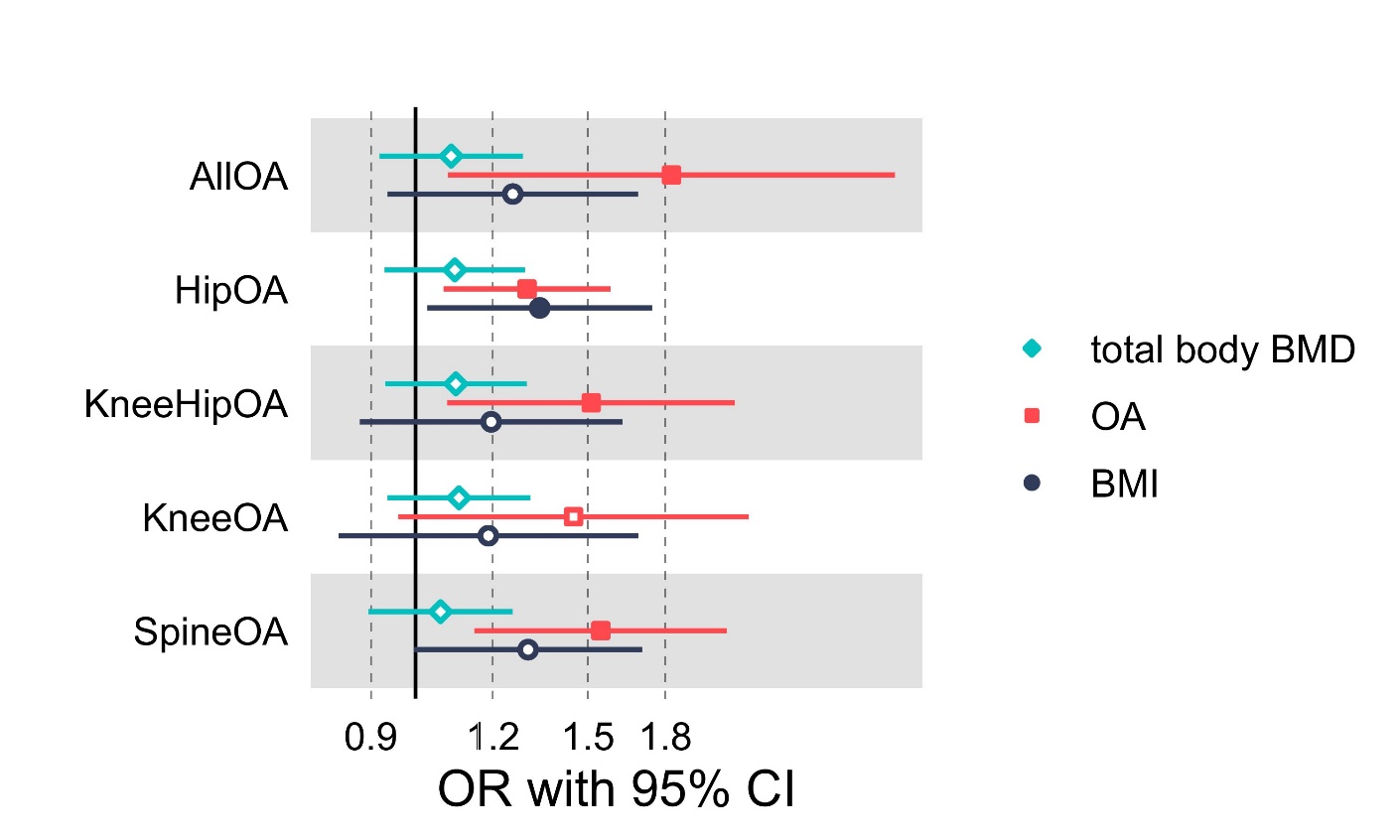
**Supplementary Figure 36.** Multivariable Mendelian randomization results for the jointly modelled effect of genetic susceptibility for total body BMD, BMI and liability for osteoarthritis (all or site-specific) on spinal stenosis (UKBB). The odds ratios are scaled per SD increase of risk factors and doubling in the risk of osteoarthritis. Filled point shapes represent p-value < 0.05.

**Supplementary Figure 37.** Multivariable Mendelian randomization results for the jointly modelled effect of genetic susceptibility for femoral neck BMD, BMI and liability for osteoarthritis (all or site-specific) on spinal stenosis (FinnGen). The odds ratios are scaled per SD increase of risk factors and doubling in the risk of osteoarthritis. Filled point shapes represent p-value < 0.05.

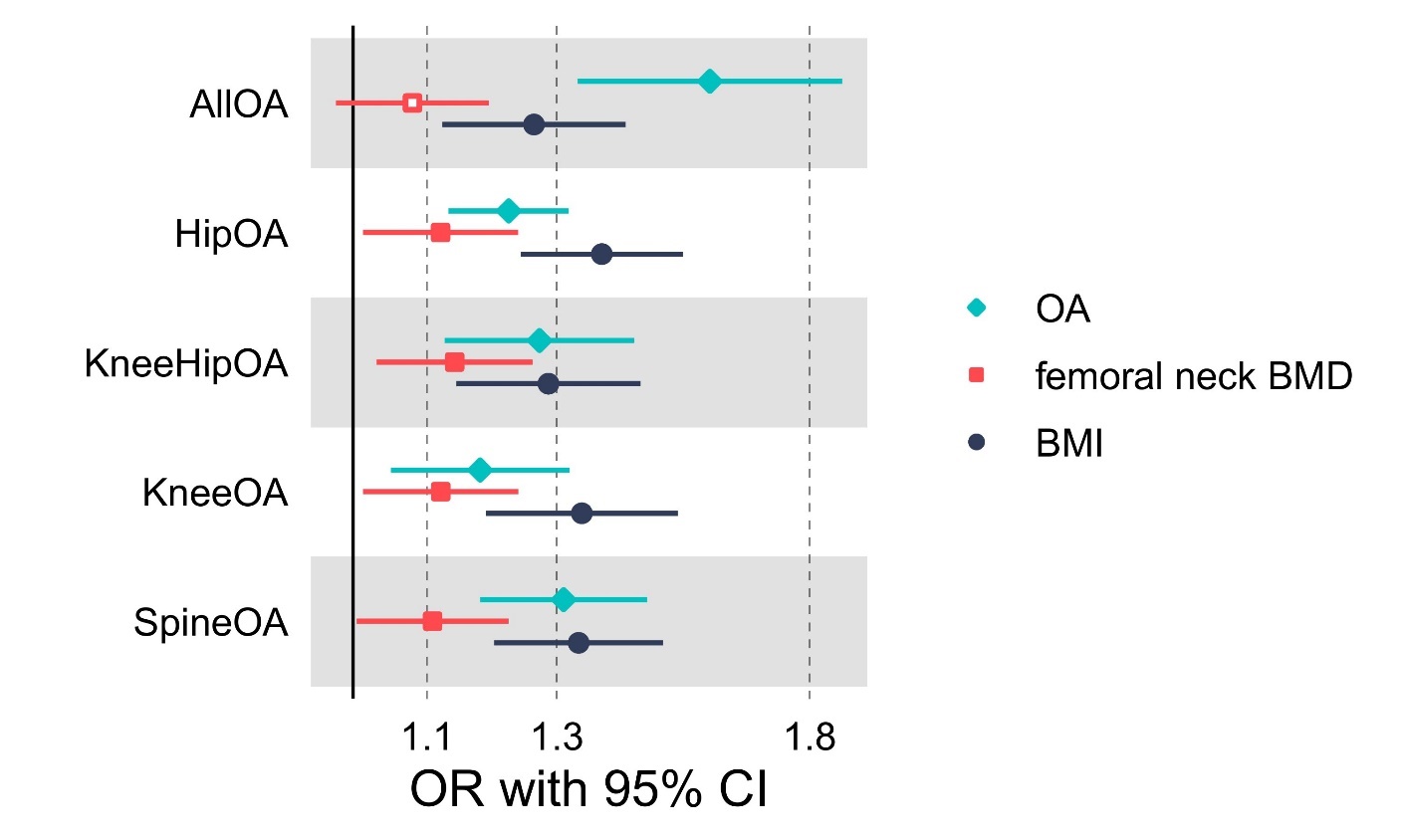

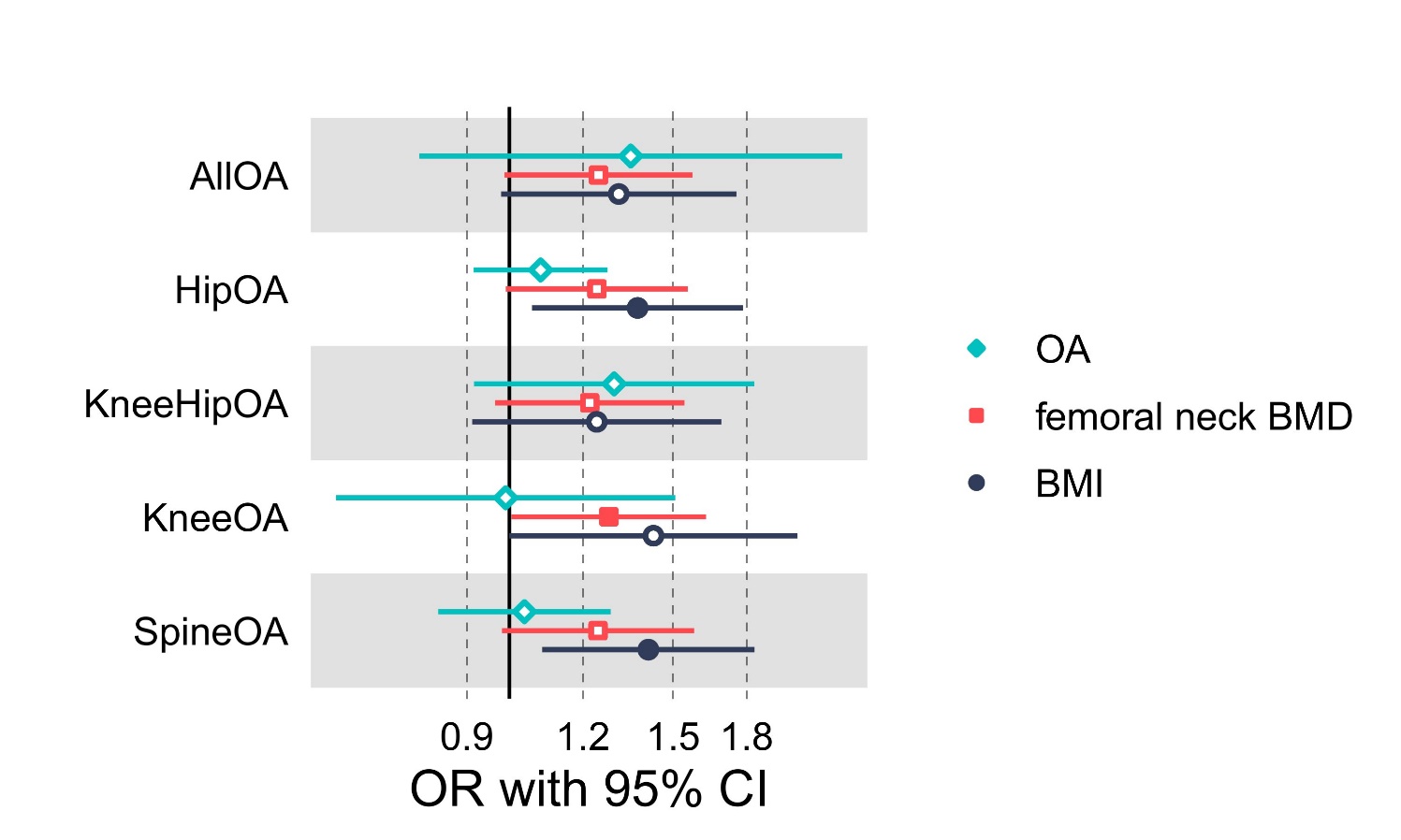
**Supplementary Figure 38.** Multivariable Mendelian randomization results for the jointly modelled effect of genetic susceptibility for femoral neck BMD, BMI and liability for osteoarthritis (all or site-specific) on spinal stenosis (UKBB). The odds ratios are scaled per SD increase of risk factors and doubling in the risk of osteoarthritis. Filled point shapes represent p-value < 0.05.

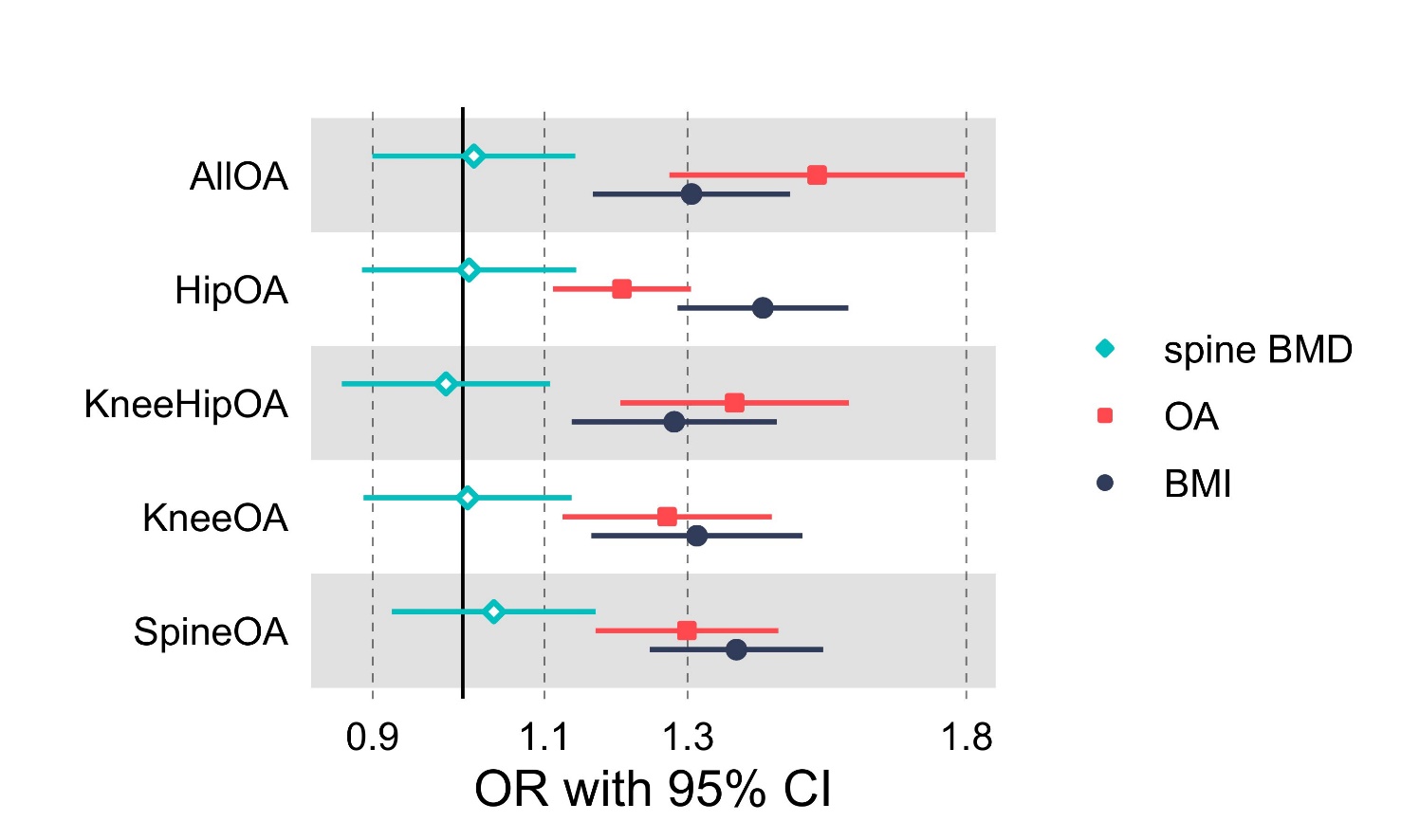
**Supplementary Figure 39.** Multivariable Mendelian randomization results for the jointly modelled effect of genetic susceptibility for lumbar spine BMD, BMI and liability for osteoarthritis (all or site-specific) on spinal stenosis (FinnGen). The odds ratios are scaled per SD increase of risk factors and doubling in the risk of osteoarthritis. Filled point shapes represent p-value < 0.05.

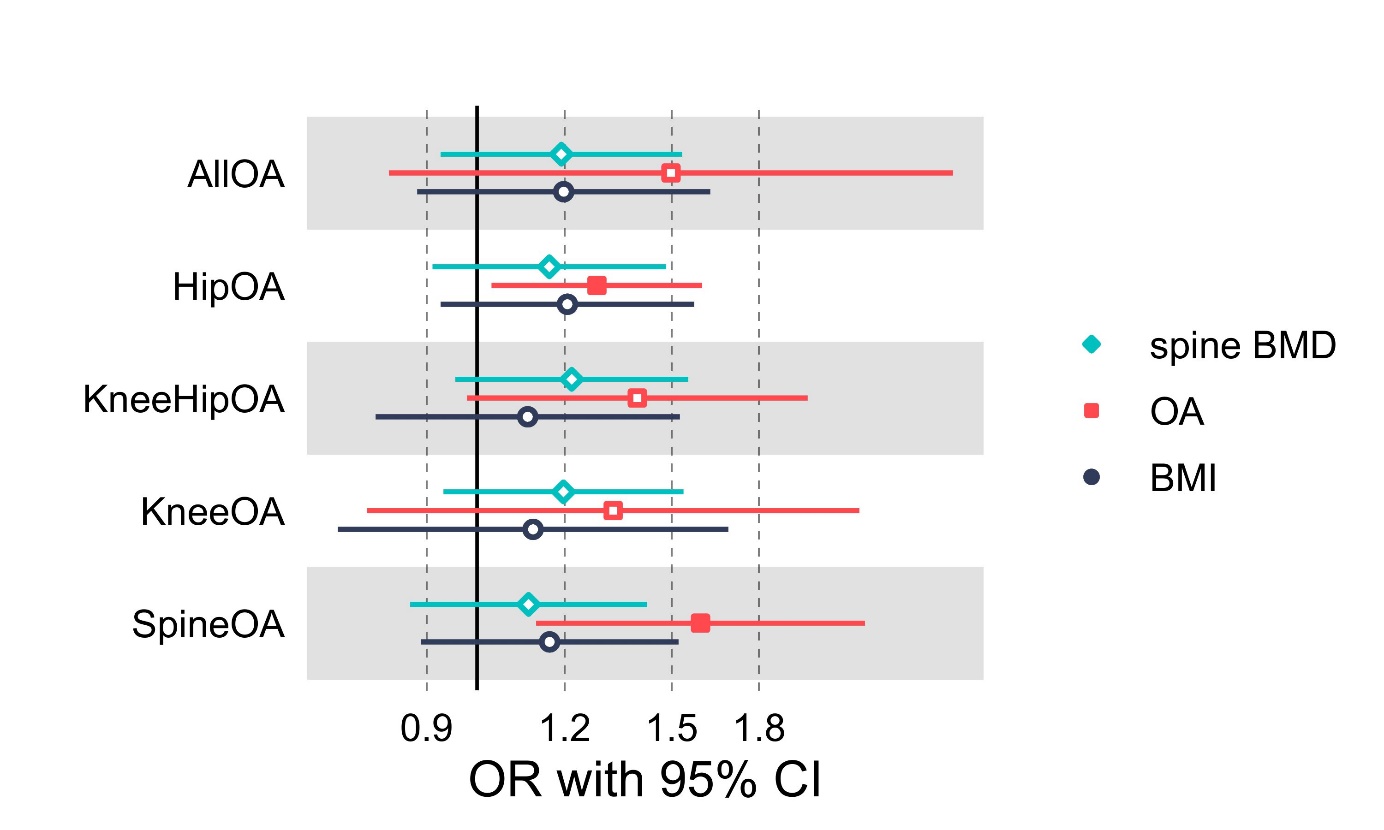
**Supplementary Figure 40.** Multivariable Mendelian randomization results for the jointly modelled effect of genetic susceptibility for lumbar spine BMD, BMI and liability for osteoarthritis (all or site-specific) on spinal stenosis (UKBB). The odds ratios are scaled per SD increase of risk factors and doubling in the risk of osteoarthritis. Filled point shapes represent p-value < 0.05.
