## Supplementary Methods for "Causal relationships between anthropometric traits, bone mineral density, osteoarthritis and spinal stenosis: a Mendelian randomization investigation"

**Femoral neck bone mineral density GWAS**

### Population and phenotype measurement

The UK Biobank (UKBB) is a prospective cohort study with phenotypic and genetic data collected on approximately 500,000 individuals from the United Kingdom, aged between 40 and 69 years at baseline (2006–2010)[1]. The UKB Ethics Advisory Committee oversees the maintenance, development, and use of UKB data, and its approval covers this study. All subjects provided informed consent before participation. Following standardised manufacturers’ protocol high‐resolution dual-energy X-ray absorptiometry scans (iDXA GE‐Lunar, Madison, WI, USA) of both hips are being collected as part of the Imaging Enhancement study[2](follow‐up 2), which began in 2014.

FN BMD derived by DXA was inspected for any potential outliers or anomalous values. An average of left and right FN BMD measurement was calculated if both were measured or left/right used if only one hip was scanned.

### Genotyping and QC

Genotyping, imputation and quality control (QC) were performed by UKB as previously described. Samples were genotyped using two genotyping arrays; Applied Biosystems UK BiLEVE Axiom Array by Affymetrix (49,950 participants) and Applied Biosystems UK Biobank Axiom Array (438,427 participants). Data were imputed using the HRC reference panel, and the merged UK10K and 1000 Genomes phase 3 reference panels in IMPUTE4.
A subset of European individuals from UKB was used for FN BMD GWAS. Ancestry assignment of UKB participants was performed as follows: the UKB sample was projected onto the first 20 principal components estimated from the 1000 Genomes Phase 3 (1000G) project (where ancestry was known) using GCTA version 1.93.2. Projections used a curated set of 38,512 LD-pruned HapMap 3 Release 3 (HM3) REF bi-allelic SNPs that were shared between the 1000G and UKB genotyped datasets (i.e. MAF > 1%, minor allele count > 5, genotyping call rate > 95%, Hardy-Weinberg P > 1x10^-6^, and regions of extensive LD removed). Uniform Manifold Approximation and Projection for Dimension Reduction (UMAP) was used in conjunction with the first 20 principal components to cluster 486,445 individuals using the following parameters: min_dist=0.0001, n_components=3, n_neighbors=45, random_state=10293082. UKB participants that clustered together with the 1000G European sub-populations were manually identified by visual inspection (N=461,920) and used for downstream genetic analyses.

### Genome-wide association analysis

FN-BMD was stratified by sex and adjusted for age, age^2^, 20 ancestry principal components and genotyping chip using ordinary least squares linear regression. Male and female residuals were extracted, standardised (mean=0, SD=1) and then combined to produce the outcome variable for GWAS analysis. We tested genetic variants for association with FN BMD assuming an additive allelic effect, in a linear mixed model implemented in BOLT-LMM[3] v2.3.4 to account for cryptic population structure and relatedness. Only SNPs with an info-score threshold of > 0.4 were analysed.

### **Albumin-corrected calcium GWAS**

Albumin and calcium were measured in the serum in the UK Biobank cohort. The albumin-corrected calcium level was calculated by the following format: corrected calcium = calcium + 0.0177*(45.2- albumin). The resulting corrected calcium phenotype was then filtered by removing outliers beyond 4 standard deviations (SD). We also removed highly-related individuals and those who had withdrawn consent, leaving 424,537 participants in the GWAS analysis. Albumin-corrected calcium GWAS was conducted by using the custom IEU UK Biobank GWAS pipeline[4]. BOLT-LMM, which took population stratification and relatedness into consideration, was used for the GWAS analysis. The model included genotyping chip, sex, and age as covariates.

*References*

1	Bycroft C, Freeman C, Petkova D, *et al.* The UK Biobank resource with deep phenotyping and genomic data. *Nature* 2018;**562**:203–9. doi:10.1038/s41586-018-0579-z

2	Littlejohns TJ, Holliday J, Gibson LM, *et al.* The UK Biobank imaging enhancement of 100,000 participants: rationale, data collection, management and future directions. *Nat Commun* 2020;**11**:2624. doi:10.1038/s41467-020-15948-9

3	Loh P-R, Tucker G, Bulik-Sullivan BK, *et al.* Efficient Bayesian mixed-model analysis increases association power in large cohorts. *Nat Genet* 2015;**47**:284–90. doi:10.1038/ng.3190

4	Mitchell R, Elsworth BL, Raistrick CA, *et al.* MRC IEU UK Biobank GWAS pipeline version 2. *Bristol, UK Univ Bristol* 2019.
